# Comprehensive adjudication identifies 111 high-confidence loci for Alzheimer’s disease and related dementias

**DOI:** 10.64898/2026.05.14.26353247

**Authors:** Yuk Yee Leung, Edoardo M. Marcora, Adam Naj, Tulsi Patel, Kathy Sedgwick, Zivadin Katanic, Ryan M. Corces, Li-San Wang, Richard Mayeux, Alison Goate, Lindsay Farrer, Gerard D. Schellenberg, Brian Kunkle, Badri Vardarajan

## Abstract

**Background:** The Alzheimer’s Disease Sequencing Project Gene Verification Committee developed a systematic framework to adjudicate genetic evidence for AD and related dementias, addressing wide variation in association quality.

**Methods:** Phase 1 established tiered criteria by evaluating 23 nominated loci across study designs. Phase 2 applied this framework to 29 large-scale genome-wide studies published since 2015, tiering 163 unique loci.

**Results:** Phase 1 yielded 17 high-confidence loci (12 linked to specific genes), and Phase 2 identified 111 high-confidence loci/genes with replicated associations across ancestries and convergent single-variant/variant-set evidence. Prioritized loci highlight APP processing, microglial immunity, and lipid metabolism pathways, including genes not captured by existing resources like Agora or Open Targets. Summarized results can be viewed at https://topgenes.niagads.org/.

**Conclusion:** This rigorously adjudicated catalog represents the most comprehensive AD/ADRD genetics resource to date, providing a foundation for functional validation and therapeutic discovery with broad applicability to complex diseases.

## Introduction

One of the primary goals of the Alzheimer’s Disease Sequencing Project (ADSP) is to identify genetic variants that increase or reduce the risk of Alzheimer’s disease (AD) to elucidate biological mechanisms and identify novel potential therapeutic targets for AD and AD-related dementias (ADRD). The ADSP Gene Verification Committee (GVC) (**Appendix 1**) was established to evaluate genetic signals for AD and AD-related phenotypes. Retrospective analyses show that therapeutic targets supported by genetic evidence are 2–3 times more likely to succeed compared to targets not supported by genetic findings^1–8^. Thus, genes discovered through genome-wide association studies (GWAS) and sequence-based studies of AD and ADRD are a validated source of potential therapeutic targets. Developing a therapeutic target from a gene is a resource-intensive process. Therefore, it is critical that the genetic evidence supporting a target be carefully evaluated.

A large number of publications report evidence of genetic loci associated with AD and dementia^9–37^. However, the quality of evidence presented is highly variable. To clarify which loci are potential true signals versus false positives, the GVC (**Appendix 1**) established criteria for ranking evidence supporting a locus or gene. These criteria were developed by evaluating a list of loci and genes from different types of studies (**Phase 1,** Box 1, **Table 1**).

**Table 1.** Twenty-three loci evaluated in Phase 1. Phase 1 loci were nominated by GVC members, selected to represent different types of study design. The evidence category column summarizes the kind of evidence supporting the nomination. We identified 17 high-confidence loci with sufficient evidence for association (Tier 1), and 12 of these were associated with a specific gene with high confidence (“Yes” in “Locus=Gene” column) (see **Box 1** and **Appendices 2** and **3** for ranking criteria).

| Gene | Evidence category | Tier of Locus<sup>a</sup> | Locus = Gene |
| --- | --- | --- | --- |
| *ABCA7* | GWAS | 1 | Yes |
| *ABI3* | Sequence-based (WGS/WES) or exome chip | 1 | Yes |
| *ADAM10* | Targeted sequencing | 2 |  |
| *APOE* | Linkage study and association studies | 1 | Yes |
| *APP* | Linkage study, Monogenic families | 1 | Yes |
| *BIN1* | GWAS | 1 | Yes |
| *CASP7* | Sequence-based (WGS/WES) or exome chip | 1 | Yes |
| *CD33* | Sequence-based (WGS/WES) or exome chip | 3 |  |
| *CR1* | GWAS | 1 |  |
| *HLA-DRB5* | GWAS | 1 |  |
| *IGHV1-67* | Region/gene-based analysis | 3 |  |
| *MS4A4A/MS4A6A* | GWAS | 1 |  |
| *PLCG2* | Sequence-based (WGS/WES) or exome chip | 1 | Yes |
| *PPARGC1A* | Region/gene-based analysis | 3 |  |
| *PSEN1* | Linkage study, Monogenic families | 1 | Yes |
| *PSEN2* | Linkage study, Monogenic families | 1 | Yes |
| *RORA* | Region/gene-based analysis | 3 |  |
| *SCARA3* | Region/gene-based analysis | 3 |  |
| *SORL1* | Targeted sequencing/Monogenic families/GWAS | 1 | Yes |
| *SPI1* | GWAS | 1 | Yes |
| *TP53INP1* | Region/gene-based analyses | 1 |  |
| *TREM2* | Sequence-based (WGS/WES) or exome chip, | 1 | Yes |
|  | Targeted sequencing |  |  |
| *ZNF423* | Region/gene-based analyses | 3 |  |
| <sup>a</sup> Tier assignment follows criteria outlined in Box 2. See Appendix 4 for more details. |  |  |  |

Expanding the criteria developed in Phase 1, we then performed a structured literature search (**Phase 2, Figure 1**) to identify publications reporting AD and dementia association signals and evaluate the strength of the evidence supporting these signals (**Phase 2, Table 2**). A list of high-confidence genes and/or loci was developed from Phases 1 and 2 (**Tables 1 and 3**) that consisted of 12 genes and 97 loci. All genes in **Table 1** overlap with signals in **Table 3** except *PSEN1* and *PSEN2*. Signals at *APOC1, APOC1P1, APOC2, APOC4, TOMM40* and *PVRL* are close to *APOE* and may simply reflect linkage disequilibrium (LD) with the primary gene, *APOE*.

**Figure 1.**
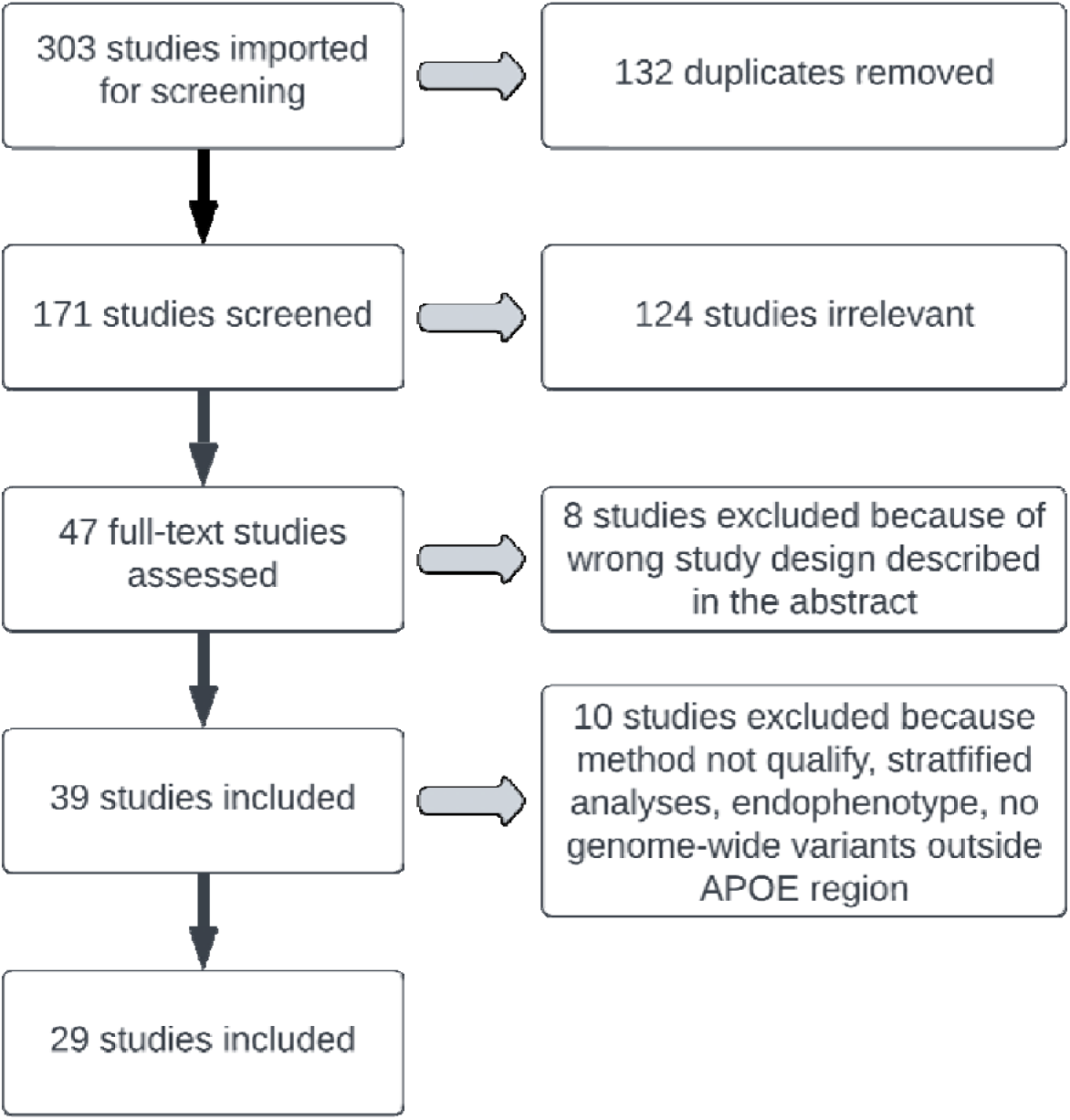
Structured literature search for Phase 2. Studies were identified through systematic literature search via GWAS catalog and PubMed databases using AD-related Experimental Factor Ontology IDs. A total of 303 studies were retrieved; 132 were removed as duplicates, leaving 171 studies for title and abstract screening. Of these, 124 studies were deemed irrelevant based on the inclusion/exclusion criteria. 47 full-text studies were assessed. Among these, 8 studies were excluded because of wrong study design described in the abstract. Upon detailed review by the GVC, 10 studies were excluded for reasons described below. The remaining 29 studies (details shown in **Table 2**) are publications focused on in this study.

**Table 2.** Publications evaluated for Phase 2. Twenty-nine publications were included for adjudication in Phase 2 (Figure 1). Twenty-three were evaluated using the single-variant (SV) criteria, 12 were evaluated using the variant-set (VS) criteria (**Box 3**), and 2 were evaluated using the “Family” criteria (**Box 4**). Twenty publications analyzed AD-only data, 6 analyzed “AD+Dementia,” and 3 focused on “Dementia” subjects. There are 5 multi-ancestry studies. For the remaining publications, 3 included Asian ancestry samples, 1 included African ancestry samples, and the rest included non-Hispanic white-European ancestry (NHW-EA) samples. Ancestry is defined using a combination of self-identified or ascribed and inclusion criteria provided by the authors.

| PMCID Link | Study | Evaluation Criteria | Phenotype | Ancestry |
| --- | --- | --- | --- | --- |
| PMC7573798 | Kunkle 2021<sup>9</sup> | SV, VS | AD | African |
| PMC4554442 | Tosto 2015<sup>10</sup> | SV | AD | Multi-ancestry<sup>a</sup> |
| DOI: 10.1097/YPG.000000000000090 | Hirano 2015<sup>11</sup> | VS | AD | Asian |
| PMC7925686 | Shigemizu 2021<sup>12</sup> | SV | AD | Asian |
| PMC9135624 | Shigemizu 2022<sup>13</sup> | SV | AD | Asian |
| PMC5496797 | Jun 2017<sup>14</sup> | SV, VS | AD | Multi-ancestry<sup>b</sup> |
| PMC4970971 | Herold 2016<sup>15</sup> | Family | AD | NHW-EA |
| PMC6043775 | Raghavan 2018<sup>16</sup> | VS | AD | Multi-ancestry<sup>b</sup> |
| PMC5318231 | Mez 2017<sup>17</sup> | SV | AD | NHW-EA |
| PMC5669039 | Sims 2017<sup>18</sup> | SV | AD | NHW-EA |
| PMC5959890 | Marioni 2018<sup>19</sup> | SV | AD+Dementia | NHW-EA |
| PMC6463297 | Kunkle 2019<sup>20</sup> | SV | AD | NHW-EA |
| PMC6330399 | Nazarian 2019<sup>21</sup> | SV, VS | AD | NHW-EA |
| PMC6503572 | Tosto 2019<sup>22</sup> | VS | AD | Multi-ancestry<sup>b</sup> |
| PMC6408965 | Zhang 2019<sup>23</sup> | SV, VS | AD | NHW-EA |
| PMC6836675 | Jansen 2019<sup>24</sup> | SV | AD+Dementia | NHW-EA |
| PMC7081222 | Prokopenko 2020<sup>25</sup> | SV | AD | Multi-ancestry<sup>c</sup> |
| 10.1016/j.jalz.2019.06.4950 | Moreno-Grau 2019 <sup>26</sup> | SV | Dementia | NHW-EA |
| PMC6375806 | Bis 2020 <sup>27</sup> | SV, VS | AD | NHW-EA |
| PMC8204704 | Katsumata 2020 <sup>28</sup> | SV, VS | AD | NHW-EA |
| PMC8017764 | Le Guen 2021 <sup>29</sup> | SV | AD | NHW-EA |
| PMC8519060 | Prokopenko 2021 <sup>30</sup> | Family, VS | AD | NHW-EA |
| PMC8184987 | de Rojas 2021 <sup>31</sup> | SV | AD+Dementia | NHW-EA |
| PMC7610386 | Schwartzentru<br>ber 2021 <sup>32</sup> | SV | AD+Dementia | NHW-EA |
| PMC10243600 | Wightman 2021 <sup>33</sup> | SV | AD+Dementia | NHW-EA |
| PMC7910483 | He 2021 <sup>34</sup> | SV | Dementia | NHW-EA |
| PMC8715147 | He 2021 <sup>35</sup> | SV, VS | Dementia | NHW-EA |
| PMC9126808 | Prokopenko 2022 <sup>36</sup> | VS | AD | NHW-EA |
| PMC9005347 | Bellenguez 2022 <sup>37</sup> | SV | AD+Dementia | NHW-EA |

<sup>a</sup> Multi-ancestry groups in this study include NHW-EA, African, and Hispanic/Latino subjects.
<sup>b</sup> Multi-ancestry groups in this study include NHW-EA, African, and Asian subjects.
<sup>c</sup> Multi-ancestry groups in this study include NHW-EA and African subjects.

**Table 3.**
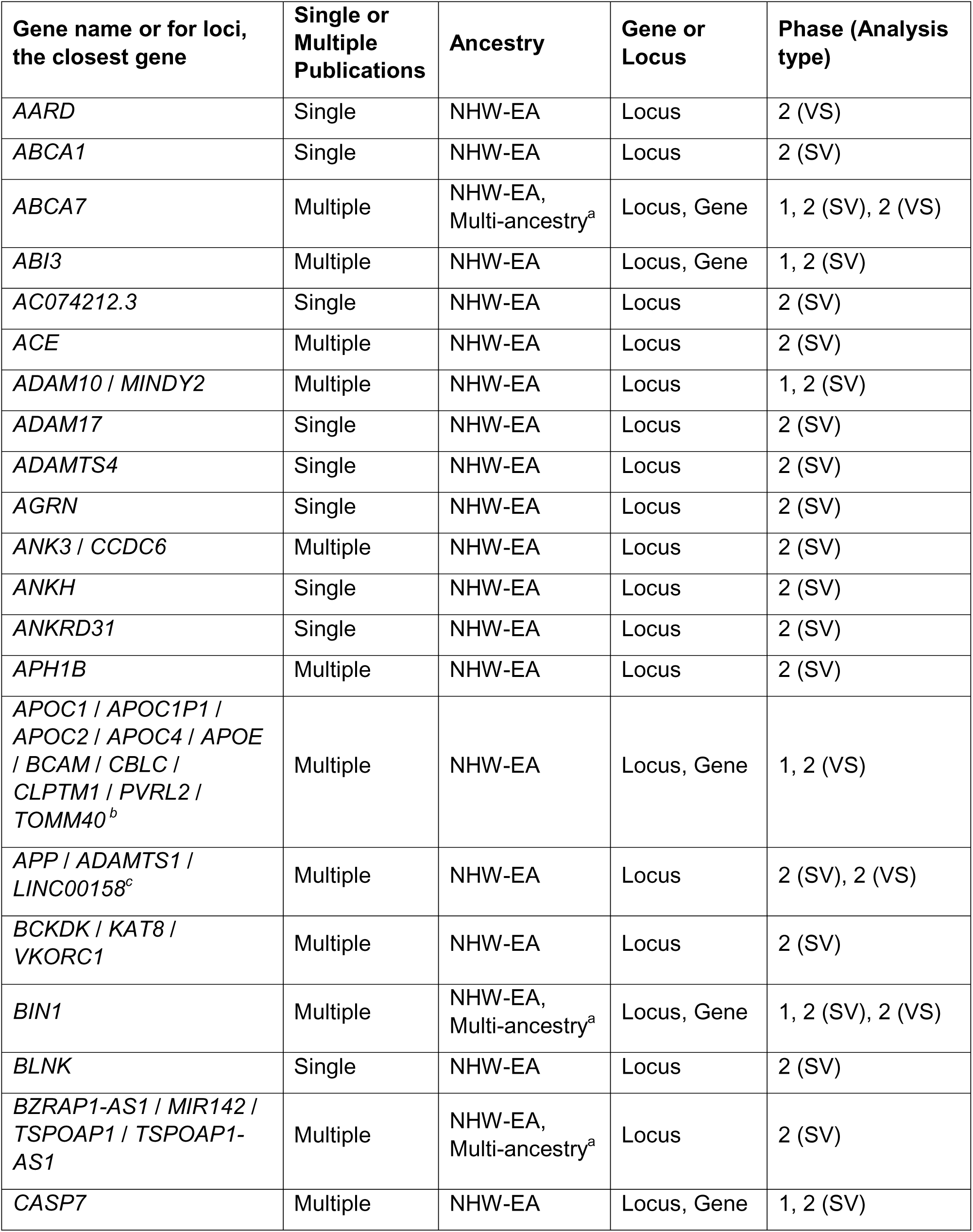
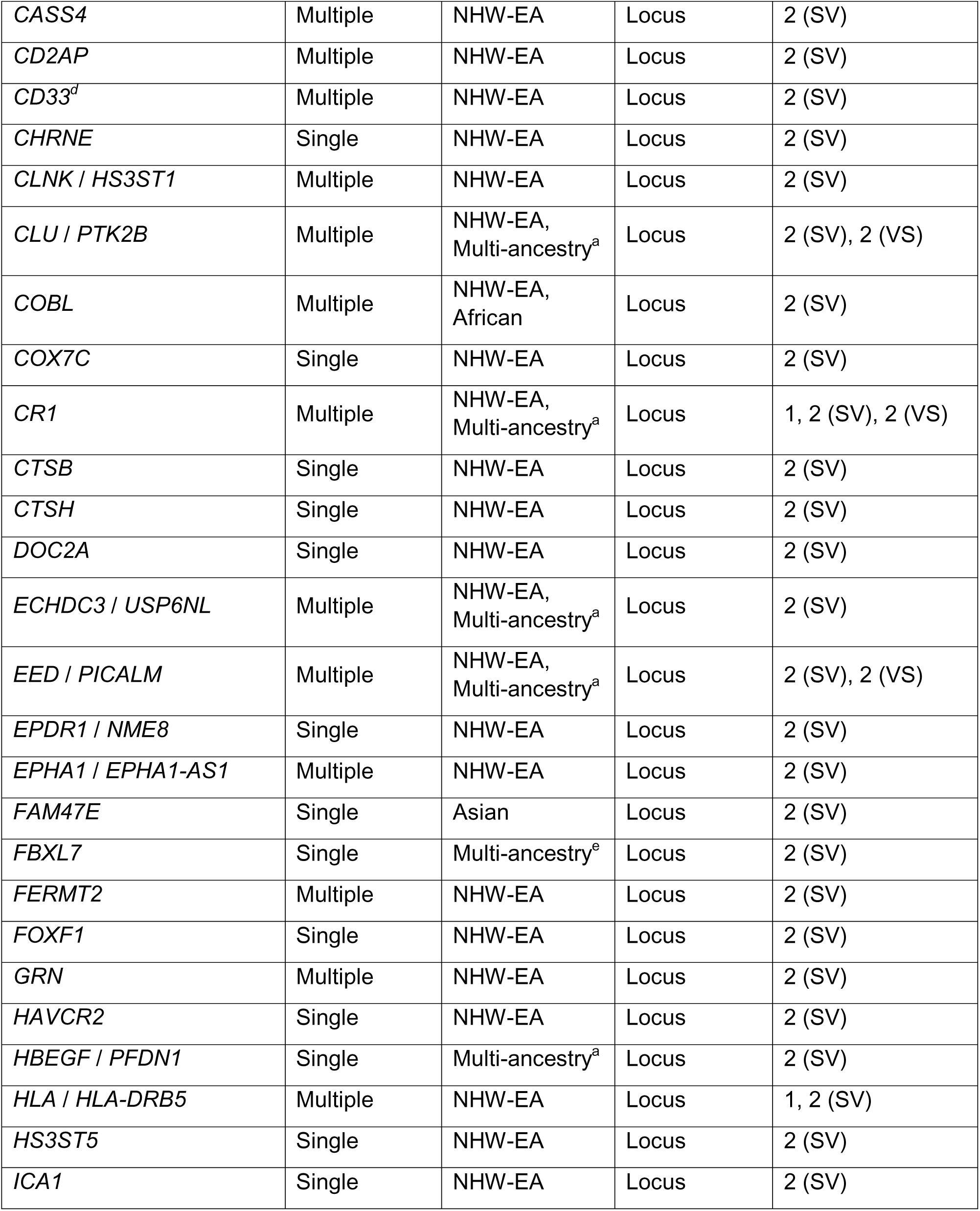
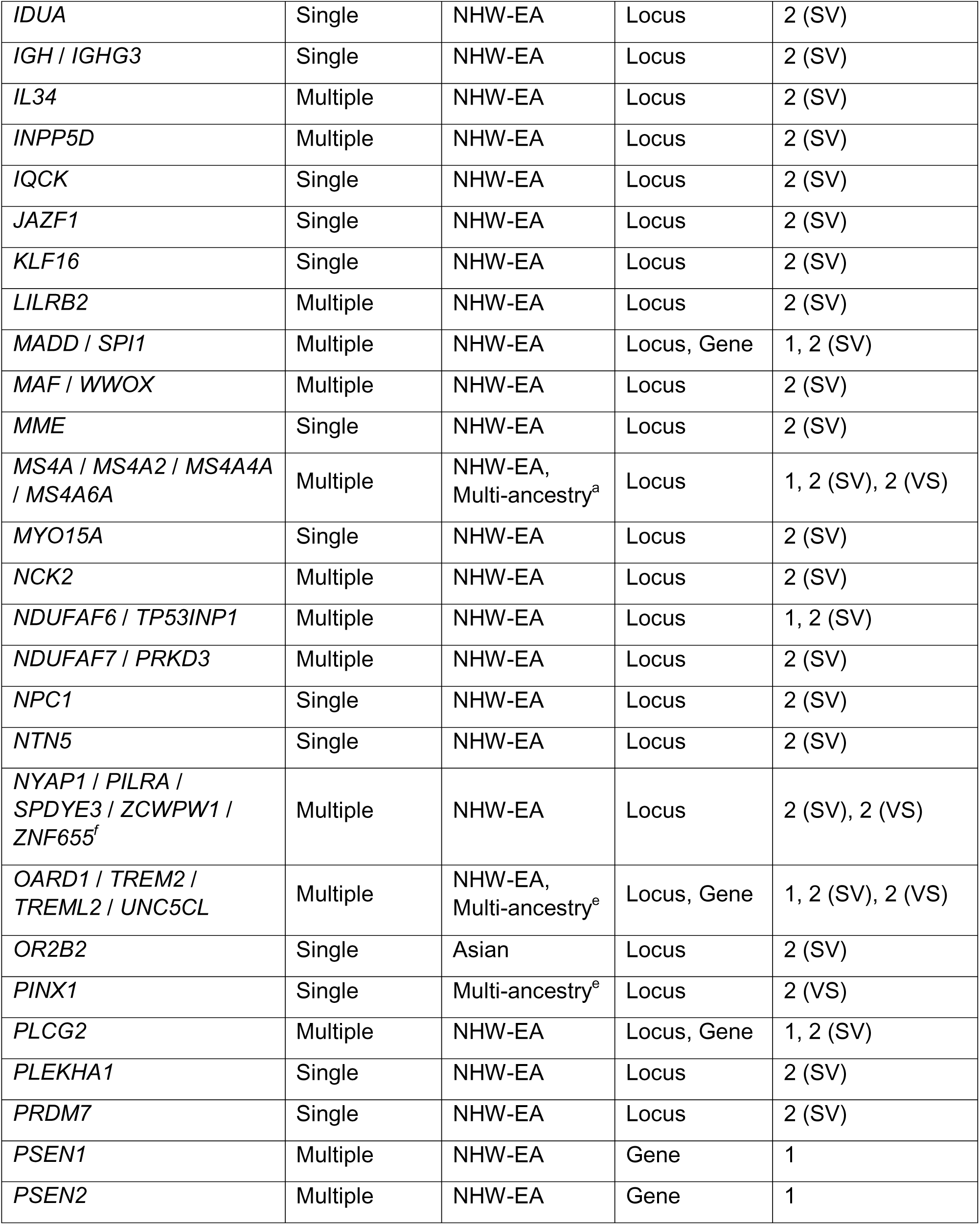
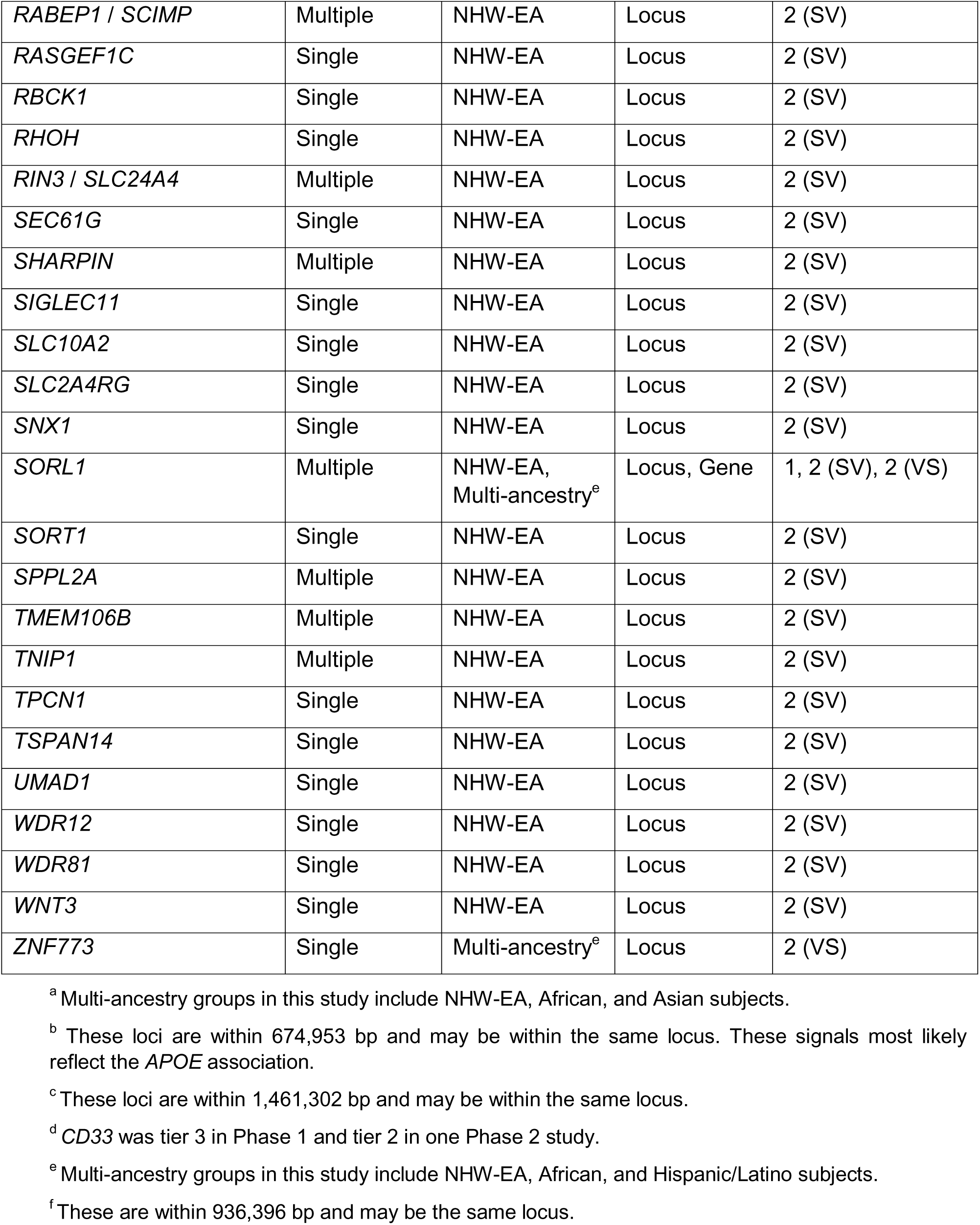
All phase 1 and phase 2 genes and loci with tier 1 or 2 ranking. Some of these genes are also in **Table 1**. Analysis types include single variant (SV) and variant set (VS).

## Results

The GVC evaluated publications where the diagnosis of AD or classification as a control was based on clinical and/or pathological evidence. We also considered studies that included biobank cohorts in which study participants indicated on a family history questionnaire whether their parents had or did not have “…AD/dementia….” Individuals with one or more parents labeled AD/dementia were considered proxy AD cases. Participants with elderly parents who were not identified as having AD/dementia were considered as controls and older controls were assigned higher weights compared to younger controls. Proxy cases and controls were not directly examined, and their medical records were not evaluated. This proxy information is less precise than a clinical or neuropathologic based diagnosis, because of lack of specificity to dementia due to AD, and thus was evaluated as a separate phenotype which encompasses all AD related dementias. A neuropathologic diagnosis of AD with clinical dementia is considered as the gold-standard. A clinical diagnosis, without neuropathologic confirmation, may include other causes of dementia with a similar clinical presentation.

### Phase 1

The GVC selected 23 loci or genes for initial evaluation (**Table 1**). These were selected to represent different study designs including analysis of monogenic forms of AD, linkage analysis in families, gene or region-based studies, targeted sequence evaluations, sequence-based studies including exome chip publications, and GWAS. Information on mutations and structural variants was included in the evaluations. We evaluated evidence supporting an association signal (locus) using a structured tier system for single-variant (SV) reports (**Box 1).** The forms used with additional details are in **Appendix 2**. We also performed an in-depth evaluation of genes nominated for the loci in Phase 1. For monogenic forms of AD, we reviewed evidence for co-segregation of non-synonymous variants and structural variants, and linkage analysis. We also considered variant-set (VS)-based analyses that included gene-based, VS-based, and region-based analyses. For these studies we considered significance levels and the methods used to evaluate the data. For SV association studies, we considered the level of significance, the quality of the study design, and the consistency of results from multiple studies. Other information reviewed included pleiotropy, ancestry of study participants, functional genomics, and gene/protein function (**Appendix 5**). We utilize genetic ancestry classifications as reported in the original publications, whether determined through self-identification or genetic inference methods, and examine the similarities and differences in genetic findings across genetically defined ancestry groups. Though no formal criteria were developed due to the heterogeneity of the evidence reviewed, the GVC reached a consensus that *ABCA7*, *ABI3*, *APOE*, *APP*, *BIN1*, *CASP7*, *PLCG2*, *PSEN1*, *PSEN2*, *SORL1*, *SPI1*, and *TREM2* are genes that contribute to the respective association signals (**Table 1**), although other genes in close proximity to *APOE, SPI1*, and *TREM2* may also contribute to AD risk. For the remaining loci, the specific gene was not assigned. For Phase 1, we found 17 high-confidence loci (Tier 1 or 2) with sufficient evidence for association, with 12 associated with a specific gene with high confidence (**Table 1**).

### Phase 2

Phase 2 was a more comprehensive evaluation of genetic studies that were selected using a structured literature search (**Figure 1**) that returned 304 published peer-reviewed articles. After manually curating the publications to remove duplicates and ineligible reports identified by automated methods, 171 remained that were individually screened by at least two reviewers. Studies were included if they reported at least one locus with suggestive or genome-wide significant evidence for association with AD, proxy AD/dementia, AD+proxy dementia, or dementia. Studies were excluded if the outcomes were not AD or AD/dementia by proxy, or if the total sample size was <1,000 total participants. After this process, 29 studies remained for further evaluation (**Table 2**) including large GWAS^19,20,24,37–39^, genome-wide variant-set (VS) based association studies^14,21,22,27,40^, and whole exome and genome sequence-based studies^13,16,27,30,35,36^.

We developed a tier system for evidence of association with rankings ranging from 1 (strongest) to 7 (weakest). Assignment to specific tiers was based on the information outlined in **Box 2** and the form in **Appendix 4**. For each publication, we considered all loci in the entire dataset (combined or meta-analyses of all stages for the largest sample size in a publication) and assigned a tier to all reported loci with association with the phenotype being considered. When possible, AD defined by clinical/neuropathological criteria was considered separately from dementia defined by proxy data. However, in some reports, results for the two phenotypes could not be separated; for these mixed phenotype studies, dementia was considered the study phenotype. VS-based studies were scored as defined in **Box 3** with an additional criterion that signals were considered for tier 1 only if there were 2 or more significant variants not in strong LD contributing to the signal. For family-based studies (defined for this purpose as >50% of the samples were from families) conducted using linkage analysis, we modified our ranking system (**Box 4**), and a genome-wide significance threshold was defined as a Logarithm of the Odds (LOD) score of >3.6 and suggestive evidence is a LOD score >2.0^41^.

For SV analyses, there were 163 unique loci as grouped by FUMA (Figure 2)^42^ across all phenotypes, tiers, and ancestries (**Supplementary Table 1)**. Of these, 97 (60%) loci received a ranking of 1–2. Non-Hispanic whites with European ancestry (NHW-EA) were by far the largest group studied, with 20 out of 29 studies being exclusively NHW-EA cohorts. For NHW-EA studies of AD risk, there were 29 high-confidence loci (tiers 1 or 2) (**Table 4**, **Supplementary Table 1**). When the disease definition was expanded to AD and proxy-AD, this number increased to 88 loci. Some of these loci were assigned lower tiers based on evidence presented for the following reasons: 1) small sample sizes; 2) fluctuations in the significance of (reported effect size or p-values) results across studies with borderline levels of significance; 3) missing data preventing full evaluation of association evidence; 4) inclusion of individuals who were ascertained by variable methods; and 5) addition of new samples with population differences not detectable using current methods. Studies of other populations including African, Hispanic/Latino, and Asian ancestries identify far fewer top tier loci (∼10%, **Table 3**), principally due to much smaller sample sizes for these groups.

**Figure 2.**
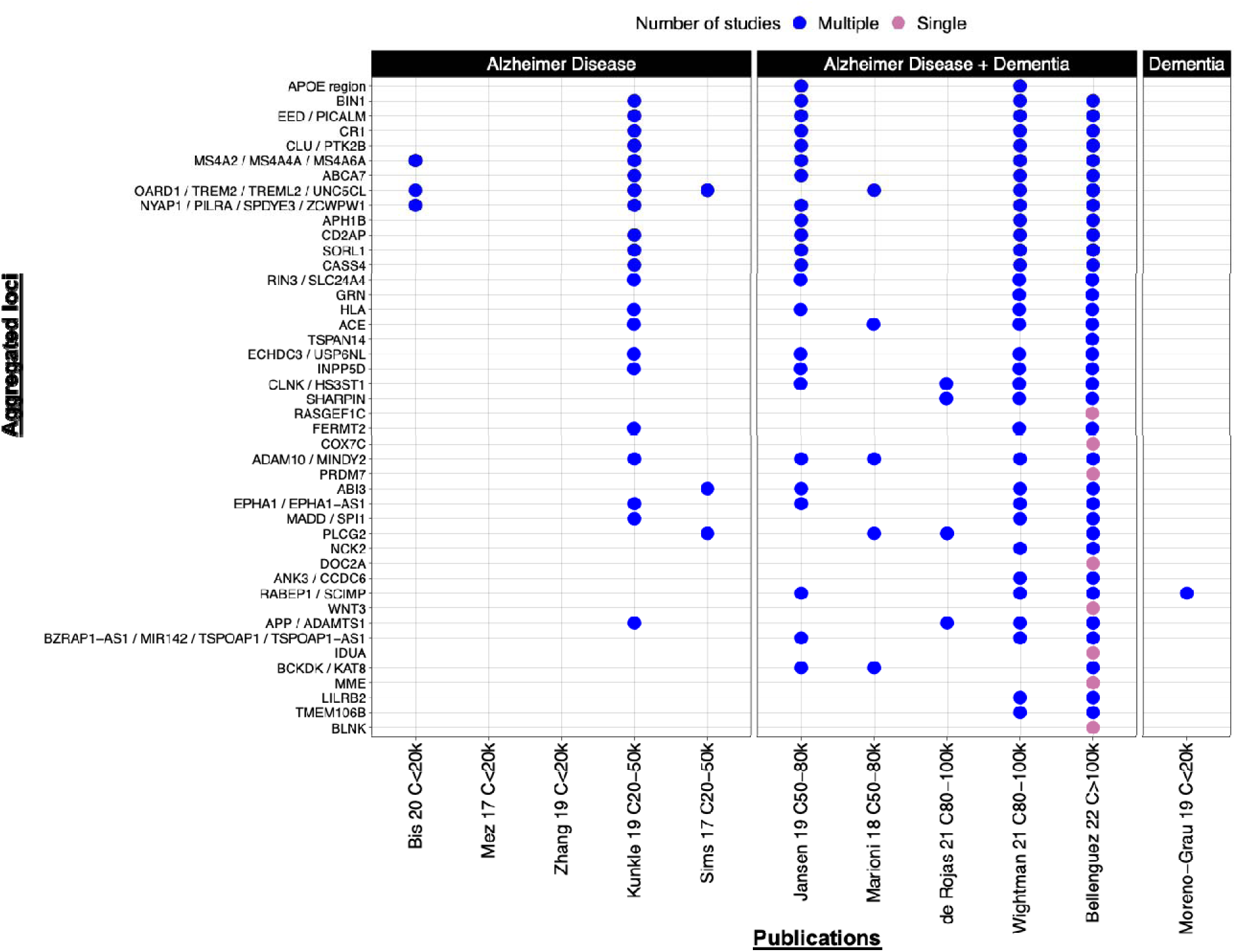
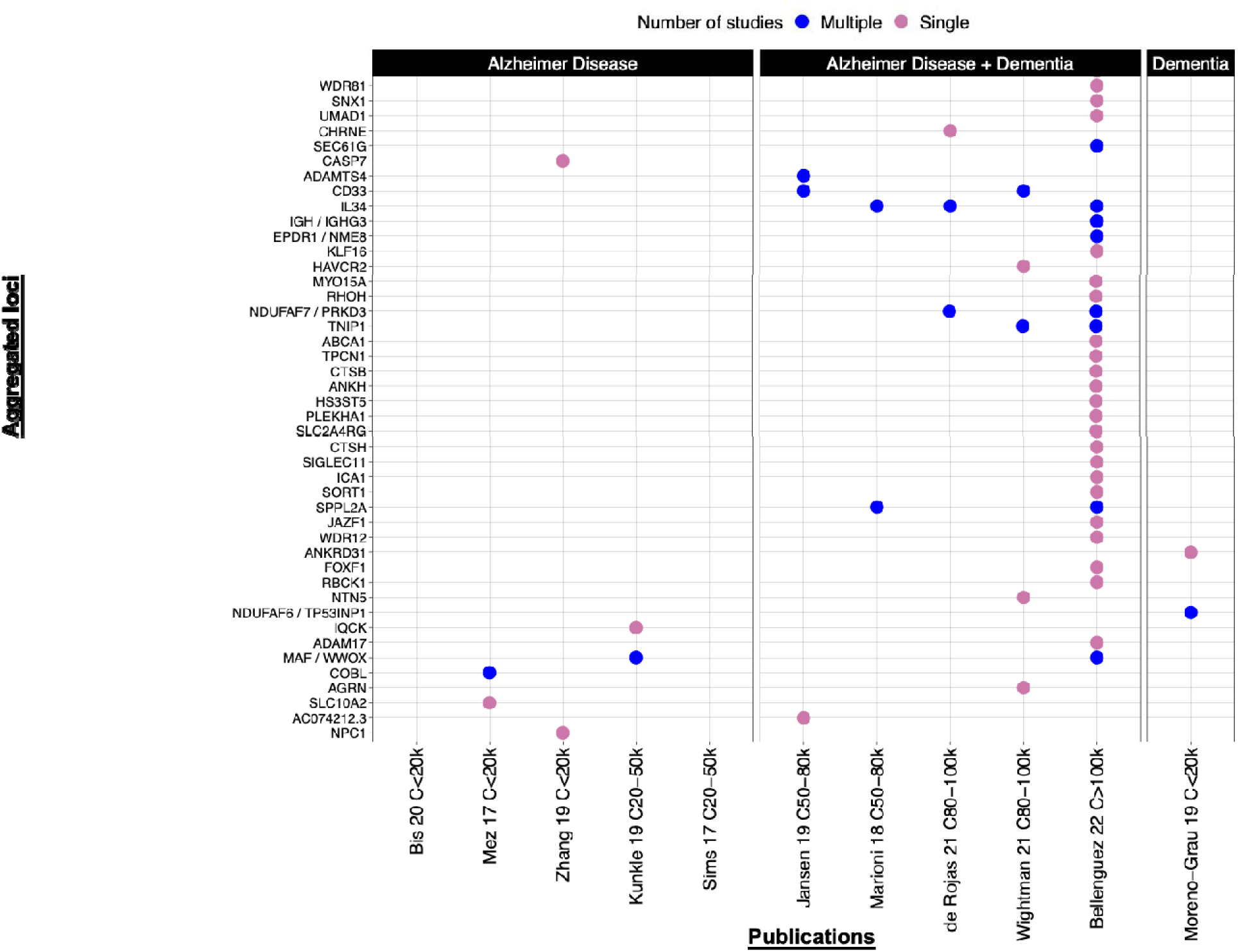
Phase 2 single-variant (SV), high-confidence loci (Tiers 1 or 2) for AD, AD+Dementia, and Dementia phenotypes reported in one or more publication. Variants (loci) that are physically close were grouped by linkage disequilibrium (LD) pruning using FUMA^42^. Names of the loci reflect the closest gene to the sentinel single nucleotide variant (SNV). Data are from NHW-EA samples. Loci are sorted by association p-value with the lowest p-values at the top and largest p-values at the bottom (and from left to right). The publications are on the X-axis with the case sample size bin for each study indicated. Even though *APOE* generates by far the smallest p-value, only 2 studies reported results for this gene. Details about the genes or gene groups on the Y-axis (n = 88) are listed in **Supplementary Table 1**. Red dot: loci reported in only one publication. Blue dot: loci reported in multiple studies. Only loci with Tier rankings of 1 and 2 are displayed. *SEC61G* (row 5 in second half of the figure) is a Tier 1 locus in Bellenguez 2022^37^ but a Tier 3 locus in He 2021^34^.

**Table 4.** Summary of Phase 2 loci across phenotypes and ancestry groups by single-variant (SV) or variant-set (VS) analysis.

| Analysis | Tiers and phenotypes | NHW-EA | Others including Multi-ancestry |
| --- | --- | --- | --- |
| SV | T1–4 AD loci | 39 | 15 |
|  | T1–2 high confidence AD loci | 29 | 7 |
|  | T1–4 AD+Dementia loci | 88 | 0 |
|  | T1–2 high-confidence AD+Dementia loci | 83 | 0 |
| VS | T1–4 AD loci | 23 | 13 |
|  | T1–2 high confidence AD loci | 14 | 11 |
|  | T1–4 dementia loci | 3 | 0 |
|  | T1–2 high confidence dementia loci | 0 | 0 |

Significant associations were identified with nine loci or gene clusters in both SV and VS analyses (**Table 5**). These results suggest that more than one non-codling regulatory region is influencing risk for a specific locus or that a single regulatory element or regulatory elements in proximity are influencing more than one gene.

**Table 5.** High-confidence AD loci (Tiers 1–4) that were identified in both single-variant (SV) and variant-set (VS) association tests. The genes listed in the loci column were what the publication reported. Details for the SV and VS association tests are in **Supplementary Tables 1 and 2**. The SNV IDs are those used in the SV analysis.

| Loci | Ancestry group | Analyses | SNV IDs<sup>c</sup> |
| --- | --- | --- | --- |
| *ABCA7* | NHW-EA | SV<sup>20,24,33,37</sup> | rs12151021<sup>c</sup>, rs3752246, rs111278892 |
|  | Multi-ancestry<sup>a</sup> | VS<sup>27</sup> | 43 SNVs |
| *CR1* | NHW-EA | SV<sup>20,24,33,37</sup> | rs679515<sup>c</sup>, rs4844610, rs2093760 |
|  | NHW-EA | VS<sup>14</sup> | Not reported |
| *APOE / TOMM40* | Multi-ancestry<sup>b</sup> | SV<sup>9,10,24,33</sup> | rs41289512, rs157591, rs394819, rs429358 |
|  | NHW-EA | VS<sup>21</sup> | 70 SNVs |
| *BIN1* | NHW-EA | SV<sup>20,24,33,37</sup> | rs6733839<sup>c</sup>, rs4663105<sup>c</sup> |
|  | NHW-EA | VS<sup>11,14</sup> | Not reported |
| *CLU / PTK2B* | NHW-EA | SV<sup>20,24,33,37</sup> | rs11787077, rs4236673, rs9331896, rs1532278 |
|  | NHW-EA | VS<sup>14</sup> | Not reported |
| *EED / PICALM* | NHW-EA | SV<sup>20,24,33,37</sup> | rs3851179, rs867611, rs3851179, rs561655 |
|  | NHW-EA | VS<sup>14</sup> | Not reported |
| *HLA* | NHW-EA | SV<sup>20,24,33,37</sup> | rs6605556, rs6931277, rs9271058, rs1846190 |
|  | NHW-EA | VS<sup>11</sup> | Not reported |
| *MS4A / MS4A2 / MS4A4A / MS4A6A* | NHW-EA | SV<sup>20,24,33,37</sup> | rs1582763<sup>c</sup>, rs7232, rs2081545, rs7933202 |
|  | NHW-EA | VS<sup>14</sup> | Not reported |
| *OARD1 / TREM2 / TREML2 / UNC5CL* | NHW-EA | SV<sup>18-20,27,33,37</sup> | rs75932628<sup>c</sup>, rs9381040, rs187370608 |
|  | Multi-ancestry<sup>b</sup> | VS<sup>22,27</sup> | 33 SNVs, Not reported |

<sup>a</sup> Multi-ancestry groups in this study include NHW-EU, African, and Asian subjects.
<sup>b</sup> Multi-ancestry groups in this study include NHW-EU, African, and Hispanic/Latino subjects.
<sup>c</sup> Variants tested in more than one publication in VS analyses.

Distinct and high-confidence (Tier 1 or 2) loci for AD, AD+Dementia, and Dementia were grouped across studies using FUMA^42^ (**Figure 2)**. Note that the largest studies specified AD+dementia as the outcome^43^ and identified the largest numbers of tier 1 and 2 loci. To generate a complete locus/gene list, we combined high-confidence signals from Phase 1, Phase 2, and variant-set studies. This yielded 111 high confidence loci/genes for AD/dementia across all ancestry groups examined (**Tables 1, 3, and Supplementary Table 1)**.

For many of the loci detected, as the sample size increased, the effect size remained robust, p-value became smaller (more significant), and the confidence interval (CI) for the odds ratio (OR) was narrowed. For example, the initial publication showing genome-wide significant association of *BIN1* with AD reported a P-value of 1.59 × 10^−11^ and OR of 1.13 with a 95% CI of 1.06-1.21 using 8,371 cases, and 26,965 controls (**Table 6**)^44^. In a later, larger study that included 30,344 cases and 52,427 controls, the significance of the association increased and the CI narrowed (P= 2.1 × 10^−44^, OR=1.2, 95% CI = 1.3) ^20^. A much larger study of AD+Dementia^37^ including 719,803 controls and 111,326 cases reported an even more significant association (P=6.10 × 10^−118^) with a similar effect size (OR=1.17) and narrower CI (95% CI 1.16-1.19)^37^. Similar trends were observed for *CLU*^44,45^]. In contrast, association with *CD33*, a gene that has received attention as a potential drug target for AD[46], was first reported in a study^47^ of 1,376 subjects from 410 families, noting that the result was not genome-wide significant (P=4 × 10^−6^ for SNP rs3826656). A subsequent case-control study by Naj et al ^48^ including 11,840 cases and, 10,931 controls reported a more significant association with *CD33* SNP rs3865444 (P=1.1 × 10^−7^, OR=0.88, 95% CI 0.84-0.93). A genome-wide significant association with rs3865444 was observed in a more recent larger study of 24,087 cases and 55,058 controls (P=4.3 × 10^−8^)^24^. However, evidence for association with this variant was greatly reduced in two much larger GWAS for AD/dementia, one that included 71,880 cases and 383,548 controls (P=6.3 × 10^−9^) and the other including 383,548 cases and 719,803 controls (P=3.1 × 10^−4^) ^24,37^. Finally, a very recent genome-wide consensus meta-analysis study from Bellenguez et al ^49^ that included 128,681 AD cases from Bellenguez et al ^49^ that included 128,681 AD cases from Bellenguez et al[37], the ADGC, DemGene, EADB, EADI, GERAD, GR@ACE/DEGESCO, IGAP, and PGC-AL resulted in the most significant result for rs3865444 to date (P=4.59 × 10^−13^ OR=0.95 (0.94-0.96)]. Compared to their previous report, the most recent Bellenguez et al study contained a much larger sample of AD cases, including the addition of participants from the Nordic countries of Iceland, Sweden, and Norway (**Supplementary Table 3**). Further examination of these results may reveal whether this major boost in significance is due to a higher frequency of the rs3865444 effect allele or a biologically causal variant in LD in the Nordic samples. Alternatively, the Nordic samples may be enriched for an AD subtype that is more strongly associated with causal variants in *CD33*. Of note, we tested but did not find LD between *APOE* and *CD33* markers that are separated by 6.32 Mb on chromosome 19 (**Supplementary Figure S1)**.

**Table 6.** Comparison of association signals across publications for selected loci: *BIN1*, *CD33,* and *CLU*.

|  |  |  |  |  |  |  |  |  |  |  |
| --- | --- | --- | --- | --- | --- | --- | --- | --- | --- | --- |
| ***BIN1*** | **Publication** | Seshadri 2010<sup>43</sup> | Jansen 2019<sup>a</sup><sub>24</sub> | Kunkle 2019<sup>20</sup> | Jansen 2019<sup>a, 24</sup> | Wightman 2021<sup>33</sup> | Bellenguez 2022<sup>37</sup> |  |  |  |
|  | **Sample size – Total** | 35,336 | 79,145 | 82,771 | 455,258 | 1,126,563 | 831,129 |  |  |  |
|  | **Sample size – Cases** | 8,371 | 24,087 | 30,344 | 71,880 | 90,338 | 111,326 |  |  |  |
|  | **Phenotypes** | AD | AD | AD | AD+Dementia | AD+Dementia | AD+Dementia |  |  |  |
|  | **OR or Z score** | OR=1.13, 95% CI 1.06-1.21 | NA | OR=1.18; 95% CI 1.15-1.22 | Z = 13.94 | NA | OR=1.17 95% CI 1.16-1.19 |  |  |  |
|  | **Location (hg19)** | 2:127894615 | 2:127891427 | 2:127892810 | 2:127891427 | 2:127891427 | 2:127892810 |  |  |  |
|  | **SNV** | rs744373 | rs4663105 | rs6733839 | rs4663105 | rs4663105 | rs6733839 |  |  |  |
|  | **P-value** | 1.59 x 10<sup>-11</sup> | 3.58 x 10<sup>-29</sup> | 2.10 x 10<sup>-44</sup> | 3.38 x 10<sup>-44</sup> | 3.92 x 10<sup>-58</sup> | 6.10 x 10<sup>-118</sup> |  |  |  |
| ***CD33*** | **Publication** | Bertram 2008<sup>b47</sup> | Katsumata 2020<sup>28</sup> | Naj 2011<sup>*48</sup> | Hollingsworth 2011<sup>*39</sup> | Lambert 2013<sup>* 80</sup> | Jansen 2019<sup>24*a</sup> | Jansen 2019<sup>a24</sup> | Bellenguez 2022<sup>37</sup> | Bellenguez 2025 (unpublished) |
|  | **Sample size – Total** | 1376 (410 families) | 11,330 | 22,771 | 59,716 | 74,076 | 79,145 | 455,528 | 831,129 | 978,514 |
|  | **Sample size – Cases** | NA | 5,142 | 11,840 | 19,870 | 25,580b | 24,087 | 71,880 | 111,326 |  |
|  | **Phenotypes** | AD | AD | AD | AD | AD | AD | AD+Dementia | AD+Dementia | AD+Dementia |
|  | **OR or Z score** | NA | NA | OR=0.88; 95% CI 0.84-0.93 | OR=0.91; 95% CI 0.88-0.93 | OR=0.94<sup>**</sup>, I2 (%) 0 | Z = -5.48 | Z = -5.81 | OR=0.97 | OR=0.95<sup>**</sup> |
|  | **Location (hg19)** | 19:51726613 | 19:5128477 | 19:51727962 | 19:51727962 | 19:51727962 | 19:51727962 | 19:51727962 | 19:51727962 | 19:51727962 |
|  | **SNV** | rs3826656 | rs12459419 | rs3865444 | rs3865444 | rs3865444 | rs3865444 | rs3865444 | rs3865444 | rs3865444 |
| | **P-value** | $4.00 \times 10^{-6}$ | $4.00 \times 10^{-4}$ | $1.10 \times 10^{-7}$ | $1.60 \times 10^{-9}$ | $3.0 \times 10^{-6}$ | $4.30 \times 10^{-8}$ | $6.30 \times 10^{-9}$ | $3.10 \times 10^{-4}$ | $4.59 \times 10^{-13}$ |
| **CLU** | **Publication** | Harold 2009<sup>45</sup> | Jansen 2019<sup>24</sup> | Kunkle 2019<sup>20</sup> | Schwartzentruber 2021<sup>32</sup> | Jansen 2019<sup>24</sup> | He 2021<sup>34</sup> | Wightman 2021<sup>33</sup> | Bellenguez 2022<sup>37</sup> |  |
|  | **Sample size - Total** | 16,152 | 79,145 | 82,771 | 408,942 | 455,258 | 488,377 | 1,126,563 | 831,129 |  |
|  | **Sample size - Cases** | 5,964 | 24,087 | 30,344 | 53,042 | 71,880 | NA | 90,338 | 111,326 |  |
|  | **Phenotypes** | AD | AD | AD | AD+Dementia | AD+Dementia | AD+Dementia | AD+Dementia | AD+Dementia |  |
|  | **OR or Z score** | OR=0.86 95% CI (0.82-0.90) | NA | OR=0.88 95% CI 0.85 – 0.91 | OR=1.11<sup>**</sup> | Z = -8.98 | NA | NA | OR=0.91<sup>**</sup> |  |
|  | **Location (hg19)** | 8:27464519 | 8:27464929 | 8:27467686 | 8:27468503 | 8:27464929 | 8:27465264 | 8:27466315 | 8:27465312 |  |
|  | **SNV** | rs11136000 | rs4236673 | rs9331896 | rs867230 | rs4236673 | rs35500730 | rs1532278 | rs11787077 |  |
| | **P-value** | $8.5 \times 10^{-10}$ | $6.36 \times 10^{-20}$ | $4.60 \times 10^{-24}$ | $7.71 \times 10^{-26}$ | $2.61 \times 10^{-19}$ | $4.40 \times 10^{-10}$ | $1.57 \times 10^{-22}$ | $1.70 \times 10^{-44}$ | |

\* Publications were not part of the Phase 2 adjudication effort.
\*\* Confidence interval not reported.
<sup>a</sup> The Jansen paper<sup>24</sup> reported two *CD33* results: one using 24,087 AD cases and 48,496 controls and one using 71,880 AD+Dementia cases and 382,648 controls. GVC used the latter with a larger sample size for the tier ranking for *CD33*.
<sup>b</sup> Family study; first report implicating *CD33*. Reported P-value ( $4.00 \times 10^{-6}$ ) is not directly comparable to others listed in the table and a genome-wide significance cutoff used for this SNP, $4.88 \times 10^{-6}$ , which is greater than the genome-wide significance level of $5 \times 10^{-8}$ used for other studies.

### Candidate genes associated with GVC loci

GWAS signals are typically annotated by the closest gene. However, most common variants identified by GWAS are non-coding and are thought to influence disease risk by modulating the activity of gene regulatory elements, such as enhancers, rather than by directly altering protein-coding sequences. These regulatory variants can act in cis to affect the expression of one or more causal genes, which may be located some distance away from the associated SNVs. Indeed, several studies using LD score regression and epigenomic annotations across diverse cell types have shown that AD risk SNVs are specifically enriched in enhancers active in myeloid cells, including microglia^50^. Thus, to capture potential candidate causal genes mediating AD risk associations at GVC loci, we generated lists of protein coding genes that were either ±150 kb or ±500 kb from all reported SNVs from any Tier 1-2 NHW-EA loci (Phase 2 SV studies) using ENSEMBL version 109 to annotate genes. Loci identified by VS analysis studies were also included. Of the 88 GVC high-confidence loci considered, 5 did not have a coding gene in these intervals and thus were not considered for further evaluation. For the ±150kb and ±500kb bins, there were 501 and 1,344 genes, respectively, and 508 and 1,346 genes, respectively, if including the Phase 1 genes (**Supplementary Table 4**).

To refine the connection between the associated SNV and the causative AD-related gene(s), we examined the topologically associating domain (TAD) structure for the loci in **Tables 1** and **3**. Non-coding SNVs are potentially in cis-acting regulatory elements (CREs) that influence the expression of neighboring genes by interacting with the target gene promoter. The target gene might not always be the closest gene but is typically within the same TAD as the associated SNV. Typically, in 60–70% of SNP associations, the closest gene is the true target, and in 10–15%, the target gene is the second closest gene. We used brain-derived TADs from hippocampus and dorsolateral prefrontal cortex (DLPFC) as defined by Hi-C experiments^50^. The majority of variants and their corresponding candidate genes were located within the same TAD (Category 1), with a higher proportion observed for the hippocampus-derived TADs (84%) compared with DLPFC-derived TADS (74%). Fewer than 5% of variants had their associated SNV locus and the candidate gene in a different TAD (Category 2) (**Supplementary Table 5a**). For *APP/ADAMST1/LINC00158*, the critical common variant (rs2830489, 1000genomes minor allele frequency for EUR is 0.2962) is not in the same TAD as the *APP* gene or promoter. This suggests that the common variant signal may be regulating *ADAMTS1* and not *APP*. However, rare variant analysis unambiguously identifies *APP* as a causal AD gene. There are three other variant-based Tier 1 loci (rs112404845 for *COBL*, rs7068231 for *ANK3*, and rs72824905 for *PLCG2*) where the promoter region of the reported candidate gene is not within the same TAD as the most significant variant in the association analysis (**Supplementary Table 5b**). This suggests that these AD- associated SNVs might influence disease mechanisms by regulating genes close to the proposed candidate gene.

We compared the GVC prioritized gene lists to the candidate list from Agora (**Figures 3A and B**)^51^. The Agora list contains genes nominated by investigators based on genomic and proteomic and/or metabolomic data from human brain samples. For the Agora list, we compared the Target Risk Scores, the Genetic Risk Scores, and the Multi-omic Risk Scores for each gene in the GVC ±150kb and ±500kb lists (**Supplementary Table 4**). The Target Risk score is the sum of the Genetic Risk score (**Supplementary Figure S1B**) and the Multi-omic Risk score. Scores are provided in columns S–U in **Supplementary Table 6**. Even though the GVC list is expected to contain false-positive genes since it includes all genes in a region flanking a true causative gene or genes, the GVC list yielded significantly higher Genetic Risk Scores compared with the Agora list (**Supplementary Figure S2A**). The 500 kb and 150 kb lists include 120 (8.8%) and 72 (7.6%) genes overlapping the Agora list of 947 genes, respectively (using the GVC set as the denominator). Of the 12 high-confidence GVC genes (**Table 1**), 4 GVC genes are not in the Agora list (*CASP7, PSEN1, PSEN2, and SORL1*) (**Supplementary Figure S3, Supplementary Table 4**). Overall, 43 GVC loci bins do not contain an Agora gene, and 3 Agora genes are not in a GVC bin (**Supplementary Figure S3**). Thus, the GVC identifies valid loci missed by Agora.

**Figure 3.**
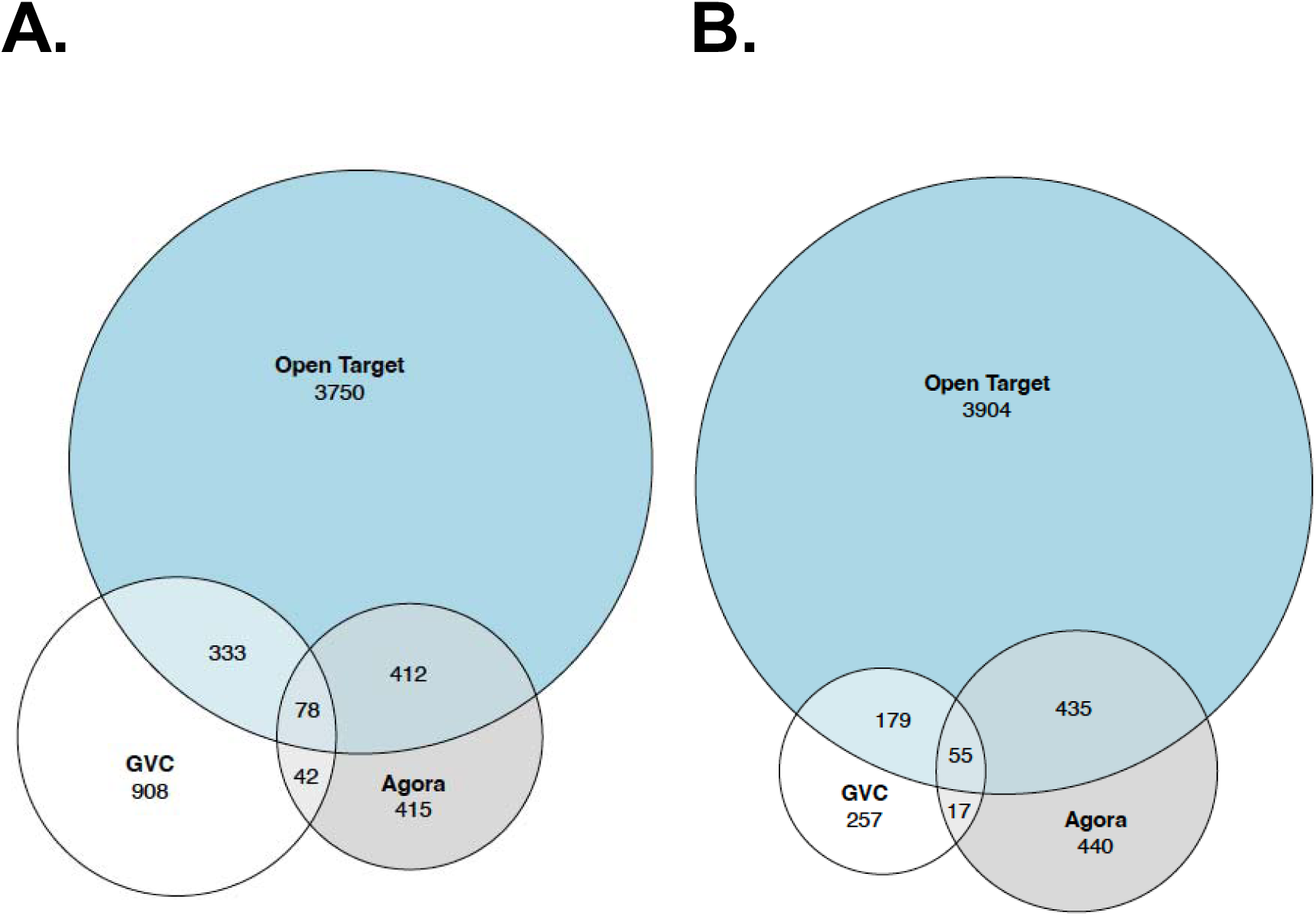
For the GVC, the number of coding genes ±500 kb from all SNVs within high-confidence loci (**Supplementary Table 1**, Tiers 1 and 2), additional Tier 1 and 2 loci from variant-set analyses (**Supplementary Table 2**), and Phase 1 loci (Tiers 1 and 2)(**Table 4)**. This yielded 1,361 GVC genes (**Supplementary Table 3**). The Agora gene list of 947 genes is from https://agora.adknowledgeportal.org/genes/nominated-targets (downloaded on 03/21/24). The Open Targets gene list of 4,573 AD genes is from https://platform.opentargets.org/disease/MONDO_0004975/associations (v24.03) (download on 06/04/24). Figure 3B. GVC genes that are ±150 kb from the most significant SNV from the same sources as Figure 3A yielded 508 genes. Agora and Open Targets gene lists remain the same. Note: Coding proteins are from ENSEMBL.

We also reviewed the results from the Open Targets platform, which generates its scores (0: weakest evidence, 1: strongest evidence) by integrating large-scale GWAS data, gene expression and protein abundance data, chromatin interaction and conformation data, and a variety of other functional genomics datasets from many cell types and tissues^52^. Note that strong AGORA scores do not always correlate with strong Open Targets scores because AGORA uses AD–specific multi-omic and genetic risk data, while Open Targets integrates broader, multi-disease GWAS and functional genomics evidence with different scoring methods.

In addition, we reviewed other work where candidate genes were nominated for GWAS signals ^20,32,37,53–55^. Evidence evaluated included presence of coding or splicing variants, expression in tissue relevant to AD, transcriptomics data, epigenomic annotations, eQTL data including co-localization with AD, pathway analysis, chromatin accessibility, three-dimensional chromatin interactions, Braak stage association, and differential expression in AD versus normal brains^20^. Some work focused primarily on myeloid and microglial cells ^53,55,56^. Focusing on novel mechanisms, the functional genomics studies we evaluated here excluded *APOE*, due to its well-established genetic and functional contributions to AD biology. **Supplementary Table S6** shows the candidate genes nominated by these studies in the GVC ±500 kb gene list. Of the list of 75 overlapping genes reported by GVC, Agora, and Open Targets (column K in **Supplementary Table 6**), 35 genes across 25 GVC loci were prioritized as candidate AD genes by at least one publication reviewed (**Table 7** and column X in **Supplementary Table 6**). Of these 35 genes, 21 were prioritized by more than one study (*ABCA7, ACE, ADAM10, CCDC6, BIN1, CASS4, CD33, CLU, PTK2B, CR1, CTSH, USP6NL, PICALM, EPHA1, HLA-DRA, INPP5D, SPI1, MS4A4A)*.

**Table 7.** 35 genes across 25 GVC loci reported by GVC, Agora, and Open Targets were prioritized as candidate AD genes using functional data by at least one publication reviewed by the GVC.

| **GVC Locus** | **GVC expanded list of possible genes (500kb)** | **Is there evidence for rare risk, protective, or causal variants for AD within the gene? And what is the strength of evidence?\***<sup>57</sup> | **N of Studies Prioritizing Gene** |
| --- | --- | --- | --- |
| *ABCA7* | *ABCA7* | Yes, replicated risk gene<sup>81-84</sup> | 4 |
| *ABI3* | *ZNF652* |  | 1 |
| *ACE* | *ACE* | Yes, replicated risk gene<sup>85,86</sup> | 2 |
| *ADAM10 / MINDY2* | *ADAM10* | Yes, risk gene needing replication<sup>87</sup> | 2 |
|  | *LIPC* |  | 1 |
| *ADAMTS4* | *ADAMTS4* |  | 1 |
|  | *FCER1G* |  | 1 |
|  | *NDUFS2* |  | 1 |
| *ANK3 / CCDC6* | *CCDC6* |  | 2 |
| *BIN1* | *BIN1* | Yes, replicated risk gene<sup>88,89</sup> | 5 |
| *CASS4* | *CASS4* |  | 5 |
| *CD2AP* | *CD2AP* | Yes, risk gene needing replication<sup>88</sup> | 1 |
| *CD33* | *CD33* |  | 4 |
| *CHRNE* | *RABEP1* |  | 1 |
| *CLU / PTK2B* | *CLU* | Yes, replicated risk gene<sup>90</sup> | 4 |
|  | *PTK2B* |  | 3 |
|  | *SCARA3* |  | 1 |
| *CR1* | *CR1* | Yes, risk gene needing replication<sup>91</sup> | 3 |
| *CTSH* | *CTSH* |  | 2 |
| *DOC2A* | *DOC2A* |  | 1 |
| *ECHDC3 / USP6NL* | *USP6NL* |  | 4 |
| *EED / PICALM* | *PICALM* |  | 5 |
| *EPHA1 / EPHA1-AS1* | *EPHA1* | Yes, risk gene needing replication<sup>88</sup> | 3 |
| *HLA / HLA-DRB5* | *HLA-DRA* |  | 2 |
|  | *HLA-DRB1* |  | 1 |
| *INPP5D* | *INPP5D* |  | 4 |
| MADD / SPI | C1QTNF4 |  | 1 |
|  | NDUFS3 |  | 1 |
|  | NR1H3 |  | 1 |
|  | SPI1 |  | 3 |
| MS4A / MS4A2 / MS4A4A / MS4A6A | MS4A4A |  | 3 |
|  | MS4A6A |  | 4 |
| OARD1 / TREM2 / TREML2 / UNC5CL | TREM2 | Yes, replicated risk gene<sup>92,93</sup> | 3 |
| PLCG2 | PLCG2 | Yes, replicated risk gene<sup>18,94,95</sup> | 1 |
| RIN3 / SLC24A4 | RIN3 | Yes, risk gene needing replication (28738127)<sup>91,96</sup> | 4 |

\*Categories adapted from Khani et al.<sup>57</sup> Variants identified in Khani et al.<sup>57</sup> and through expert curation.

**Table 7** also shows whether a GVC locus harbors a rare variant that has been identified as either being causal or risk increasing for development of AD. Three categories are represented, as defined by Khani et al. ^57^, and rare variants identified within these categories by this publication are noted with their PMID in the column. The three categories are: 1) genes harboring rare variants that cause AD, 2) replicated genes harboring rare variants with reduced penetrance and contributing to AD risk, and 3) genes harboring rare variants with an initial association to AD. Genes in the first two categories are likely important to development of AD, while genes in the last category require independent replication before our confidence in their involvement is increased. In all, three causal genes (*APP, PSEN1, PSEN2*), eight replicated risk genes (*ABCA7, ABI3, BIN1, CLU, NCK2, TREM2, PLCG2, SORL1*) and 17 additional genes with an initial rare variant association to AD have been identified within GVC loci.

### Pathway analysis of candidate genes

To understand the mechanism(s) underlying AD and all-cause dementia, we performed pathway analyses using the three gene sets described above (**Figure 4**; GVC, Agora, and Open Targets). The GVC set included all genes in bins ±500 kb from the top GWAS SNV. Pathways related to APP processing, cellular response to amyloid-beta, immune response, efferocytosis, and cholesterol efflux/lipid transport are detected across all three gene sets.

**Figure 4.**
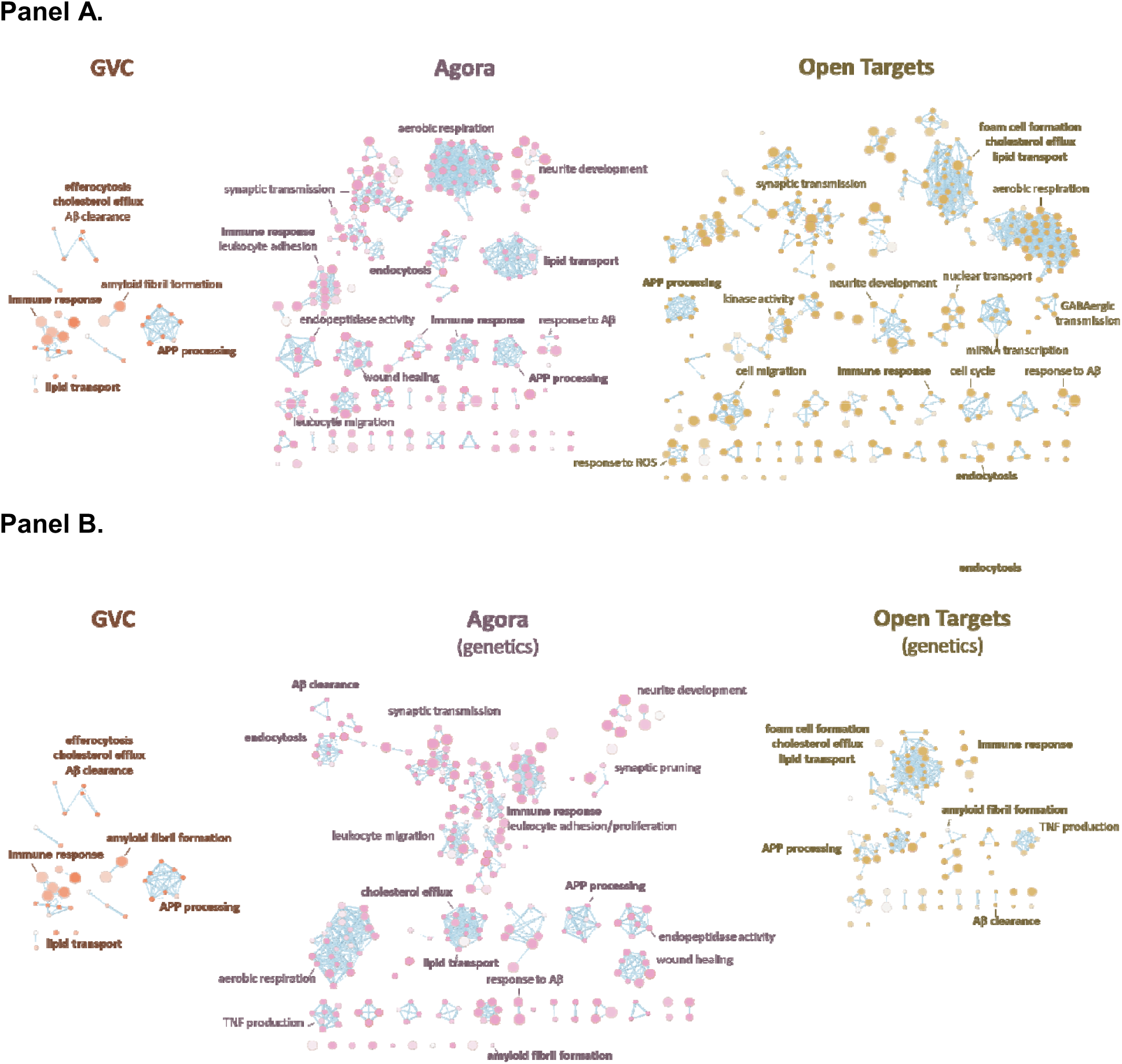
EnrichmentMap visualizations^97^ of pathway enrichment results for candidate AD risk gene sets from GVC, Agora (sorted by target risk score or genetics score), and Open Targets (filtered and sorted by Open Targets score or Open Targets Genetics score). Pathways that share many genes cluster together into functional modules, which are labeled with representative biological processes such as “immune response,” “lipid transport,” or “APP processing.” The same networks with GO Biological Process labels for each node and the corresponding enrichment statistics are included in Supplementary Tables 7–11. In each network, nodes represent significantly enriched pathways, and edges between nodes reflect the degree of gene overlap between those pathways. The size of each node is proportional to the number of genes in the corresponding pathway, and thickness of connecting edges indicates the extent of shared genes (i.e., thicker edges denote higher gene overlap). The color of each node reflects the statistical significance of the enrichment, with darker nodes representing lower adjusted p-values. Panel A displays all genes in each list; Panel B is filtered for genetic risk.

The Agora and Open Targets initiatives incorporate substantial transcriptomic and functional genomics data on top of genetic data. Therefore, their pathway analyses using genes with high multi-omics scores identify processes additional to core AD pathways. These include neuronal and synaptic function as well as energy metabolism (e.g., synaptic transmission, aerobic respiration, neurite development, GABAergic transmission, and cellular stress responses). Yet, when we used genes with high genetic scores within the Agora and Open Targets portal, the resulting pathways are more comparable with that derived from the GVC list. Specifically, pathways involving Aβ clearance, cholesterol efflux, APP processing, immune response, lipid transport, and amyloid fibril formation are emphasized, closely mirroring the GVC findings. We summarized these results using EnrichmentMap[58] (**Figure 4)**, displaying pathway enrichment results on genes from GVC, genes with high genetic scores in Agora, and genes with high Open Targets Genetics Score in Open Targets.

## Discussion

### Development of criteria to evaluate and rank the strength of evidence of genetic signals for complex disease

The GVC developed criteria for evaluating the strength of evidence supporting the association of a genetic signal with AD/ADRD. Multiple lines of evidence were considered. **First**, for common variants, the signal from the sentinel variant must be supported by other correlated variants. Unsupported signals are typically artifacts arising from faulty array assays or chance that is most commonly observed when the effect allele frequency difference between cases and controls is small. **Second**, potential population substructure differences between cases and controls must be examined and accounted for to avoid artefactual associations. Similarly, analysis must adjust for differences in ancestry, especially when multiple cohorts are combined, **Third**, diagnostic criteria and methods should be consistent across subjects. **Fourth**, replication requires an independent sample of similar or larger size however, some experts advocate combining all available cohorts into a single discovery sample to maximize statistical power^58^. Because nearly all large published ADRD GWAS include data from previous studies and augment that with data from additional individuals, increasing the significance and observing consistent effect direction is often considered as replication.

Importantly, these criteria could be applied to the evaluation of the strength of genetic evidence for associated loci of other complex disease traits. While strategies for systematic assessment of evidence linking genes to complex disease etiology have been used^59–62^, guidelines for evaluation of support for genetic variants and genes in disease have largely focused on assessment of variants implicated in the etiology of Mendelian disorders. For instance, in 2015 an expert working group of the American College of Medical Genetics and Genomics (ACMB) and the Association for Molecular Pathology (AMP) developed consensus recommendations for use of standard terminology to describe the likely pathogenicity of genetic variants (“pathogenic,” “likely pathogenic,” “uncertain significance,” “likely benign,” and “benign”) implicated in Mendelian disorders^63^. These classifications are based on the evaluation of several types of variant evidence (e.g., population data, computational data, functional data, segregation data). This scheme for assessment of variant pathogenicity was incorporated into a classification framework developed by the NIH-funded Clinical Genome Resource (ClinGen), where genetic and experimental evidence for support of gene-disease relationships are evaluated and classified as ““Definitive,” “Strong,” “Moderate,” “Limited,” “No Reported Evidence,” or “Conflicting Evidence^64^. These evaluations, which are reviewed and confirmed or adjusted by disease experts periodically, are made available via the database ClinVar^65^. A few recently developed classification schemes have incorporated and extended this framework to assess genetic variants related to complex disorders^66,67^ including a framework for assessing genetic evidence related to autism spectrum disorder which recognized the importance of assessment of the quality of phenotype. This scheme begins with a list of strong genetic candidates for assessment based on number of published studies and number of individuals reported with relevant variants in the gene. The ClinGen framework is then used to generate a preliminary score for variants, and then level of confidence is determined based on a phenotype quality assessment which is then used to adjust the preliminary ClinGen score.

Although guidelines exist for evaluating the strength of support for sequence variants implicated via clinical testing or sequence analyses of Mendelian and a few complex disorders, comprehensive recommendations on how to evaluate and rank genetic signals (e.g., GWAS loci) from complex disease association studies have not been published. Several methods for evaluating certain aspects of GWAS loci association quality or strength have been published, however. These include a recent method, the GWAS quality score (GQS), which aims to identify suspicious associated loci via a quantitative and automated analysis of summary statistics^68^. This and other similar methods require raw genotype or summary statistics data^69^ for evaluation of association quality. Dombos et al. proposed human genetic evidence (HuGE) guidelines for summarizing the genetic support for a gene in a human disease which combines evidence from GWAS and WES associations^59^. The score combines summary statistics of both common variant and rare variant association results to rank genetic support for a gene in a six-tier system of “no evidence,” “anecdotal,” “moderate,” “strong,” “very strong,” and “compelling.” While we recognize the value of incorporating automated quantitative evaluation of summary statistics in evaluation of an associated locus’s quality, our proposed framework relies on expert evaluation of a studies quality, in addition to a locus’s strength of association to rank a locus’s evidence for involvement in disease. Furthermore, it does not require summary statistics, which may not be available for all published studies targeted for evaluation, though we strongly support efforts to require their availability upon publication. Indeed, for meta-analyses it is ideal to have summary statistics available for each cohort used within the study to evaluate evidence of heterogeneity in the strength of support for individual loci across cohorts.

### Clinical data without biomarker support also has a significant misdiagnosis rate

One of the challenges in interpreting the robustness and specificity of genetic association findings is the variability within and across studies of the case definition. An autopsy diagnosis is the gold standard because it can distinguish between AD and other causes of dementia. Unfortunately, the number of subjects with autopsy is limited. Furthermore, (over 70% of autopsied AD cases present with mixed pathology, particularly in older individuals, making interpretation challenging. PET imaging using compounds that bind Aβ or tau or cerebrospinal fluid measures of Aß and tau can enhance diagnostic accuracy in living individuals, but these are expensive and invasive, reducing their utility in large-scale genetic studies. The recent development of blood-based biomarkers for AD and other dementias holds great promise as an inexpensive non-invasive way of improving diagnostic accuracy. Currently only a minority of cohorts have biomarker data, and harmonization of these data, particularly those that are a part of the ADSP, by the phenotype Harmonization Consortium (PHC) is in progress. Despite these efforts, there is a trade-off between diagnostic accuracy/specificity and sample size, noting that the largest AD GWAS include subjects with a rigorous clinical diagnosis of AD, a self-report or electronic medical record indication of AD or unspecified dementia, and proxy AD cases. A clinical diagnosis can differ from post-mortem pathology in up to 30% of subjects. Disorders that clinically overlap with AD include some forms of FTD, hippocampal sclerosis (HS), Lewy body dementia (LBD), Parkinson disease with dementia, vascular dementia, and other rarer causes of dementia. Since AD is the most common cause of dementia, contamination by clinically misdiagnosed cases is most likely a bigger problem in GWAS of rarer ADRD.

### Co-morbidity versus common mechanisms

Several genetic loci associated with AD/ADRD were previously implicated in other neurodegenerative diseases such as progranulin in which high penetrance mutations are associated with risk for frontotemporal dementia (FTD). Similarly, several AD GWAS loci are linked to other neurodegenerative diseases. Variants in the *MAPT/KANSL1* locus increase risk of AD, PD, PSP, CBD, and ALS, while *SETD1A*/*KAT8*, *LCORL*, *WWOX*, *GRN,* and *CLU* variants are associated with both AD and PD. *GPX3, HS3ST5/HDAC2/MARCKS, and TSPOAP1* perturb both ALS and AD related pathways. Colocalization analysis supports the same causal variants in *CLU, WWOX and LCORL* underlying AD and PD risk and in *TSOAP1* for AD and ALS risk. Despite limited GWAS sample sizes in Lewy Body Dementia, shared genetic risk factors include BIN1 and APOE with AD, and SNCA and GBA with PD. While these genes could perturb common neurodegenerative pathways, it is also possible that study participants diagnosed with clinical AD might have been misdiagnosed or have concomitant neurodegenerative pathologies. This is exacerbated particularly in studies such as the UK Biobank cohort, where AD is not clinically diagnosed, but is assigned based on a report of dementia in a parent.

A study of over 1000 participants clinically diagnosed with AD found that only 21% had pathology limited to AD^70,71^. AD clinical diagnosis was associated with LBD pathology in another 33% and CVD pathology in 16.5%, and nearly 7% were reported to have hippocampal sclerosis (HS), possibly reflecting TDP-43 related pathology^70^. In the Religious Orders Study and the Rush Memory and Aging Project (ROS/MAP), 21% of individuals with dementia had pathology limited to CVD, but 56% of the 1,767 individuals had mixed pathologies including TDP-43 deposits, LBD and HS^71^. Mixed pathologies are found more often among older individuals and in community-based studies^72^.

Although GRN variants are strongly associated with FTD, several studies identified genome-wide significant association of variants in this gene with AD^73,74^, but they are not likely to be causal. Thus, it is possible that progranulin (the protein encoded by GRN) has a role in AD, FTLD, and other neurodegenerative diseases through convergent pathways such as lysosomal storage in neurons and microglia. GRN variants might also account for concomitant tauopathies or other manifestations in AD neuropathology.

### Genetic versus omics approaches to gene discovery

Although core AD pathways are conserved in GVC gene lists, the Agora and Open Targets initiatives incorporate genetic data with substantial transcriptomic and functional genomics data, notably including analysis results from transcriptomic data obtained from post-mortem AD brain tissue samples. Consequently, these pathway analyses identify additional processes involving neuronal and synaptic function, as well as energy metabolism—specifically highlighting pathways such as synaptic transmission, aerobic respiration, neurite development, GABAergic transmission, and cellular stress responses. This expanded set of pathways likely reflects secondary pathological events (e.g., neuronal and synaptic dysfunction), probably perturbed in later stages of disease that may result from the primary gene driven causative events (e.g., APP processing and microglial efferocytosis)^75,76^. Indeed, when the Agora and Open Targets lists are filtered and re-sorted by Open Targets Genetics scores, the resulting pathway profile becomes significantly more aligned with that derived from the GVC list of candidate AD risk genes. These genetically focused analyses, highlight pathways involving Aβ clearance, cholesterol efflux, APP processing, immune response, lipid transport, and amyloid fibril, closely mirroring the GVC findings. This convergence strongly supports the critical importance of genetically driven analyses to accurately identify etiological pathways in AD, whereas the inclusion of transcriptomic and other types of data appears to introduce pathways reflecting secondary pathological changes rather than primary etiological mechanisms.

### High-Priority Targets

Our principled approach has identified high priority targets that can be further evaluated for treatment and diagnostics in AD as follows:

1. Functional Validation: Prioritized genes provide high-confidence targets for functional studies, such as CRISPR-based gene editing, RNA interference, or overexpression experiments in cell lines, organoids, or animal models. These approaches help clarify the biological role of the gene in AD mechanisms.
2. Pathway and Network Analysis: With a refined gene list, researchers can conduct pathway enrichment and gene network analyses to identify common biological processes or regulatory modules. This approach can help uncover converging mechanisms in AD and across neurodegenerative diseases, guiding hypotheses about disease etiology or progression.
3. Therapeutic Target Discovery: Genes/pathways with strong, verified associations are more likely to represent viable drug targets. Prioritization increases confidence that modulating the function of these genes/pathways could impact disease risk or progression, making them compelling candidates for drug screening and development efforts.
4. Biomarker Development: High confidence genes can be explored as potential biomarkers for early diagnosis, disease staging, or therapeutic response, especially if their expression or protein products are detectable in accessible tissues like blood or cerebrospinal fluid.
5. Model System Design: Animal and cellular models can be designed to reflect the genetic background of high-priority genes. This increases the relevance and translational potential of experimental findings, allowing for more precise testing of mechanistic hypotheses and interventions.
6. Integration with Multi-Omics Data: Verified genes can anchor integrative analyses including transcriptomic, epigenomic, and proteomic data, helping to contextualize genetic risk within broader biological systems and identify regulatory elements or non-coding variants involved in disease.

### Future Directions

Building upon our assessment and tiered classification of genetic association signals for AD identified by GWAS, future research should prioritize functional characterization of the top-tier loci to elucidate their biological relevance to AD pathogenesis. This includes, but is not limited to, integrating multi-omics data (e.g., transcriptomic, epigenomic, and proteomic) to understand the protein altering effects and regulatory mechanisms underlying the associated variants. Additionally, leveraging single-cell sequencing and spatial transcriptomic data derived from relevant brain tissues may provide insight into cell-type-specific effects, particularly since microglia/macrophages constitute a small proportion of cells in the brain but the most enriched cell populations for AD risk alleles. Further evaluation of these loci in large diverse populations is critical to assess the generalizability of these findings and to uncover population-specific variants. Finally, genes underlying these prioritized loci should be considered as more promising candidates for experimental modeling and therapeutic target validation, ultimately bridging the gap between genetic discovery and clinical translation in AD.

## Methods

### Phase 2. Systematic literature search and reviews

We conducted a structured literature search to identify published peer-reviewed articles reporting the results of genome- or exome-wide association studies (GWAS) of AD **(Figure. 1).** We searched the GWAS Catalog^77^ and PubMed via R packages (gwasrapidd v0.99.9 ^78^, easyPubMed v2.21 <u>file:///C:/work.stuff/ADSP/Gene verification committee/Manuscript 2023/13</u>) in September and August 2022, respectively. From the GWAS Catalog, we retrieved publications from studies with AD-related Experimental Factor Ontology (EFO) IDs: EFO_0000249, EFO_1001870 or MONDO_0004975 and publication date ≥ 2015-01-01.

From PubMed, we retrieved publications using the following queries:

- “Alzheimer’s Disease Sequencing Project” [CN]
- (“Alzheimer disease” OR “Alzheimer’s disease”) AND (whole-exome OR whole-genome) AND (genome-wide-association-study OR GWAS) NOT Review[PT] AND English[LA] AND 2010/01/01[EDAT]:2020/09/24[EDAT]
- (“Alzheimer disease” OR “Alzheimer’s disease”) AND (whole-exome OR whole-genome) AND (genome-wide-association-study OR GWAS) NOT Review[PT] AND English[LA] AND 2010/01/01[EDAT]:2022/08/16[EDAT]

A total of 304 publications were retrieved, and the corresponding reference data were exported from PubMed in XML format and imported into Covidence ^79^. Covidence recognized 303 of the 304 studies, of which 132 were removed as duplicates, leaving 171 studies for title and abstract screening. Within these, 124 studies were deemed irrelevant by at least two reviewers based on the following inclusion/exclusion criteria:

#### Inclusion criteria

- The phenotype is solely or mainly based on clinical diagnosis of AD (e.g., AD case/control status, age-at-onset of AD, or AD liability score). In addition to clinical diagnosis of AD, the phenotype can include other diagnostic measurements (e.g., neuropathological assessment or disease biomarkers).
- The study is a genome-wide association study of SVs or aggregations thereof.
- The article is published after 2015-01-01.
- The study is found in a search of the GWAS catalog, or it is an ADSP paper in PubMed.
- The analysis includes at least 1,000 total subjects.

#### Exclusion criteria

- The phenotype is a specific non-AD dementia or an endophenotype.
- The phenotype is solely based on neuropathological or biomarker outcomes.
- The article is a review.
- The analysis is performed using a novel method.
- The analysis is stratified (e.g., by sex or APOE status).
- The article was published before 2015-01-01.

The remaining 47 studies were assessed for eligibility based on full-text review and the same inclusion/exclusion criteria. Of these 47 studies, 8 studies were excluded due to inappropriate study design. The remaining 39 studies were assigned to at least two reviewers for extraction and review of GWAS associations.

### Phase 2. Pathway analysis of GVC, Agora, and Open Targets candidate AD genes

We conducted gene set over-representation analysis (ORA) of GVC (1344 genes), Agora, and Open Targets candidate AD gene lists using gprofiler2 package v 0.2.0 (See Data and Code availability information). We excluded electronic Gene Ontology (GO) annotations and filtered out results using a p-value <0.005 (corrected by g:SCS algorithm).

We used the following sorted gene lists for ORA analysis 1) Agora’s nominated genes ordered by Agora’s genetics, multi-omics, or target risk score; 2) Open Target’s AD associated genes ordered by decreasing Open Targets’ genetics portal or global score; 3) GVC gene list ordered by decreasingly Agora’s or Open Targets’ scores; 4) genes in common between the gene lists 1-3 (Figure 3A-B, Venn diagram built using Venn Diagram package v1.6.20 and three sets corresponding to gene lists 1-3, decreasingly ordered by the aforementioned Agora or Open Targets scores).

## Code availability

https://github.com/marcoralab/adsp_gvc_paper

https://github.com/marcora/gvc_agora_opentargets

## Data availability

Agora’s gene list of Alzheimer’s disease nominated targets (site version 3.4.0; data version syn13363290-v68) were downloaded from https://agora.adknowledgeportal.org/genes/nominated-targets. Agora’s gene scores (data version syn25741025-v12) were obtained from https://www.synapse.org/Synapse:syn25741025 on October 24^th^ 2024. We retrieved Open Targets’ gene list of Alzheimer’s disease (EFO:MONDO_0004975) associated targets (data version v24_09) from https://platform.opentargets.org/disease/MONDO_0004975/associations on October 24, 2024.

Curated gene and locus information used in this study is available through the TopGenes portal (https://topgenes.niagads.org/), a resource developed by the Alzheimer’s Disease Sequencing Project (ADSP) Gene Verification Committee that provides expert-reviewed evidence for genes and loci implicated in AD and related dementias.

## Data Availability

All data produced in the present work are contained in the manuscript.

https://topgenes.niagads.org/

## Acknowledgments

The authors thank Drs. Elizabeth Blue and Anita DeStefano for their early contributions to the literature review and development of the locus/gene review criteria.

## Grant Support and Potential Competing Interests

Dr. Leung, Mr. Katanic, Dr. Wang, and Dr. Schellenberg are supported by grants from the National Institutes of Health (U24AG041689, U54AG052427, and UO1AG032984).

Dr. Goate was supported by grants from the National Institutes of Health (U01AG058635, U01AG032984, and P30AG066514) and Freedom Together Foundation. Dr. Goate serves on scientific advisory boards for Genentech and Muna Therapeutics and is a consultant for Merck.

Dr. Farrer is supported by grants from the National Institutes of Health (R01AG048927, U01AG058654, U01AG062602, P30-AG072978, U19AG068753, R01AG080810, and U01AG082665). He serves as an advisor to ALZAI Health.

Dr. Kunkle is supported by grants from National Institutes of Health (U01AG058654, U01AG057659, U01AG062943, U01AG062943-02S1, U01AG076482, U01AG066767, and U01AG066767-02S1).

Dr. Mayeux is supported by National Institutes of Health grants R01AG067501, R01AG066107, U01AG076482, U24AG056270, and R01AG072474.

Dr. Naj is supported by grants from the National Institute on Aging (R01AG054060, U01AG066767, U01AG058654, RF1AG061351, U01AG062943, R01AG066152, U54AG052427, R01AG073435, and R56AG074604).

Dr. Marcora was supported by grants from the National Institutes of Health (U01AG058635 and R01HL168174).

Dr. Marcora serves on scientific advisory board for BrainStorm Therapeutics.

Dr. Patel was supported by a grant from the National Institutes of Health (U01AG058635).

Dr. Corces is supported by grants from the National Institutes of Health (P01AG073082, U01AG072573, and UM1HG012076).

Dr. Vardarajan is supported by grants from National Institutes of Health (R01AG067501, R01AG066107, U01AG066752, U01AG076482 and U01AG068028).

## Appendices

Appendix 1: Gene Verification Committee Mission Statement

Appendix 2: Single Variant Test Thresholds for Common Variants, Phase 1

Appendix 3: Phase 1 Gene-Based Test Thresholds

Appendix 4: Phase 2 Ranking Criteria: Single-Variant, Variant-Set, and Family-Based Analysis

Appendix 5: Phase 1 Locus/Gene Reports

## Boxes

#### Box 1. Phase 1 Tier Definitions: Single-Variant (SV) Analyses (see Appendix 2 for more detail)

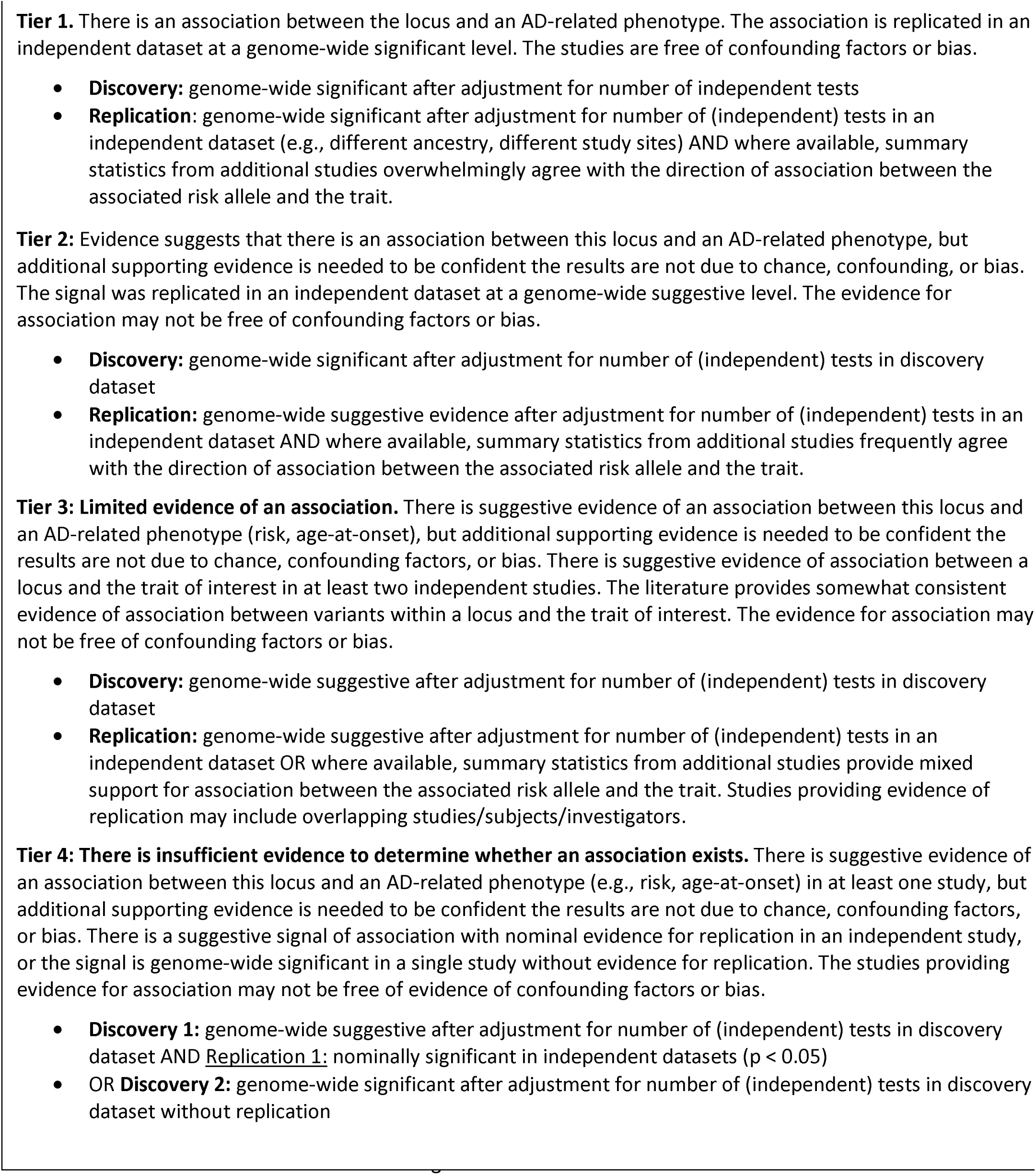

#### Box 2. Phase 2 Tier Definitions: Single-Variant (SV) Case-Control Analyses (see Appendix 4 for more detail)

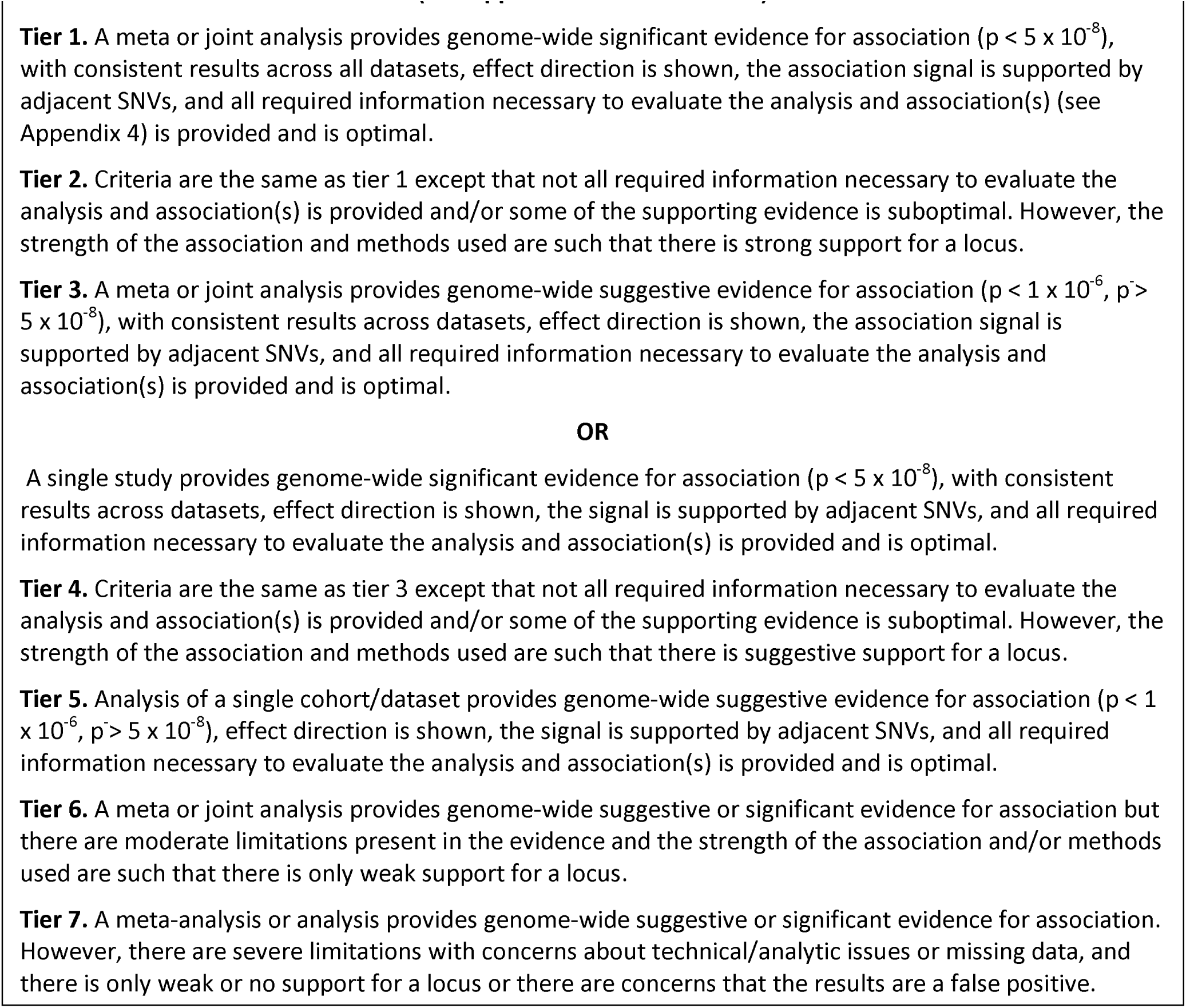

#### Box 3. Phase 2 Tier Definitions: Variant-Set (VS) Analyses (see Appendix 4 for more detail)*

| Criteria | Notes |
| --- | --- |
| Significance | Standard criteria are applied that consider number of genes/regions tested (e.g., FDR, Bonferroni correction, permutation testing).<br>Evidence comes from more than one SNV/SV (not in LD) for gene/region-based analysis.<br>Based on 20,000 genes (1/20,000) $5 \times 10^{-5}$ is considered “suggestive.” |
| Consistent Results | Results are consistent across datasets/cohorts.<br>Results from different ancestries can differ. |
| Robust Results (additional criteria) | Conditional analysis not needed.<br>LocusZoom is not needed.<br>If information on number of SNVs/SVs is missing, then rank as Tier 2 or 4. |
| Robust Analysis (additional criteria) | Batch effects are accounted for.<br>Sites are not equally covered between cases and controls.<br>Test performed accounts for LD between SNVs/SVs. |

<sup>a</sup>Tier assignment follows criteria outlined in Box 2.

#### Box 4. Phase 2 Tier Definitions: Family-Based Studies

Box 4. Phase 2 Tier Definitions: Family-Based Studies<sup>a</sup>
| Criteria | Notes |
| --- | --- |
| Significance | For linkage analysis, LOD score $\geq 3.6$ = genome-wide significance.<br>For linkage analysis, LOD score $\geq 2.0$ and $< 3.6$ = suggestive.<br>For association analysis, same as for single-variant case control (**Box 2**). |
| Robust Results<br>(additional<br>criteria) | Linkage plot should be presented.<br>LocusZoom is not needed. |
| Robust<br>Analysis<br>(additional<br>criteria) | Mendelian transmission patterns checked.<br>Relationship checks within and between families.<br>Batch effects checked (e.g., all subjects sequenced at the same center).<br>Family relations considered in association studies.<br>Within family ancestry taken into account if needed. |

<sup>a</sup> Tier assignment follows criteria outlined in Box 2. See Appendix 4 for more detail.

---

**Supplementary Figure S1.**
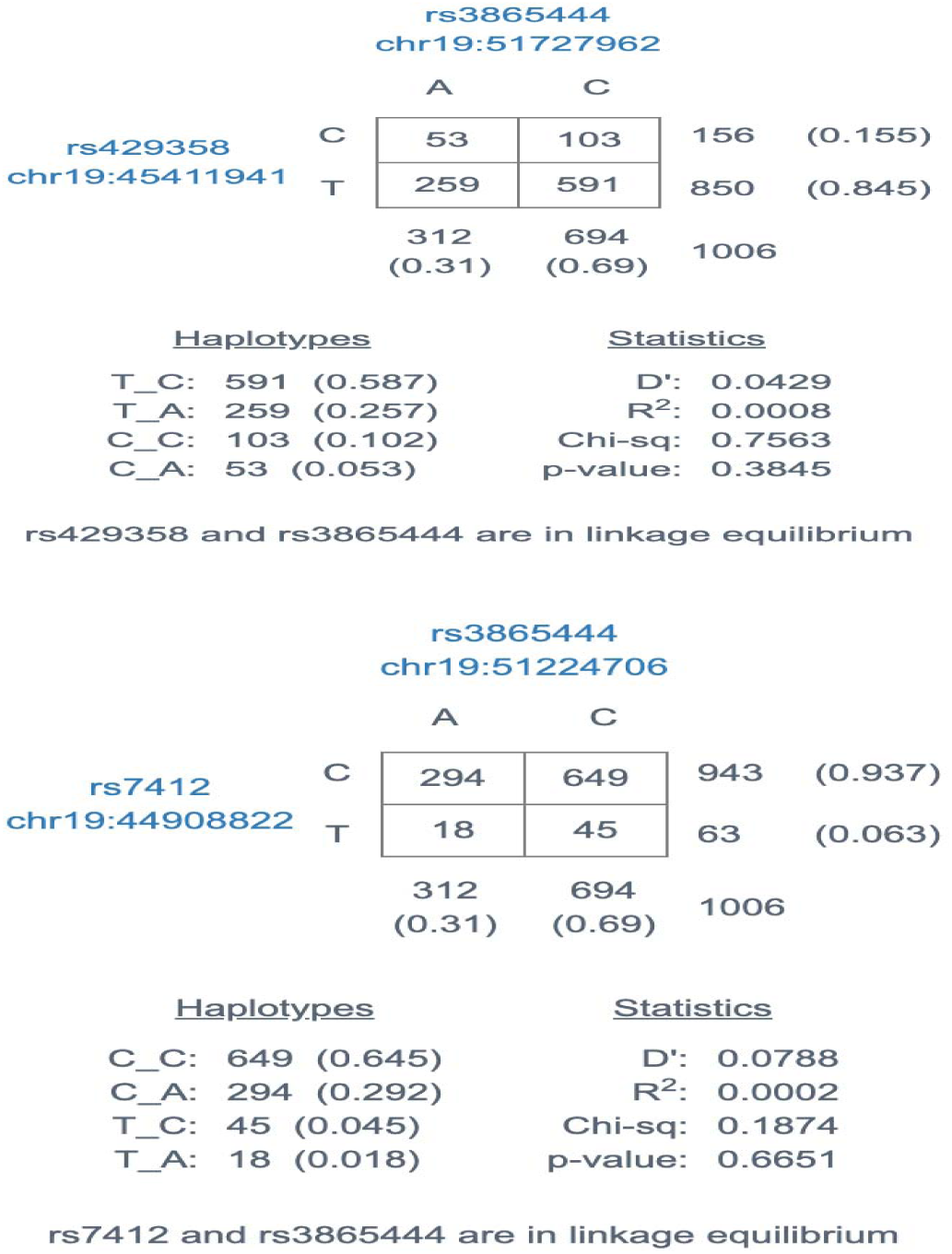
Linkage disequilibrium (LD) calculations from LD-link using GRCh38 assembly and all European ancestry populations.

**Supplementary Figure S2**

Target Risk Score, Genetic Risk Score, and Multi-omic Risk Score were obtained for every gene listed in the AD Knowledge Portal. Target Risk Score is the sum of the gene’s Genetic and Multi-omic Risk Scores and represents a gene’s general relevance to AD. Genetic Risk Score is a summary of genetic evidence supporting the target gene association with AD from multiple genetic studies. Scores range from 0 to 3, with higher scores indicating a greater likelihood of disease association. Multi-omic Risk Score is a summary of transcriptomic and proteomic evidence supporting the target gene association with AD from multiple studies. Multi-omic Risk Scores range from 0 to 2, with higher scores indicating a greater likelihood of disease association.

Target Risk score is shown in **Supplementary Figure S2A**; Genetic Risk score in **Supplementary Figure S2B**, Genetic Risk score, and Multi-comic Risk Score in **Supplementary Figure S2C** for the GVC genes (n = 484), Agora genes (n = 925), and Open Targets genes (n = 4,303). These genes are a subset of genes in Figure 3A and B with Agora scores. We displayed the scores by resource (x-axis), and raw scores are reported in the y-axis. The number of genes shown in the figures were smaller than those described earlier as some genes do not have scores in the AD Knowledge portal. We performed a non-parametric Wilcoxon test on scores from any of the two resources. Displayed p-values have undergone Bonferroni correction.

**Supplementary Figure S2A.**
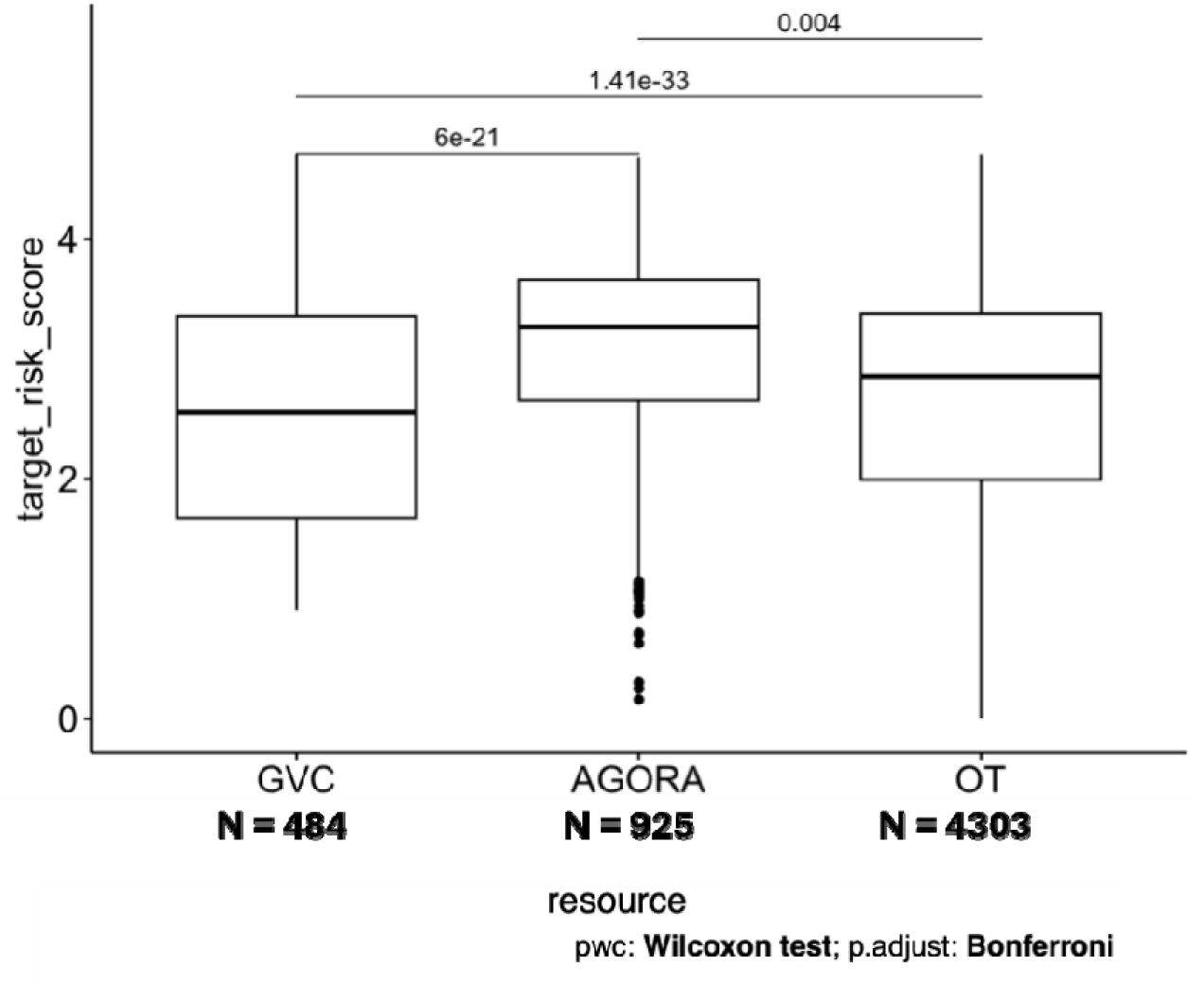
Target risk scores are significantly different across all three resources.

**Supplementary Figure S2B.**
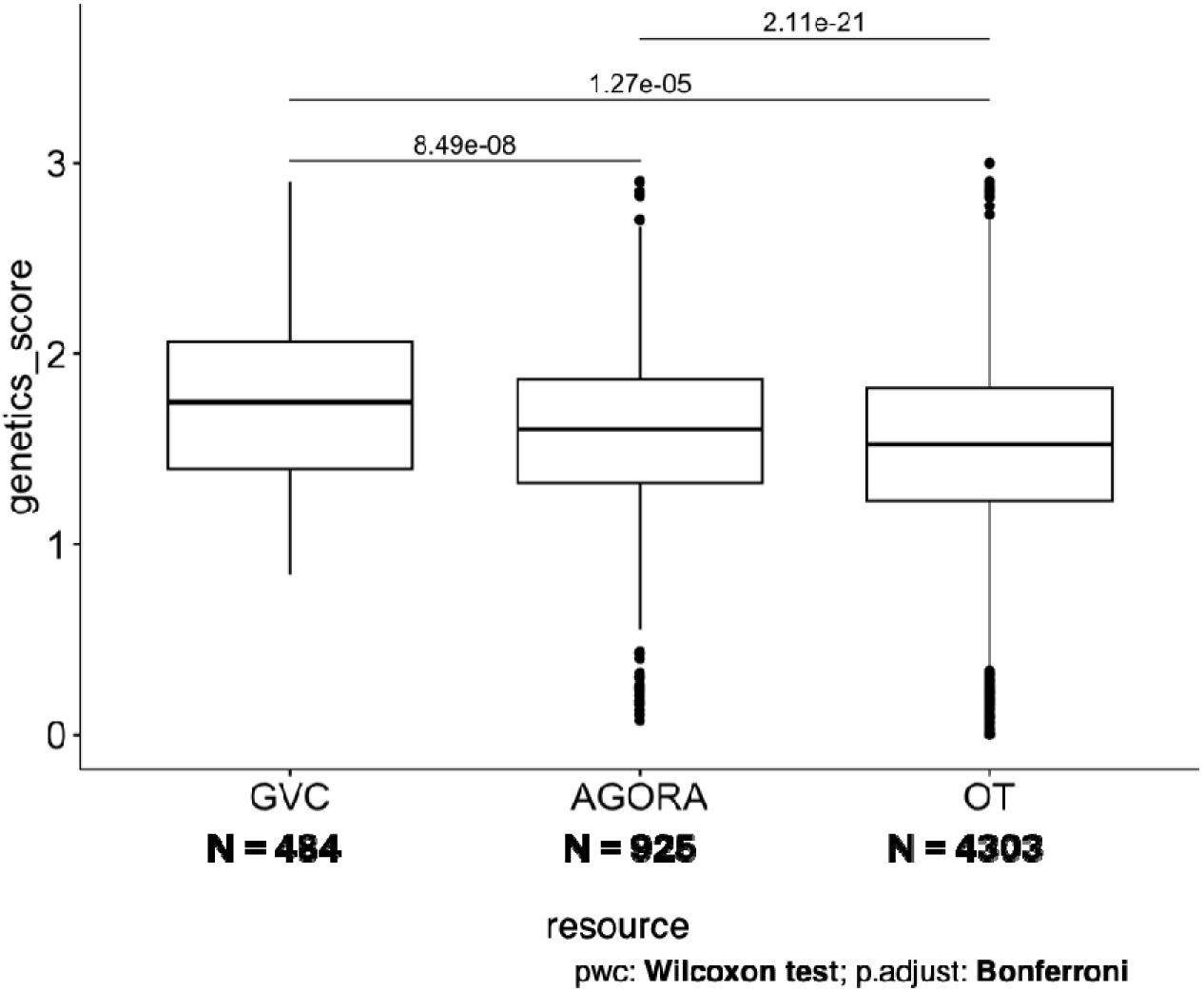
Genetic risk scores are similar between GVC and Agora but not Open Targets.

**Supplementary Figure S2C.**
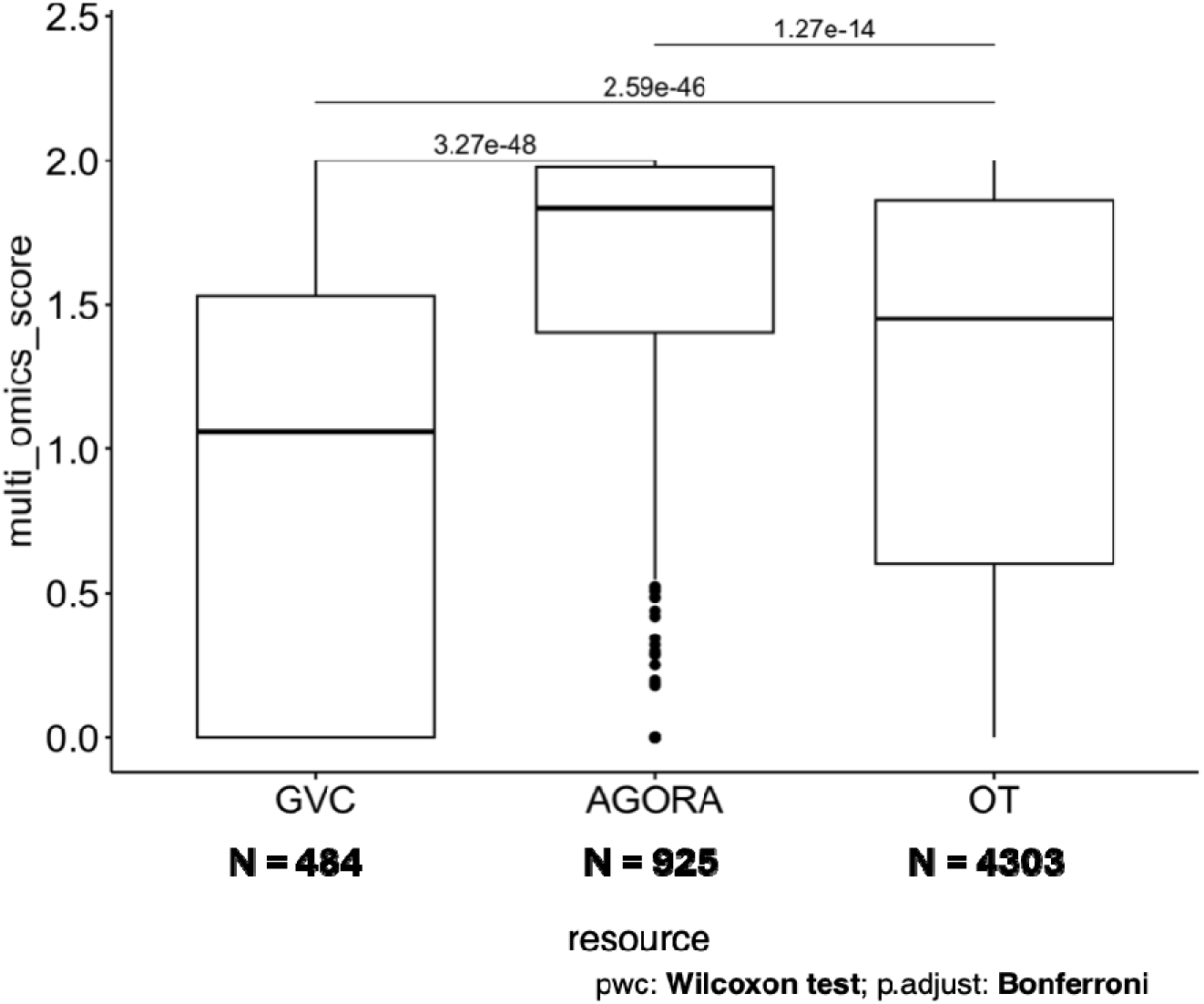
Multi-omics risk scores are significantly different across all three resources.

**Supplementary Figure S3.**
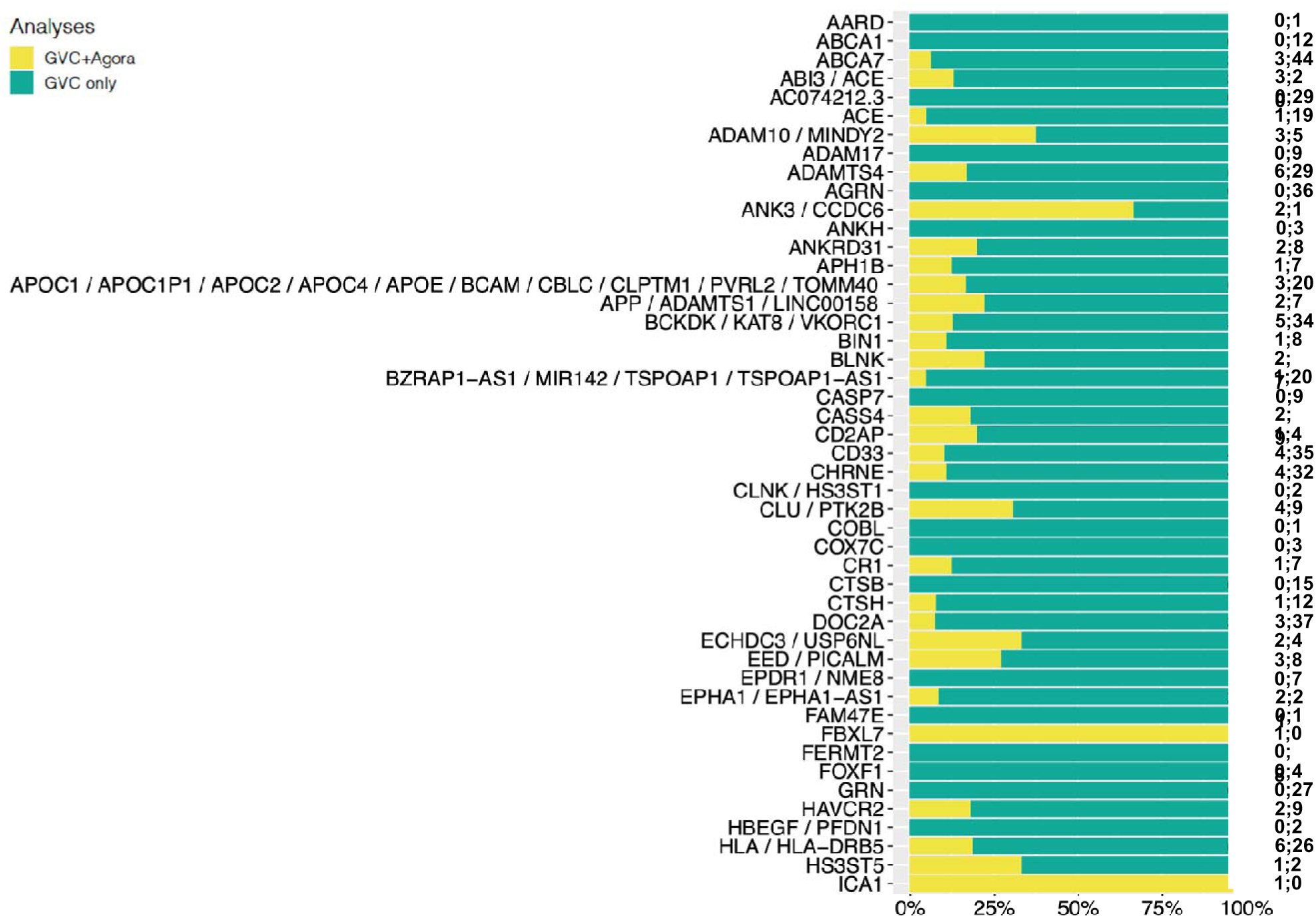
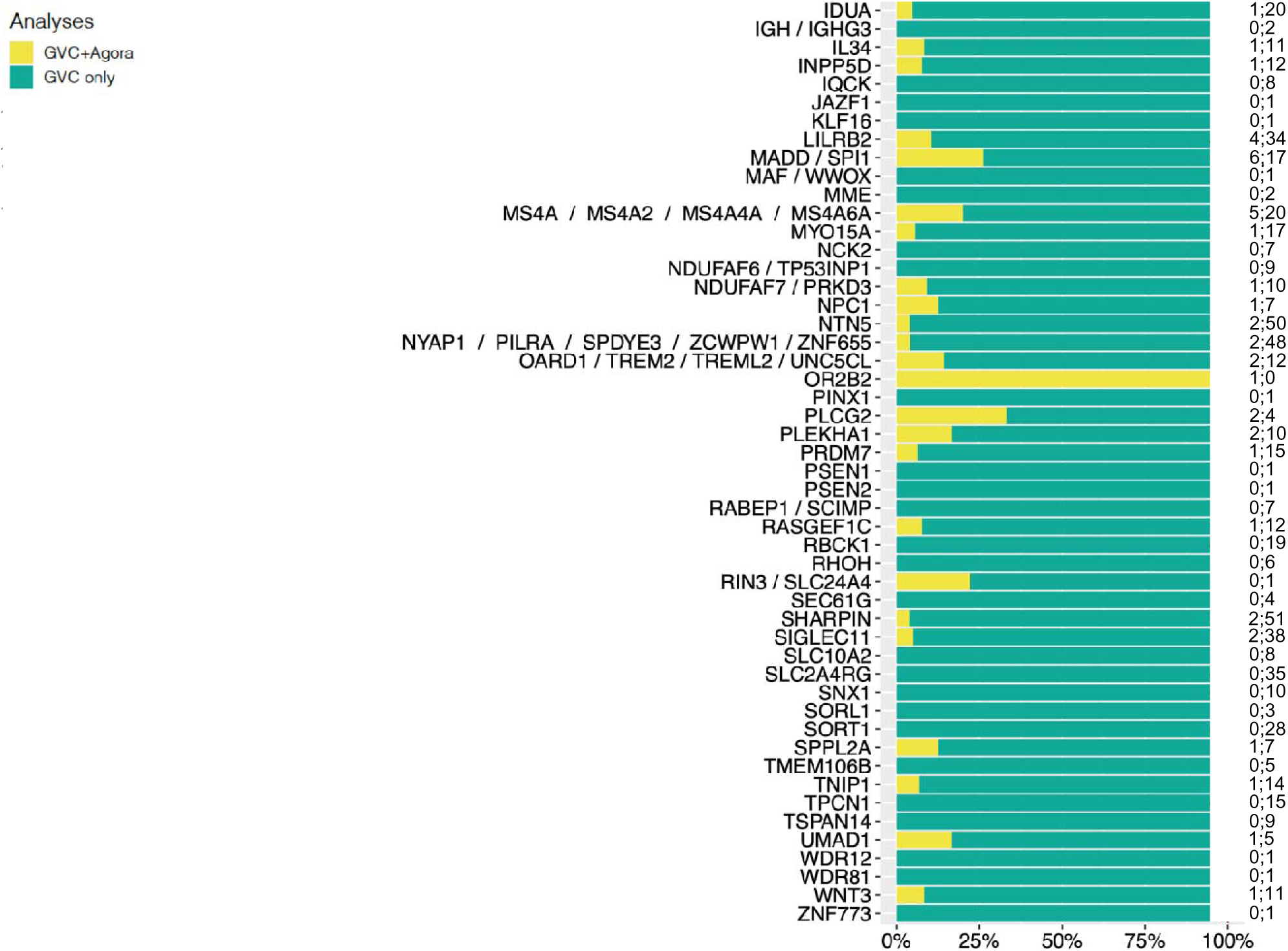
Comparison of 1,361 GVC genes from 500 kb bins (Fig. 3A) to 947 Agora genes. GVC genes used to define bins are given in the y axis on the left. The Agora gene names were compared to the GVC list to determine what fraction of the Agora genes are in the GVC list (X axis). Numbers on the Y axis on the right show the number of GVC genes that overlap Agora genes, followed by the number of genes found only in the GVC list.

**Supplementary Figure S4.**
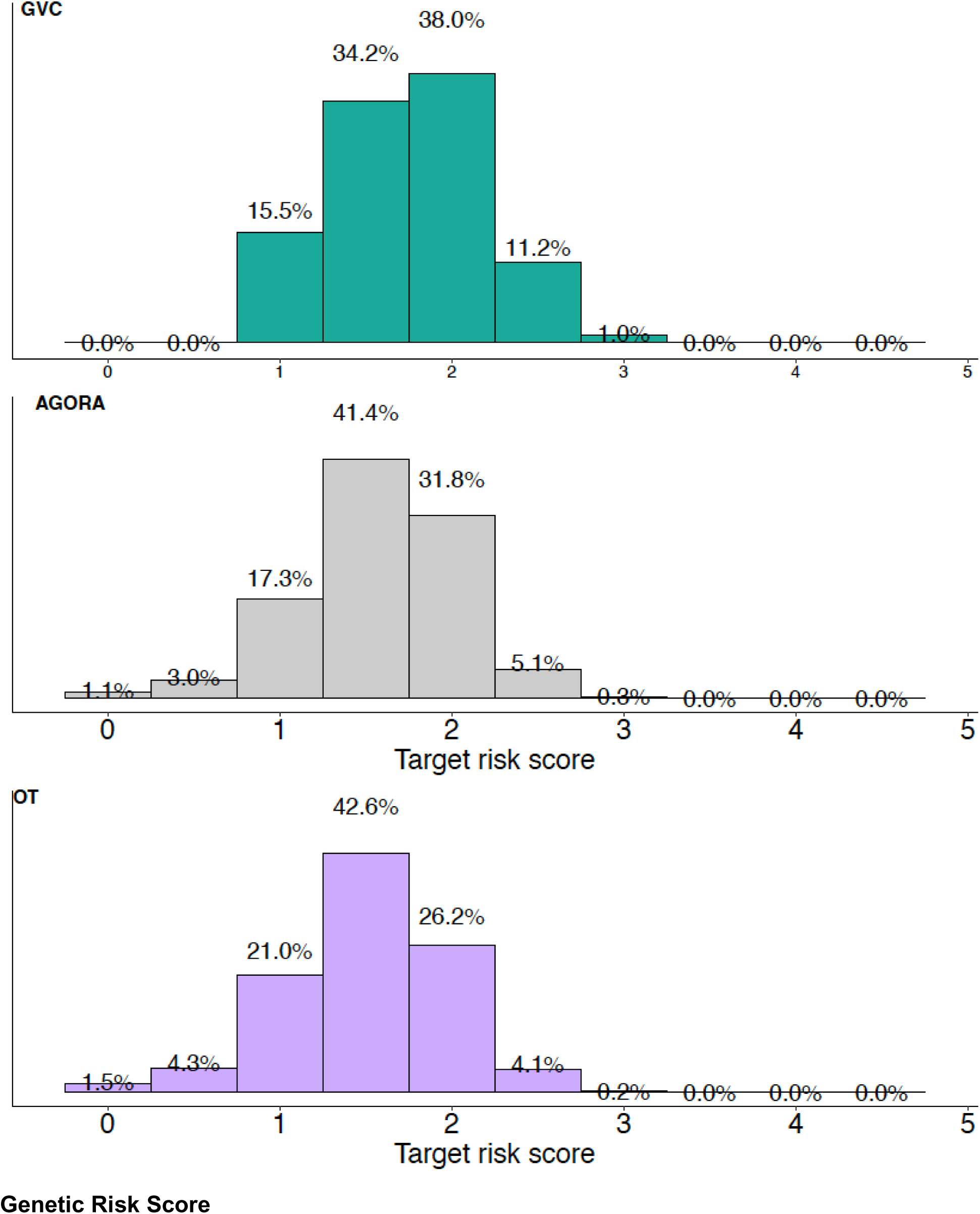
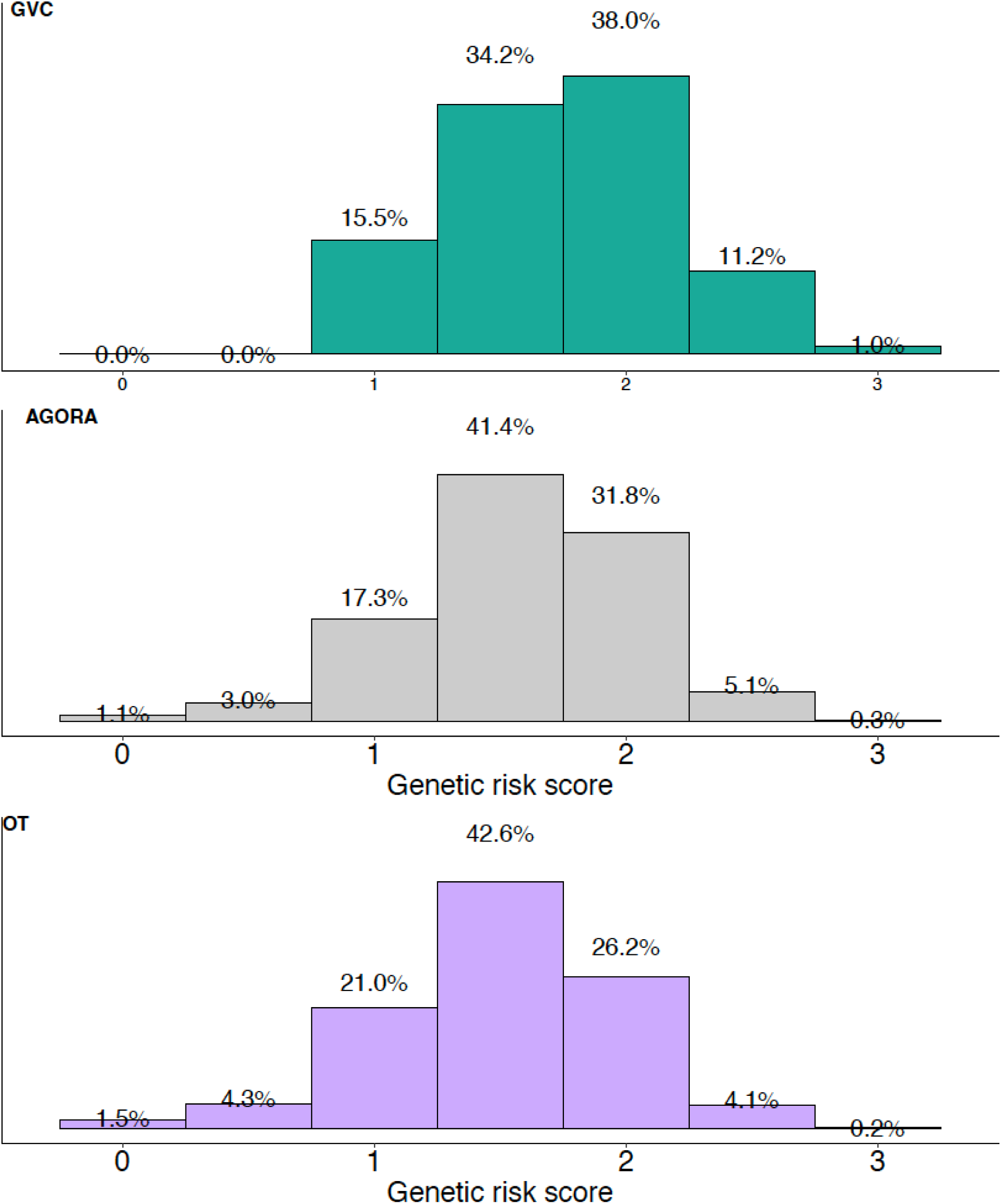
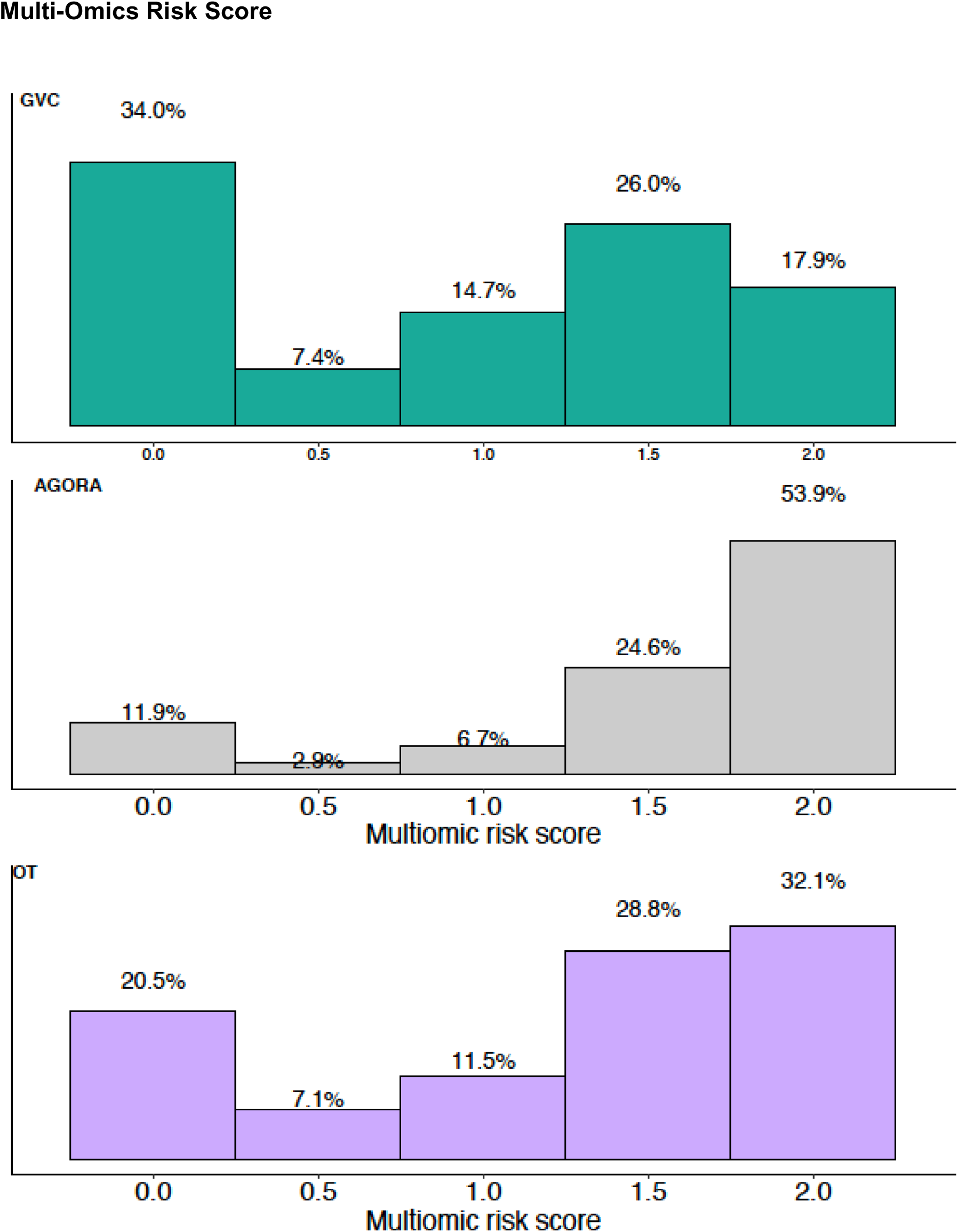
Supplementary Figure 2 in binned version. Target Risk Score

## Notes

### Competing Interest Statement

The authors have declared no competing interest.

### Funding Statement

P01AG073082
P30AG066514
P30AG072978
R01AG048927
R01AG054060
R01AG066107
R01AG066152
R01AG067501
R01AG072474
R01AG073435
R01AG080810
R01HL168174
R56AG074604
RF1AG061351
U01AG032984
U01AG057659
U01AG058635
U01AG058654
U01AG062602
U01AG062943
U01AG066752
U01AG066767
U01AG068028
U01AG072573
U01AG076482
U01AG082665
U19AG068753
U24AG041689
U24AG056270
U54AG052427
UM1HG012076

