## Appendices for "Comprehensive adjudication identifies 111 high-confidence loci for Alzheimer’s disease and related dementias"

Appendix 1: Gene Verification Committee Mission Statement

Appendix 2: Single Variant Test Thresholds for Common Variants, Phase 1

Appendix 3: Phase 1 Gene-Based Test Thresholds

Appendix 4: Phase 2 Ranking Criteria: Single-Variant, Variant-Set, and Family-Based Analysis

Appendix 5: Phase 1 Locus/Gene Reports

Note: Names of tables and figures included in the Appendices begin with an A. Tables and figures included in supplementary materials begin with an S.

**Gene Verification Committee Mission Statement: Appendix 1**

**November 5, 2025**

**AD therapeutic targets.** Retrospective analysis shows that therapeutic targets supported by genetic evidence are 2–3 times more likely to succeed compared to targets not supported by genetic findings.^1^ Thus, genetic-based gene discovery work for Alzheimer’s disease (AD) and related disorders (ADRD) including all causes of related dementias are a validated source of therapeutic targets. Going from a genetic locus to a therapeutic target is a resource-intensive process, and thus it is critical that the genetic evidence supporting a candidate therapeutic target be carefully evaluated.

**Gene Verification Committee (GVC).** A large number of publications report genetic evidence that a gene or locus influences or causes AD. The quality of evidence presented in these publications is highly variable. To clarify which loci and genes are valid versus potential false positives, the GVC is reviewing published human genetic studies of AD and dementia to determine the quality of evidence supporting a genetic signal (locus) or a specific gene.

**Genome-wide association (GWAS) signals.** GWAS signals identify loci that are regions of the genome where one or more ADRD/dementia risk genes are present. GWAS studies are a powerful method and have identified a large number of AD-associated loci. However, the region associated with ADRD/dementia may contain several genes, and the actual causative gene(s) is/are not clearly identified. To evaluate published genetic studies, the GVC performed a structured literature search to select publications to evaluate and developed structured criteria for evaluating genetic associations (Appendix 4). A tier system was developed (ranking 1–7) to identify the quality of evidence, with tier 1 loci having the highest quality of evidence supporting an association. Table 3 in the body of the paper lists 97 unique tier 1 loci with genome-wide evidence of association with AD/dementia. The most recent large GWAS study found 75 of these sites as genome-wide significant loci.^1^

**Causative AD/dementia genes.** In some cases, the actual causative gene is known or can be inferred using bioinformatic approaches. These genes are located at loci where genome-wide evidence of association or linkage exists. Causative genes are being identified by two processes.

1. **Expert evaluation of supporting evidence**. A list of 23 genes were evaluated by an expert panel that reviewed both genetic and functional evidence. The result was a list of high-confidence genes causative for AD genes (Table 1). Note that for four of these genes, the encoded protein has been the target of drug trials for either AD or an ADRD (*APP, PSEN1, APOE*).
2. **Bioinformatic approaches**. More recently, the GVC has begun to use bioinformatic approaches to identify high-confidence causative genes including Open Targets^2^ (Table 3) and Inferno.^3^

**Single Variant Test Thresholds for Common Variants**

**Phase 1: Appendix 2**

**Tier 1: Sufficient evidence of an association.** Evidence is sufficient to conclude that there is an association between this locus and an AD-related phenotype (ex., risk, age-at-onset). This means that there has been a strong signal of association between a locus that has been replicated in an independent data set at a genome-wide significant level. The literature provides consistent statistical evidence of association between variants within a locus and the trait of interest. The studies providing evidence for association are free of evidence of confounding or bias.

- **Discovery:** genome-wide significant after adjustment for number of (independent) tests in discovery data set
- **Replication:** genome-wide significant after adjustment for number of (independent) tests in an independent data set (e.g., different ancestry, different study sites, etc.) AND where available, summary statistics from additional studies overwhelmingly agree with the direction of association between the associated risk allele and the trait.

**Tier 2: Suggestive evidence of an association.** Evidence suggests that there is an association between this locus and an AD-related phenotype (ex., risk, age-at-onset), but additional supporting evidence is needed to be confident the results are not due to chance, confounding or bias. This means that there has been a strong signal of association between a locus that has been replicated in an independent data set at a genome-wide suggestive level. The literature provides somewhat consistent evidence of association between variants within a locus and the trait of interest. The studies providing evidence for association may not be free of evidence of confounding or bias.

- **Discovery:** genome-wide suggestive evidence for an association after adjustment for number of (independent) tests in discovery data set
- **Replication:** genome-wide suggestive after adjustment for number of (independent) tests in an independent data set AND where available, summary statistics from additional studies frequently agree with the direction of association between the associated risk allele and the trait.

**Tier 3: Limited evidence of an association.** There is suggestive evidence of an association between this locus and an AD-related phenotype (e.g., risk, age-at-onset), but additional supporting evidence is needed to be confident the results are not due to chance, confounding or bias. This means that there has been a suggestive signal of association between a locus and the trait of interest in at least two independent studies. The literature provides somewhat consistent evidence of association between variants within a locus and the trait of interest. The studies providing evidence for association may not be free of evidence of confounding or bias.

- **Discovery:** genome-wide suggestive after adjustment for number of (independent) tests in discovery data set
- **Replication:** genome-wide suggestive after adjustment for number of (independent) tests in an independent data set OR where available, summary statistics from additional studies provide mixed support for association between the associated risk allele and the trait. Studies providing evidence of replication may include overlapping studies/subjects/investigators.

**Tier 4: Insufficient evidence to determine whether an association exists.** There is suggestive evidence of an association between this locus and an AD-related phenotype (ex., risk, age-at-onset) in at least one study, but additional supporting evidence is needed to be confident the results are not due to chance, confounding or bias. This means that there has been a suggestive signal of association between a locus and the trait of interest in a study with nominal evidence for replication in an independent study, or the signal is genome-wide significant in a single study without evidence for replication. The literature may not provide consistent evidence of association between variants within a locus and the trait of interest. The studies providing evidence for association may not be free of evidence of confounding or bias.

- **Discovery 1:** genome-wide suggestive after adjustment for number of (independent) tests in discovery data set AND Replication 1: nominally significant in independent data sets (p < 0.05)
- OR **Discovery 2:** genome-wide significant after adjustment for number of (independent) tests in discovery data set without replication

**Phase 1 Gene-Based Test Thresholds: Appendix 3**

1. Tier 1: highly confident
   a. p< 2.5x-10-6 and replication (p<.05/n where n=number of genes tested)
2. Tier 2: likely
   a. p< 2.5x-10-6 (but no replication) or
   b. suggestive p that reaches p< 2.5x-10-6 in meta-analysis
3. Tier 3: of interest
   a. p<5x10-6 and replication or
   b. p<5x10-6 p-value that reduces in meta-analysis
4. Tier 4: of lower interest
   a. p<1x10-5

Other criteria

A. Information should include whether the gene-based test was specifically a rare variant test.
B. Filtering criteria on variants included in gene-based test?
C. Type of gene-based test?
D. Number of variants driving result?
E. Minimum sample size for either main study or replication

**Phase 2 Ranking Criteria: Single-Variant, Variant-Set, and Family-Based Analysis: Appendix 4**

**Study selection.** The Gene Verification Committee (GVC) performs literature reviews of large-scale genome-wide association studies (GWAS) of Alzheimer’s disease (AD) and closely related phenotypes, including studies of array genotypes, whole exome sequencing (WES) and whole genome sequencing (WGS) data. We also review studies identified by expert investigators (GVC group) and review loci listed in the Alzheimer’s Disease Variant Portal (ADVP) and the National Human Genome Research Institute/[European Bioinformatics Institute](https://en.wikipedia.org/wiki/European_Bioinformatics_Institute) (EBI) GWAS catalog. Reports from BioRxiv or medRxiv are not considered. See Figure 1 for full selection criteria. The following types of publications are examined but not formally reviewed:

1. Publications with sample sizes less than 1,000 subjects.
2. Publications where novel untested methods were used. A novel method can be reviewed if it produces results consistent with those of more traditional methods.

**Evaluation.** Association signals from these studies are classified into seven tiers, based on the evidence supporting them. Each manuscript with at least one locus having suggestive or genome-wide significant evidence is then reviewed by the GVC to evaluate the evidence that the particular signal is associated with AD. Evidence from the latest/largest study may supersede evidence from previous studies when samples overlap, and analyses are comparable. Adjudication will determine which study will be given top priority.

**Phenotypes.** The GVC considers multiple phenotypes closely related to AD. We consider each phenotype independently and rank each separately. Under this system, a given locus will have multiple rankings depending on the phenotypes reviewed. Current phenotypes under consideration are:

- **AD** defined either clinically (possible or probable AD) or by autopsy (confirmed AD).
- **Dementia** is defined as evidence of cognitive impairment, and includes information collected from self-report or by proxy.

The most specific phenotype considered is clinically diagnosed and autopsy-confirmed AD, whereas the family history of AD and dementia class and clinically diagnosed probable or possible AD without autopsy or biomarker confirmation will include AD as well as other causes of dementia. In some studies, subjects diagnosed with AD are mixed with dementia cases in a way where the two subject classes cannot be distinguished; in these cases, we consider them as “Dementia” studies. Future phenotypes may include AD endophenotypes (e.g., CSF and plasma biomarkers), AD related disorders (ADRD) and stratified analysis (i.e., *APOE*, sex) of AD phenotypes.

**Sources of evidence.** Single variant, linkage, and variant-set association analysis are reviewed. To assign a locus to a tier, the GVC gathers evidence for the strength of association between the variant or another in strong linkage disequilibrium (LD), the consistency of the signal within the study, the robustness of the results, and the robustness of the analysis. Family-based association and linkage analyses, and variant-set association analyses are evaluated similarly, but include additional supplementary criteria.

Specifically:

- **Significance:** Evidence for association between the AD-related phenotype and a variant or an LD proxy is measured by the p-value (*P*).
  - **Genome-wide significance** is defined using standard criteria (P < 5 x 10^-8^). We recognize that this value may be too high when populations other than non-Hispanic Whites (non-Spanish European origin subjects) are considered or for rare-variant analyses.
  - **Genome-wide suggestive evidence** is defined using standard criteria (P < 1 x 10^-6^).
  - **Nominal evidence** is defined as *P* < 0.05.
- **Consistent association** within a meta-analysis.
  - **Within the final meta-analysis**, the direction of effect is mostly the same for the different cohorts/datasets included within the meta-analysis, and results are not driven by a single data set among data sets representing similar populations (AD clinical/neuropath and Dementia are not considered similar populations).
  - Forest plots, effect size statistics (odds ratios, beta coefficients, etc.) and i^2^ heterogeneity values (or equivalent data) should be presented so that heterogeneity of individual results within the meta-analysis can be evaluated.
- **Robust results.**
- Conditional analysis for adjacent signals are presented when applicable.
- Support from adjacent SNVs near the lead SNV (common SNVs only). LocusZoom plots showing multiple SNVs for a signal (or equivalent data) should be provided for novel signals**.** “Equivalent data” are association results in tabular form from multiple adjacent SNVs (~ +/- 1 Mb).
- **Robust analysis.** Example signatures of solid associations include:
  - Evidence for genomic inflation is evaluated and reported with a genomic inflation factor near 1. Q-Q plots should be provided but are not required.
  - Population stratification is evaluated and accounted for in the analysis model.
  - Standard quality control methods are applied (e.g., relatedness, call rate, imputation quality, and DNA sample quality filtering).
  - Batch effects such as coverage harmonization are considered and accounted for including quality control (to assess the presence of batch effects and other con founders) and model specification (inclusion of batch effects and other con founders when appropriate). Example strategies include:
    - Depth and exome capture kits are the same across cases and controls or are controlled/modeled for in the association tests.
    - Accounting for sequencing site and/or technology in the analysis model.
    - Quality control filtering including standard methods such as sites not equally covered between cases and controls are excluded from analysis.
  - Proper choice of association test including accounting for imbalance in cases and controls if applicable.
  - Significance thresholds are appropriate (genome-wide, acknowledges multiple testing).
- **Additional criteria for consideration:**
  - Sub-threshold evidence of association in an independent data set (*P* < 0.05), such as one from a different ancestry, should be recorded in the comment field of the locus report.
  - Note: Disagreement in results from another ancestry is not a reason for exclusion from a specific tier.

**Tier System.** To begin, we evaluate each analysis within the publication. If it is a meta-analysis, we only evaluate the final meta-analysis results and not each stage contributing to the meta-analysis. Both within ancestry and across ancestry results are evaluated for multi-ancestry meta-analysis. Each locus reported within the main text of the publication is ranked based on the strength of evidence for association. See **Table A1** for a summary of each tier requirements. Once tiers have been assigned for all analysis and publications, shared loci are defined using FUMA or a similar program. If the tier ranking of a locus is not the same across different publications or analysis, the GVC adjudicates the tier assignments based on sample size and overlap, phenotypes used, ethnic composition of the studies, quality of the analyses, consistence of effect direction, and support from adjacent SNVs.

Below and in Table A1, “meta-analysis” refers to evaluation of multiple cohorts/data sets. “Analysis” implies that only one data set/cohort is considered in the evaluation. The definition of a “cohort” or “data set” is made by the authors of the manuscript under review.

**Family Study Criteria.** Family-based study criteria apply to studies which are predominantly family-based (i.e., where > 50% of the sample comes from families). These studies are evaluated based on several other criteria in addition to, or in place of, the above evaluation criteria. Note that these additional criteria fall within the same general criteria categories (i.e., methods – appropriate QC) as case-control study criteria and therefore do not require additional evaluation categories. The additional criteria include:

- **Significance:** For studies performing linkage and association, criteria include evaluation with a LOD score with significance defined as standard criteria of 3.6 LOD for genome-wide significance and 2.0 LOD for suggestive significance. Significance for association studies remain the same as for case-control studies.
- **Additional robust results criteria**:
  - For linkage studies, a linkage plot should be presented instead of a Locus zoom plot.
- **Additional robust analysis criteria:**
  - Mendelian transmission pattern checks were performed.
  - Relationship checks of pedigrees, both within and between families, are performed.
  - Batch effects strategies for families are accounted for, such as sequencing all members of a family at the same center.
  - Family relationships are taken into account in association testing.
  - Within family ancestry is taken into account if necessary (i.e., multi-ancestry families).

**Variant-set Association Analysis Criteria.** The GVC also reviews publications where multiple variants are considered together for evidence of association with AD or dementia. These are often called “gene-based” methods, but analysis of a region containing a specific gene or a sliding window approach are closely related. We use the term ‘variant-set analysis’ to describe methods that test the association between a set of variants (e.g., all coding variants within a gene) with the trait of interest. As with family-based studies, criteria for these analysis are evaluated based on several other criteria in addition to, or in place of, the evaluation criteria described for single variant association analysis. In contrast to single variant results where we evaluate the final meta-analysis results only for meta-analysis, for variant-set analysis we evaluate both the ‘main discovery analysis results’ of the manuscript and any final meta-analysis results, as replication of variant-set analysis can have several limitations including possible missingness of key variants driving the discovery result in the replication analysis data set(s).

**Significance**: Genome-wide significance using standard criteria, such as permutation testing, false-discovery rate (FDR) or Bonferroni correction for the number of sets tested (significant after correcting for the number of sets tested (P < 0.05/n where n=number tested), in at least one large-scale robust meta-analysis of the same phenotype, or in two or more robust independent studies of the same phenotype. The signal comes from more than one single nucleotide variant (SNV) or structural variant and these multiple variants are not in strong LD (i.e.*,* LD is accounted for in the test). Suggestive significance using standard criteria, such as permutation testing, false-discovery rate (FDR) or Bonferroni correction for the number of regions/genes tested (significant after correcting for the number of genes/aggregated units/genomic segments tested P < 1/n where n=number tested), in at least one large-scale robust meta-analysis of the same phenotype, or in two or more robust independent studies of the same phenotype. Alternatively, a single study may have genome-wide significance using standard criteria as described in Tier 1. The signal comes from more than one single nucleotide variant (SNV) or structural variant and these multiple variants are not in strong LD (i.e., LD is accounted for in the test). If not specified and the unit of aggregation is a gene, the standard 1/20,000 genes (5x10^-5^) should be used for the suggestive significance level.

- **Consistent results:** If tests are performed across multiple data sets or stages, then consistency of results should be evaluated. Note that this criteria should not be applied to across ancestry analysis, though, as with single variant association analysis, observations on the agreement or lack there-of between cross-ancestry results can be recorded in the comments field.
- **Additional robust results criteria:** Conditional analysis and LocusZoom plots criteria do not apply.
  - Variant-set association analysis should include two or more variants per test unit. Significant or suggestive variant-set associations that are driven by only one variant OR don’t provide information to evaluate this should be considered as having ‘some sources of evidence are missing, or evidence is not optimal’ and thus should be placed in Tier 2 or Tier 4, assuming they meet all other criteria for these tiers.
- **Additional robust analysis criteria:**
  - Batch effects are accounted for including sites not equally covered between case and controls are excluded from analysis.
  - The test performed accounts for LD of variants with the set if applicable.

| **Tier 1: Sufficient evidence of an association**  A meta or joint analysis provides:   - Genome-wide significant evidence for association   (Col U)   - Robust analysis   (Col AO decision)   - Consistent association   (Col AA decision)   - Robust association (Col AE decision) - All required information (see Table A2) necessary to evaluate the analysis and association signals is provided and is fully supportive of a robust/consistent evaluation.   (i.e., all option “1”s in cols AO, AA and AE)   - All data/analysis is provided to evaluate effect direction and support from adjacent SNVs for a signal   (cols AB, AC and AD, and/or AE), | **Tier 2 Sufficient evidence of an association but some sources of evidence are missing, or evidence is not optimal.**  A meta or joint analysis provides:   - Genome-wide significant evidence for association   (Col U)   - Not all required information (see Table A2) necessary to evaluate the analysis and associations is provided and/or some of the information is suboptimal. - However, the strength of the association and methods used are such that there is strong support for a locus.   (col U and AF to AL? (judgement calls for these methods whether they are good or not)  OR   - In an AD + Dementia study, most of the statistical evidence comes from either Dementia cohorts (e.g., UKBB or 23&Me) or AD clinic/path cohorts. | **Tier 3: Suggestive evidence of an association**  A meta or joint analysis provides:   - Genome-wide suggestive evidence for association - Robust signal - Consistent association - Robust association - All data/analysis is provided to evaluate effect direction and support from adjacent SNVs for a signal.   .  **OR**  Analysis of a single cohort/dataset provides:   - Genome-wide significant evidence for association - Robust analysis - Robust association - All required sources of evidence are provided, and the evidence is optimal | **Tier 4: Suggestive evidence of an association but some sources of evidence are missing, or some evidence is suboptimal.**  A meta-analysis or joint analysis provides:   - Genome-wide suggestive evidence for association   Not all required information (see Table A2) necessary to evaluate the analysis and associations is provided and/or some of the information is suboptimal  **OR**  Analysis of a single cohort/dataset* provides:   - Genome-wide significant evidence for association - Not all required information (see Table A2) necessary to evaluate the analysis and associations is provided and/or some of the information is suboptimal | **Tier 5: Suggestive evidence of an association from a single data set**  Analysis of a single cohort/dataset* provides:   - Genome-wide suggestive evidence for association - Robust analysis - Robust association - All required sources of evidence are provided, and the evidence is optimal | **Tier 6: Limited evidence for an association**  A meta or joint analysis provides:   - Genome-wide suggestive or significant evidence for association   There are moderate limitations present in the evidence for a robust signal, and/or a consistent association or robust association.   - The strength of the association and/or methods used are such that there is only weak support for a locus. | **Tier 7: Insufficient evidence of association**  A meta-analysis or analysis provides:   - Genome-wide suggestive or significant evidence for association - Severe limitations, such as: - Evidence the association is a false positive. - Concerns about technical/analytic issues - Data necessary to evaluate the robustness and consistence of the analysis are missing such that there is only weak or no support for a locus.   **OR**   - No suggestive or genome-wide significant evidence is provided |
| --- | --- | --- | --- | --- | --- | --- |

**Table A1. Summary of criteria for tier system**

**Tier 1: Sufficient evidence of an association.** Evidence is sufficient to conclude that there is an association between this locus and the phenotype based on the following:

•	There is a large-scale meta-analysis providing genome-wide significant statistical evidence of a genetic association between a variant and the trait of interest.

•	Association results are consistent across the cohorts used in the final meta-analysis presented in the main body of the paper.

•	All information is provided to support a robust association as defined below.

•	All information is provided to support a robust analysis.

•	Analysis methods are well-documented and are supported by other works in the literature.

•	All data/analysis is provided to evaluate effect direction.

•	Support from adjacent SNVs (up to +/- 1 Mb) support the signal from the peak SNV (either Locus Zoom plot or tabular data)

**Tier 2: Evidence of an association but some information missing or not optimal.**

•	A large-scale meta-analysis providing genome-wide significant statistical evidence of a genetic association between a variant and the trait of interest. At least one variant meeting the inclusion criteria, or its LD proxy, has genome-wide significant evidence for association with the AD-related trait.

•	Not all required information necessary to evaluate the analysis and associations is provided and/or is not fully supportive of robust/consistent evaluation.

•	The reviewer will note what information is missing and/or other supportive evidence.

•	Analysis methods are well-documented and are supported by other works in the literature.

•	The strength of the association and methods used are such that there is strong support for a locus.

**OR**

•	In a study that includes both cohorts where the primary phenotype is AD defined by clinical and/or neuropathologic data and where the primary phenotype is Dementia (e.g., UKBB or 23&Me or other dementia cohorts), most of the statistical evidence comes from either the Dementia cohorts or from AD clinical/pathology cohorts.

**Tier 3: Suggestive evidence of an association.** Evidence suggests that there is an association between this locus and the phenotype.

Tier 3 has:

•	A large-scale meta-analysis provides suggestive evidence of a genetic association variant and the trait of interest.

•	Results are consistent across the cohorts used in the final meta-analysis presented in the main body of the paper.

•	All information is provided to support a robust association.

•	All information is provided to support a robust analysis defined below.

•	Analysis methods are well-documented and are supported by other works in the literature.

•	All data/analysis is provided to evaluate effect direction and support from adjacent SNVs for a signal.

**OR**

•	Genome-wide significant association at a locus in a single cohort.

•	Robust analysis.

•	Robust association.

•	All required sources of evidence are provided, and the evidence is optimal.

•	All data/analysis is provided to evaluate effect direction and support from adjacent SNVs for a signal.

**Tier 4: Suggestive evidence of an association but some information is missing or suboptimal.**

•	A large-scale meta-analysis providing suggestive evidence of a genetic association between a variant and the trait of interest.

•	Not all the required information is provided to conclude that the analysis is robust.

•	Not all the required information is provided to determine that the association is robust.

•	Analysis methods are well-documented and are supported by other works in the literature.

•	The strength of the association and methods used are such that there is support for a locus.

OR

•	Analysis of a single cohort that yields genome-wide evidence for an association.

•	Not all required information (see Table AT2) necessary to evaluate the analysis and associations is provided and/or some of the information is suboptimal.

**Tier 5: Suggestive evidence of an association from a single data set**

•	Analysis of a single cohort/dataset provides genome-wide suggestive evidence for association.

•	All information is provided to support a robust analysis as defined below.

•	All information is available to support a robust association as defined below.

•	Analysis methods are well-documented and are supported by other works in the literature.

•	All required sources of evidence are provided, and the evidence is optimal.

**Tier 6: Limited evidence of an association.** There is some evidence of an association between this locus and AD or closely related phenotypes, but additional supporting evidence is needed to be confident the results are not due to chance, confounding factors, or bias. Criteria for this tier are:

•	Genome-wide suggestive or significant evidence for association.

•	Moderate limitations are present in the evidence for a robust signal.

•	Moderate limitations are present in the evidence for a robust association.

•	Moderate limitations are present in the evidence for a consistent association.

•	The strength of the association and/or methods used are such that there is only weak support for a locus.

**Tier 7: Insufficient evidence to determine whether an association exists.** While there is evidence of an association between this locus and AD or a closely related trait in at least one analysis, additional supporting evidence is needed to be confident the results are not a false-positive association due to genotyping/sequencing error, chance, confounding factors, or bias. There is reason to believe the result may not be a true association. Examples of associations that may fall in this tier include:

•	The variant may have evidence of genotyping or sequencing error.

•	The analysis model may not have adequately controlled for confounding factors, bias, etc.

•	The association was identified in a study with a small sample size (ex., N < 1,000).

•	Severe limitations, such as:

o	Evidence the association is a false positive.

o	Concerns about technical/analytic issues.

o	Data necessary to evaluate the robustness and consistence of the analysis are missing such that there is only weak or no support for a locus.

**OR**

•	No suggestive or genome-wide significant evidence for an association is provided.

The elements in Table A2 are used to define:

•	Variant definition

•	Significance

•	Consistent results

•	Robust results

•	Robust analys

| **Table A2. Evaluator decision tool details.** | | | |
| --- | --- | --- | --- |
|  |  | **Tier requirements** | **Notes** |
| Variant information | Reference genome | Must have |  |
|  | Chr:Pos:Ref | Must have | chr:pos with ref allele |
|  | Lead SNP identifier | Must have | rsID |
|  | Lead SNP position | Must have |  |
|  | Lead SNP risk allele | Must have | Reported by the publication |
|  | Can you identify the elevated or protective allele? | Must have | Reported by the publication |
|  | Lead SNP risk allele frequency | Extra | Overall AF on risk allele reported by publication |
|  | Lead SNP risk allele frequency – cases | Extra |  |
|  | Lead SNP risk allele frequency – controls | Extra |  |
|  | Lead SNP beta | Must have one of these |  |
|  | Lead SNP beta std error | Must have one of these |  |
|  | Lead SNP Z-score | Must have one of these |  |
|  | Lead SNP OR + C.I. | Must have one of these |  |
|  | lead SNP Bayes' Factor | Must have one of these |  |
|  | Lead SNP p-value | Must have |  |
| Significance | Genome-wide significance with *P* < 5 x 10^-08^ or Genome-wide suggestive with *P* < 1 x 10^-06^ | Must have | The minimum is "suggestive" for the data we are collecting |
| Consistent results | Forest plots | Must have one of these |  |
|  | Signed statistics for each separate cohort (OR, beta, *etc.*). Cohort is defined by authors. | Must have one of these |  |
|  | line plot (+/-) | Must have one of these |  |
|  | heterogeneity statistic | Must have one of these |  |
|  | phenotype match (AD, Dementia, or AD + Dementia). AD clin/path cohorts and Dementia cohorts are not considered “similar” cohorts for considering heterogeneity. | Must have an AD-related outcome. | AD-related outcome as defined in the inclusion criteria |
|  | Conclusion ranking for consistent associations | Must have | The overall rank for whether a result is consistent |
| Robust results | Was conditional analysis of close SNVs performed? | Desirable but not required – note in comments | If not present grade each 'close locus' separately but note no conditional analysis to determine independent loci |
|  | If performed, does conditional analysis concluded the locus is independent? | Desirable but not required – note in comments | If not present grade each 'close locus' separately but note no conditional analysis to determine independent loci |
|  | Locus Zoom or information from supporting nearby SNVs (required for novel common variant loci) | Must have for novel loci -supporting suggestive or significant SNVs in the region |  |
|  | Conclusion ranking for robust results | Must have | The overall rank for whether a result is robust |
| Robust analysis | Q-Q plot | Not required |  |
|  | inflation statistics | Must have |  |
|  | methodology (Novelty) | Must have | As part of the literature review, we removed papers that only used novel untested methods; if the paper has a novel untested method and an established method, we just review the established method. A novel method can be reviewed if it produces results consistent with those of more traditional methods. |
|  | methods - QC appropriate, not worried about batch effects, etc. | Must have |  |
|  | methods - model appropriate | Must have |  |
|  | significance thresholds are appropriate (genome-wide, acknowledges multiple testing) | Must have | Using fixed significance threshold of 5e-08 and suggestive threshold of 1e-06 |
|  | Population stratification corrected for (PCs, *etc*.) | Must have |  |
|  | Analysis reviewed | Must have | If a meta-analysis, focus on the meta-analysis results not individual stages. For reports where multiple models are tested, we will consider the primary analysis (if stated). If not stated, we will consider each model separately and note whether the significance was corrected for multiple testing. |
|  | Analysis type | Must have | Is the analysis of a single dataset or multiple datasets/meta-analysis |
|  | Conclusion ranking for robust analysis | Must have | The overall rank for whether an analysis is robust |

Phase 1 Locus/Gene Reports: Appendix 5

Gene/Locus	page in Appendix

*ABCA7*	p 19-24

*ABI3*	p 25-26

*ADAM10*	p 27-29

*APOE*	p 30-32

*APP*	p 33-34

*BIN1*	p 35-39

*CASP7*	p 40-42

*CD33*	p 43-45

*SCARA3 (CLU locus)*	p 46-47

*CR1*	p 48-51

*HLA-DRB5*	p 52-53

*IGHV1-67*	p 54-55

*MS4A4A/MS4A6A*	p 56-67

*PLCG2*	p 68-70

*PPARGC1A*	p 71-73

*PSEN1*	p 74-75

*PSEN2*	p 76-77

*RORA*	p 78-79

*SORL1*	p 80-82

*SPI1*	p 83-85

*TP53INP1*	p 86-88

*TREM2*	p 89-90

*ZNF423*	P 91-92

*ABCA7*

Locus – Data to collect

Gene: *ABCA7*

Evaluator: Kunkle

Date: 9/26/19

1. Significance
   a. discovery study^1^
i. study size (#cases/controls)

Stage 1: 6,688 cases and 13,685 controls

GWAS

Lead SNP: rs3764650

OR (CI): 1.22 (1.13-1.32)

P-value: 2.6 x 10^-7^

MAF: 0.10

ii. Ancestry

non-Hispanic white

iii. evaluation of population heterogeneity (yes/no/comments)

Yes in both Hollingsworth and Naj

iv. multiple test/model corrections (yes/no/comments)

logistic regression adjusting for country of origin and principal components

b. meta-analysis – discovery study

i. study size (#cases/controls)

Stage 2: 4,896 cases and 4,903 controls

Genotyping

OR (CI): 1.28 (1.14-1.44)

P-value: 1.9 x 10^-5^

Stage 3: 8,286 cases and 21,258 controls

Genotyping

OR (CI): 1.22 (1.13-1.32)

P-value: 2.9 x 10^-7^

Meta-analysis Stage 1+2+3: 19,870 cases and 39,846 controls

OR (CI): 1.23 (1.18-1.30)

P-value: 4.5 x 10^-17^

Meta-analysis Stage 1+ADGC: ?

OR (CI): 1.23 (1.17-1.28)

P-value: 5.0 x 10^-21^

ii. evaluation of population heterogeneity (yes/no/comments)

Yes

iii. multiple test/model corrections (yes/no/comments)

Inverse-variance weighted fixed effects meta-analysis

c. replication studies, common variants
- independent investigators and data – RARE^3^
  - Study size (#cases/controls): 1,968 cases and 3,928 controls
  - Ancestry: African-Americans
  - Lead SNP: rs115550680
  - MAF: 0.09 cases, 0.06 controls
  - OR (CI): 1.79 (1.47 – 2.12)
  - P-value: 2.2 x 10^-9^
  - evaluation of population heterogeneity (yes/no/comments)

Yes

- multiple test/model corrections (yes/no/comments)

Logistic regression adjusting for age, sex, PCs and APOE

- larger study with added samples^4, 5, 6,7,8^

Lambert et al. 2011

- Study size (#cases/controls):
- Ancestry: non-Hispanic white
- Lead SNP:
- MAF:
- OR (CI):
- P-value:
- evaluation of population heterogeneity (yes/no/comments)
- multiple test/model corrections (yes/no/comments)

Jun et al. 2017

- Study size (#cases/controls):
- Ancestry: non-Hispanic White, Japanese, African-American, Israeli-Arab
- Lead SNP:
- MAF:
- OR (CI):
- P-value:
- evaluation of population heterogeneity (yes/no/comments)
- multiple test/model corrections (yes/no/comments)

Kunkle et al. 2019

- Study size (#cases/controls):
- Ancestry: non-Hispanic white
- Lead SNP:
- MAF:
- OR (CI):
- P-value:
- evaluation of population heterogeneity (yes/no/comments)
- multiple test/model corrections (yes/no/comments)

Jansen et al. 2019

- Study size (#cases/controls):
- Ancestry: non-Hispanic white
- Lead SNP:
- MAF:
- OR (CI):
- P-value:
- evaluation of population heterogeneity (yes/no/comments)
- multiple test/model corrections (yes/no/comments)

Marioni et al. 2019

- Study size (#cases/controls):
- Ancestry: non-Hispanic white
- Lead SNP:
- MAF:
- OR (CI):
- P-value:
- evaluation of population heterogeneity (yes/no/comments)
- multiple test/model corrections (yes/no/comments)
d. replication studies – evidence
- signal direction
- other ethnic groups study size (#cases/controls)
- evaluation of population heterogeneity (yes/no/comments)
- multiple test/model corrections (yes/no/comments)

2. Pleiotropy
   a. AD-related phenotype (*e.g*. CSF biomarker association)

Dementia (UK Biobank – see Jansen et al. 2019 above)

b. Non-AD phenotype (*e.g*. coronary artery disease, *APOE*)

3. Recommended tier: tier 1
4. Supporting references

1. Hollingsworth et al. Common variants at ABCA7, MS4A6A/MS4A4E, EPHA1, CD33 and CD2AP are associated with Alzheimer’s disease. *Nat Genet*. 2011 May; 43(5): 429-35.

2. Naj et al. Common variants at MS4A4/MS4A6E, CD2AP, CD33 and EPHA1 are associated with late-onset Alzheimer’s disease. *Nat Genet*. 2011 May; 43(5): 436-41.

3. Reitz et al. Variants in the ATP-binding cassette transporter (ABCA7), apolipoprotein E ϵ4,and the risk of late-onset Alzheimer disease in African Americans. 2013 *JAMA* Apr 10; 309(14):1483-92.

4. Lambert et al.

5. Jun et al.

6. Kunkle et al.

7. Jansen et al.

8. Marioni et al.

**PubMed search ‘ABCA7 Alzheimer’ returns 128 articles**

- GWAS (N=4)
  1. Logue et al. A comprehensive genetic association study of Alzheimer disease in African Americans. Arch Neurol. 2011 Dec;68(12):1569-79.
  2. Kamboh et al. Genome-wide association study of Alzheimer’s disease. Transl Psychiatry. 2012 May;2:e117.
  3. Variants in the ATP-binding cassette transporter (ABCA7), apolipoprotein E ϵ4,and the risk of late-onset Alzheimer disease in African Americans. JAMA. 2013 Apr;309(14):1483-92.
  4. Chen et al. A multi-ancestral genome-wide exome array study of Alzheimer disease, frontotemporal dementia, and progressive supranuclear palsy. JAMA Neurol. 2015 Apr;72(4):414-22.
- GWAS Meta-analyses (N=5)
  1. Hollingsworth et al. Common variants at ABCA7, MS4A6A/MS4A4E, EPHA1, CD33 and CD2AP are associated with Alzheimer’s disease. Nat Genet. 2011 May;43(5):429-35.
  2. Lambert et al.
  3. Kunkle et al.
  4. Marioni et al.
  5. Jansen et al.
- Genomewide Exome/Genome Sequencing association (N= )
  1. Bis et al. Whole exome sequencing study identifies novel rare and common Alzheimer’s associated variants involved in immune response and transcriptional regulation. Mol Psychiatry. 2018 Aug;
  2. N’Songo A et al. African American exome sequencing identifies potential risk variants at Alzheimer disease loci. Neurol Genet. 2017 Apr;3(2):e141.
- Targeted/candidate gene sequencing AA association (N=9)
  1. Steinberg S et al. Loss-of-function variants in ABCA7 confer risk of Alzheimer disease. Nat Genet. 2015 May;47(5):445-7.
  2. Cuyvers et al. Mutations in ABCA7 in a Belgian cohort of Alzheimer's disease patients: a targeted resequencing study. Lancet Neurol. 2015 Aug;14(8):814-22.
  3. Vardarajan et al. Rare coding mutations identified by sequencing of Alzheimer disease genome-wide association studies loci. Ann Neurol. 2015 Sep;78(3):487-98.
  4. Del-Aguila JL. Role of ABCA7 loss-of-function variant in Alzheimer’s disease: a replication study in European-Americans. Alzheimers Res Ther. 2015 Dec;7(1):73.
  5. Cukier et al. ABCA7 frameshift deletion associated with Alzheimer disease in African Americans. Neurol Genet. 2016 May;2(3):e79.
  6. Le Guennec et al. ABCA7 rare variants and Alzheimer disease risk. Neuology. 2016 Jun;86(23):2134-7.
  7. Kunkle et al. Targeted sequencing of ABCA7 identifies splicing, stop-gain and intronic risk variants for Alzheimer disease. Neurosci Lett. 2017 May;649:124-129.
  8. De Roeck et al. Deleterious ABCA7 mutations and transcript rescue mechanisms in early onset Alzheimer’s disease. Acta Neuropath. 2017 Sep;134(3):475-87.
  9. Bellenguez C et al. Contribution to Alzheimer's disease risk of rare variants in TREM2, SORL1, and ABCA7 in 1779 cases and 1273 controls. Neurobiol Aging. 2017 Nov;59:220.e1-220.e9.
- Candidate Gene genotyping association (N=15)
  1. Shi et al. Genetic variants influencing human aging from late-onset Alzheimer’s disease (LOAD) genome-wide association studies (GWAS). Neurobiol Aging. 2012 Aug;33(8):e5-18.
  2. Engelman et al. Interaction between two cholesterol metabolism genes influences memory: findings from the Wisconsin Registry for Alzheimer's Prevention. J Alzheimers Dis. 2013;36(4):749-57.
  3. Cascorbi et al. Association of ATP-binding cassette transporter variants with the risk of Alzheimer’s disease. Pharmacogenomics. 2013 Apr;14(5):485-94.
  4. Chung et al. Association of GWAS top hits with late-onset Alzheimer disease in Korean population. 2013 Jul-Sep;27(3):250-7.
  5. Tan et al. Association of GWAS-linked loci with late-onset Alzheimer’s disease in a northern Han Chinese population. Alzheimers Dement. 2013 Sep;9(5):546-53.
  6. Yang et al. Association study of ABCA7 and NPC1 polymorphisms with Alzheimer’s disease in Chinese Han ethnic population. Psychiatr Genet. 2013 Dec;23(6):268.
  7. Liu et al. A complex association of ABCA7 genotypes with sporadic Alzheimer disease in Chinese Han population. Alzheimer Dis Assoc Disord. 2014 Apr-Jun;28(2):141-4.
  8. Liao et al. ABCA7 gene and the risk of Alzheimer’s disease in Han Chinese in Taiwan. Neurobiol Aging. 2014 Oct;35(10):2423.e702423.e13.
  9. Moreno DJ et al. Association of GWAS top genes with late-onset Alzheimer’s disease in Colombian population. Am J Alzheimers Dis Other Demen. 2017 Feb;23(1):27-35.
  10. Ma et al. Meta-analysis of the association between variants in ABCA7 and Alzheimer’s disease. J Alzheimers Dis. 2018:63(4):1261-1267.
  11. Almeida et al. Updated meta-analysis of BIN1, CR1, MS4A6A, CLU and ABCA7 variants in Alzheimer’s disease. 2018 Mar;64(3):471-77.
  12. De Roeck A. An intronic VNTR affects splicing of ABCA7 and increases risk of Alzheimer’s disease. Acta Neuropath. 2018 Jun;135(6):827-37.
- Familial
  1. Rare ABCA7 variants in 2 German families with Alzheimer disease. Neurol Genet. 2018 Mar;4(2):e224.
- Pleiotropy (AD-related phenotype) association (N=24)
  1. Karch CM et al. Expression of novel Alzheimer’s disease risk genes in control and Alzheimer’s disease brains. PLoS One. 2012;7(1):e50976.
  2. Allen et al. Novel late-onset Alzheimer disease loci variants associate with gene expression. Neurology. 2012 Jul;79(3):221-8.
  3. Shulman JM et al. Genetic susceptibility for Alzheimer disease neuritic plaque pathology. JAMA Neurol. 2013 Sep;70(9):1150-7.
  4. CR1, ABCA7, and APOE genes affect the features of cognitive impairment in Alzheimer’s disease. J Neurol Sci. 2014 Apr;339(1-2):91-6.
  5. Carrasquillo et al. Late-onset Alzheimer disease genetic variants in posterior corticol atrophy and posterior AD. Neurology. 2014 Apr;82(16):1455-62.
  6. Beecham et al. Genome-wide association meta-analysis of neuropathologic features of Alzheimer’s disease and related dementias. PLoS Genet. 2014 Sep;10(9):e1004606.
  7. Carrasquillo et al. Late-onset Alzheimer’s risk variants in memory decline, incident mild cognitive impairment, and Alzheimer’s disease. Neurobiol Aging. 2015 Jan;36(1):60-7.
  8. Yu et al. Association of Brain DNA methylation in SORL1, ABCA7, HLA-DRB5, SLC24A4, and BIN1 with pathological diagnosis of Alzheimer disease. JAMA Neurol. 2015 Jan;72(1):15-24.
  9. Satoh et al. ATP-binding cassette transporter A7 (ABCA7) loss of function alters Alzheimer amyloid processing. J Biol Chem. 2015 Oct;290(40):24152-65.
- Pleiotropy (Non-AD phenotype) association (N= )
- Mouse (N=2)
- Clinical (N=1)
  1. Van den Bossche et al. Phenotypic characteristic of Alzheimer patients carrying an ABCA7 mutation. Neurology. 2016 Jun;86(23):2126-33.

*ABI3*

Locus – Data to collect

Locus: ABI3

Evaluator:

Schellenberg

Date: 9/24/2019

1. Significance
a. discovery study^1^
i. study size (#cases/controls)

- Stage 1
- 16,097 cases/18,077 controls
- Exome chip genotyping
- P = 2.16 x 10^-5^; OR = 1.42
- MAF cases 0.013; MAF controls 0.010

ii. evaluation of population heterogeneity (yes/no/comments)

- Yes

iii. multiple test/model corrections (yes/no/comments)

- Single model

b. meta-analysis – discovery study^1^
iv. study size (#cases/controls):

- Stage 2
- De novo genotyping and imputation, 43 variants
- 14,041 cases/ 21,921 controls
- P = 8.37 x 10^-4^; OR = 1.41
- MAF cases 0.010, MAF controls 0.008
- Stage 3
- Array genotypes used for imputation (some validated against exome chip data)
- 6,652 cases/8,345 controls
- P = 1.75 x 10^-2^; OR = 1.58
- MAF cases 0.010, MAF controls 0.008
- Meta-analyses(stages 1-3)
- P = 4.56 x 10-^10^; OR = 1.43
- MAF cases 0.011, MAF controls 0.008

v. evaluation of population heterogeneity (yes/no/comments)

- yes

vi. multiple test/model corrections (yes/no/comments)

- single model

c. replication studies – independent investigators, common variant data^2^

- Phase 1 (overlaps with IGAP/ADSP data) AD phenotype
- P = 7.29 x 10^-5^
- Phase 2 (UK Biobank proxy cases, dementia phenotype)
- P = 6.80 x 10^-6^
- Phase 3 (overall)
- P = 1.87 x 10^-8^
- MAF = 0.473
- evaluation of population heterogeneity (yes/no/comments)
- yes
- multiple test/model corrections (yes/no/comments)
- single model
- other ethnic groups study size (#cases/controls)
- larger study with added samples (see above^2^
  2. Pleiotropy
  c. AD-related phenotype (*e.g*. CSF biomarker association)

i. Dementia (UK Biobank – see above)^2^
   d. Non-AD phenotype (*e.g*. coronary artery disease, *APOE*)
      1. Recommended tier (locus): tier 2? (maybe tier 1?)

Comments: Original discovery was using a rare-variant approach. Replication has two caveats: 1) the discovery variants is rare (MAF = 0.008 in controls) while the replication variant is common (MAF = 0.473); 2) The replication is uses bot AD and dementia phenotypes..

*ADAM10*

Locus – Data to collect

Locus: ADAM10

Evaluator:	Schellenberg

Date:	9/24/2019

1. Significance
a. discovery study^1^ Candidate gene (*ADAM10*) study

- Stage 1 association – NIMH families
- 436 multiplex families, 1,439 samples
- Targeted genotyping (ADMA10 only)
- P = -0.003 (rs2305421) (n = 103)
- MAF overall = 0.123
- APOE stratified (e4 positive) P = 0.0005 (n = 86)
- Comment – number of NIMH families reported in the association study was smaller than the total NIMH families available
- Stage 2 association - Consortium on Alzheimer’s disease (CAG)
- 217 families, 489 samples, discordant sibpairs
- P = 0.732 (rs2305421) (n = 19)
- APOE stratified (e4 positive) P = 0.538 (n = 19)
- Stage 3 association - Combined: NIMH + CAG
- 150 families
- P = -0.008 (rs2305421) (n = 150)
- APOE stratified (e4 positive) P = 0.0007 (n = 105 families)
- Targeted sequencing of ADAM10
- Q170H and R181G in 5 families (n = 32 NIMH families sequenced)
- Screened all NIMH families – 3 more families found with carriers
- AD mutation carriers onset ranges from 63-80 years
- Unaffected carriers observed, age-at-last exam 63-83 years.
- In each family with an AD carrier, there we AD case non-carriers
- evaluation of population heterogeneity (yes/no/comments)
- no
- multiple test/model corrections (yes/no/comments)
- Single model

b. Large scale GWAS studies – Kunkle et al.^2^
i. Stage 1 + 2; n= 30344 cases, 52,427 controls

- rs593742, P = 1.3 x 10^-7^, OR = 0.91 (0.91-0.96) MAF cases + controls = 0.295

ii. Stage 3A; 4,930 cases, 6,736 controls

- rs593742, P = 1.5 x 10^-2^, OR = 0.93 (0.85-0.98)

iii. Overall (n = 35,274 cases, 59, 163 controls

- rs593742, P = 6.8 x 10^-9^, OR = 0.93 (0.91-095)

iv. evaluation of population heterogeneity (yes/no/comments)

- yes

v. multiple test/model corrections (yes/no/comments)

- single model

vi. Comment – Locus zoom shows broad signal over ADAM10 but also over FAM63B (Mindy2) and other transcripts
c. Large-scale GWAS study - Jansen et al. ^3^
i. Phase 1: PGC-ALZ (n = 17,477), IGAP (n = 54,162), ADSP (n = 7,506)

- rs442495P = 3.09 x 10^-4^,

ii. AD by proxy (UK Biobank proxy cases, dementia phenotype)(n = 376,113)

- P = 2.65 x 10-7

iii. Overall (n = 455,258)

- P = 1.31 x 10^-9^
- MAF = 0.320

iv. Replication – DeCode (n = 6,593 cases, 180,882)

- rs442495, P = 0.121, OR = 0.964
- rs650366, P = 0.019, OR = 0.947 (strongest SNP at the same locus)

v. Comment – there is substantial overlap between this paper, Kunkle et al, and Marioni et al. Locus zoom plot looks the same as in Kunkle et al.
vi. evaluation of population heterogeneity (yes/no/comments)

- yes

vii. multiple test/model corrections (yes/no/comments)

- single model

viii. other ethnic groups study size (#cases/controls) Pleiotropy
d. Large-scale GWAS – Maroni et al.^4^

- UK Biobank proxy GWAS + IGAP summary statistics
- rs593742, 6.2 x 10^-11^, OR 1.06 (1.04 – 1.07)
- Note effect direction is not consistent with Kunkle et al or Jansen et al.

e. Large-scale GWAS – de Rojas et al.^5^

- Original discovery was 12,386 GR@ACE; 82,771 IGAP, 314,278 UKB
- Follow-up of genome-wide loci: EADB, 33,495; PG-Alz, 17,537; GR@ACE, 1,202; N x C, 1,078; ADDN, 637; Sidney NAS, 258; AD and GBCS, 3,981.
- ADAM10, rs593742, P = 3.20 x ^-15^, OR = 1.08[1.06-1.10]
- Functional evidence. In transgenic mice, ADAM10 mutations influence the α-secretase and shift Aβ cleavage to the BACE1 cleavage site and increases Aβ levels and plaque load^6^. The Q170H and R181G mutations reduce ADAM10 α-secretase activity
  2. Recommended tier (locus): tier 1; Gene – tier 2

Members of the ADAM family are cell surface proteins with a unique structure possessing both potential adhesion and protease domains. This gene encodes and ADAM family member that cleaves many proteins including TNF-alpha and E-cadherin. Alternate splicing results in multiple transcript variants encoding different proteins that may undergo similar processing. [provided by RefSeq, Feb 2016].

**Gencode Transcript:** ENST00000561288.1
**Gencode Gene:** ENSG00000137845.15
**Transcript (Including UTRs)**
   **Position:** hg38 chr15:58,597,164-58,749,707 **Size:** 152,544 **Total Exon Count:** 2 **Strand:** -
**Coding Region**
   **Position:** hg38 chr15:58,597,457-58,749,534 **Size:** 152,078 **Coding Exon Count:** 2

*APOE*

Locus – Data to collect

Locus: *APOE*

Evaluator:

Naj

Date:

10/2/2019

1. Significance
a. discovery study{Pericak-Vance, 1991 #132}
   i. study size (#cases/controls)
- Affected Pedigree Member (APM)/ LOD-based Linkage Analysis
- 293 pedigree members (87 affecteds) in 32 Familial Alzheimer Disease (FAD) Families with affecteds with age-at-onset > 60
- Microsatellite (Single Tandem Repeat (STR)) panel with 36 markers
- D19S13; LOD (overall) = 2.20; LOD (affected only) = 4.38
  ii. evaluation of population heterogeneity (yes/no/comments)
- Yes (not ethnic heterogeneity, heterogeneity of the signal across FAD families)

iii.	multiple test/model corrections (yes/no/comments)

- Two statistical models (APM/LOD), no multiple correction

b. meta-analysis – discovery study
iv. study size (#cases/controls):

- Kunkle et al. 2019 (Non-Hispanic Whites)
- GWAS meta-analysis
- 21,982 cases/ 41,944 controls
- *APOE* SNP rs429358; chr19:45411941; A1/A2: T/C; MAF(A2) = 0.216; OR (95% CI): 3.32 (3.20, 3.45); *P* = 1.2 × 10^−881^
- Jansen et al. 2019 (Non-Hispanic Whites)
- GWAS meta-analysis
- Phase 1: 24,087 cases/ 55,058 controls; Phase 2: 376,113 cases/328,320 controls
- Phase 1: *APOE* SNP rs41289512; chr19:45351516; *P* = 2.70 × 10^−194^
- Phase 2: *APOE* SNP rs75627662; chr19:45413576; *P* = 9.51 × 10^−296^
- Phase 3: *APOE* SNP rs41289512; chr19:45351516; A1/A2: C/T; MAF(A2) = 0.375; *P* = 5.79 × 10^−276^

v. evaluation of population heterogeneity (yes/no/comments)

- yes- heterogeneity between individual datasets

vi. multiple test/model corrections (yes/no/comments)

- yes, single model, empirical genome-wide significance threshold *P* = 5 × 10^-8^

c. replication studies

- independent investigators and data
- Discovery datasets identified above encompass all currently available Non-Hispanic White sample sets with available *APOE* genotype data
- Contributing datasets have similar size and direction of effect
- evaluation of population heterogeneity (yes/no/comments); NA
- multiple test/model corrections (yes/no/comments); NA
- other ethnic groups study size (#cases/controls)
- Farrer et al. 1997 (PMID: 9343467)
- Caucasian: ε2/ε4 vs. ε3/ε3 (OR=2.6, 95% Cl=1.6-4.0); ε3/ε4 vs. ε3/ε3 (OR=3.2, 95% Cl=2.8-3.8); ε4/ε4 vs. ε3/ε3 (OR=14.9, 95% CI=10.8-20.6); ε2/ε2 vs. ε3/ε3 (OR=0.6, 95% Cl=0.2-2.0); ε2/ε3 vs. ε3/ε3 (OR=0.6, 95% Cl=0.5-0.8)
- Japanese: ε2/ε4 vs. ε3/ε3 (OR=2.4, 95% Cl=0.4-15.4); ε3/ε4 vs. ε3/ε3 (OR=5.6, 95% Cl=3.9-8.0); ε4/ε4 vs. ε3/ε3 (OR=33.1, 95% Cl=13.6-80.5); ε2/ε2 vs. ε3/ε3 (OR=1.1, 95% Cl=0.1-17.2); ε2/ε3 vs. ε3/ε3 (OR=0.9, 95% Cl=0.4-2.5)
- African Americans: ε2/ε4 vs. ε3/ε3 (OR=1.8, 95% Cl=0.4-8.1); ε3/ε4 vs. ε3/ε3 (OR=1.1, 95% Cl=0.7-1.8); ε4/ε4 vs. ε3/ε3 (OR=5.7, 95% Cl=2.3-14.1); ε2/ε2 vs. ε3/ε3 (OR=2.4, 95% Cl=0.3-22.7); ε2/ε3 vs. ε3/ε3 (OR=0.6, 95% Cl=0.4-1.7)
- Hispanics: ε2/ε4 vs. ε3/ε3 (OR=3.2, 95% Cl=0.9-11.6); ε3/ε4 vs. ε3/ε3 (OR=2.2, 95% Cl=1.3-3.4); ε4/ε4 vs. ε3/ε3 (OR=2.2, 95% Cl=0.7-67); ε2/ε2 vs. ε3/ε3 (OR=2.6, 95% Cl=0.2-33.3); ε2/ε3 vs. ε3/ε3 (OR=0.6, 95% Cl=0.3-1.3)
- General findings:
  - The ε2/ε3 genotype appears equally protective across ethnic groups.
  - Among Caucasians, *APOE* genotype distributions are similar in groups of patients with AD whose diagnoses were determined clinically or by autopsy
  - APOE ε4 effect is evident at all ages between 40 and 90 years but diminishes after age 70 years and that the risk of AD associated with a given genotype varies with sex
- larger study with added samples
- Transethnic meta-analysis (Jun et al. 2017) (PMID: 28183528)
- Overall: rs283811-G, OR=2.20, 95%CI=2.12-2.29, *P*=7×10^-300^
  2. Pleiotropy

c. AD-related phenotype (*e.g*. CSF biomarker association)
i. AD Age-at-onset (Corder et al. 1993)

- 42 families
- *APOE* ε2/ε3/ε4 dosage effect on AAO
- Mean AAO (0 ε4 allele) = 84.3y; Mean AAO (1 ε4 allele) = 75.5y; Mean AAO (2 ε4 alleles) = 68.8 y
- Higher risk corresponds with lower AAO

ii. CSF Biomarkers

- Kim et al. 2011 (PMID: 21123754)
- rs429358 (19:50103781) association with Aβ_1–42_, p-tau_181p_, t-tau/Aβ_1–42_
- Additional citation PMIDs: 25027320, 28641921, 28247064, 30319691

iii. Brain Imaging

- Meda et al. 2012 (PMID: 2245343)
  - Combined association across imaging from 94 different brain regions- unnamed SNPs in *APOE* (ε4; *P*=6.6E-16; ε3; *P*=3.6E-09) and *TOMM40* (*P*=7.25E-08)
- Additional citation PMIDs: 20100581, 29860282

iv. Cognitive Decline/Memory Loss

- Raj et al. 2017 (PMID: 28078323)
  - APOE SNP rs429538 (P=1.92E-14) associated with cognitive decline in African Americans
- Additional citation PMIDs: 24468470, 30319691, 25648963

v. Posterior Cortical Atrophy variant of AD

- PMID: 26993346

vi. Brain Amyloid Deposition

- PMID: 30361487, 23419831

vii. Dementia with Lewy Bodies
     a. PMID: 29263008, 31065058, 25188341
viii. Hippocampal Atrophy
      a. PMID: 29263008, 31065058, 25188341
         3. Non-AD phenotype (*e.g*. coronary artery disease, *APOE*)

- Cardiometabolic traits:
  - Lipids (PMID: 21909109, 20686565, 28270201, 24097068, 23726366, 18262040, 26582766, 19060911, 29403010, 30275531, 29084231)
  - Blood Pressure/Hypertension (PMID: 30224653, 30578418)
  - Diabetes (PMID: 28869590)
  - Subclinical Atherosclerosis (PMID: 21909108, 30361487)
  - Cardiovascular disease phenotypes (PMID: 26343387, 28714975)
- Age-related macular degeneration (AMD) (PMID: 23455636, 23326517, 29227965)
- Longevity (PMID: 21740922, 21418511, 27029810, 31413261)
  a. Recommended tier (locus): tier 1
  b. Recommended tier (gene): tier 1

*APP*

Genes – rare coding variants

Locus – APP

Evaluator: **Goate**

Date: **9/25/2019**

A. Primary evidence
1. Non-synonymous SNV (CADD > 20?)

Multiple non-synonymous variants that segregate with disease in ADAD. Initial report described APPV717I in two families (Goate et al., *Nature*. 1991 Feb 21;349(6311):704-6).

2. Altered function SNV (stop gain, stop loss, splicing)

None

3. Structural variants – disrupt expression

Multiple different structural variants resulting in duplication of APP reported (add refs)

4. Significant association evidence with AD

Multiple mutations with evidence of segregation with AD in ADAD kindreds. <https://www.alzforum.org/mutations/app>

5. Replication in another population

APPV717I has been reported in more than 30 families from United States, England, Japan, Thailand, Germany, France, Italy, Australia, Belgium, Iran, and China

6. Gene expressed in an AD-relevant tissue/cell type

APP is broadly expressed in the body including all cell types in the brain

B. Supporting evidence
1. Multiple rare variants significantly associated with AD

More than 30 variants reported to be associated with AD. <https://www.alzforum.org/mutations/search?genes%5B%5D=348&diseases%5B%5D=145&diseases%5B%5D=147&keywords-entry=&keywords=#results>

2. Common variants associated with the same gene

unclear

3. Pleiotropy

Variants in APP are associated with AD, CAA or stroke

4. Rare variants in the same gene in another ethnic group

Yes

5. Co-segregation in families

Yes

6. Other?

Rare protective variant A673T, associated with healthy longevity, reported in people of Scandinavian ancestry.

Jonsson T, Atwal JK, Steinberg S, Snaedal J, Jonsson PV, Bjornsson S, Stefansson H, Sulem P, Gudbjartsson D, Maloney J, Hoyte K, Gustafson A, Liu Y, Lu Y, Bhangale T, Graham RR, Huttenlocher J, Bjornsdottir G, Andreassen OA, Jönsson EG, Palotie A, Behrens TW, Magnusson OT, Kong A, Thorsteinsdottir U, Watts RJ, Stefansson K. [A mutation in APP protects against Alzheimer's disease and age-related cognitive decline](https://www.alzforum.org/papers/mutation-app-protects-against-alzheimers-disease-and-age-related-cognitive-decline). *Nature*. 2012 Aug 2;488(7409):96-9. [PubMed](http://www.ncbi.nlm.nih.gov/pubmed/22801501).

Animal and cell models carrying APP mutations show altered APP metabolism and animal models show age dependent Aß deposition.

*BIN1*

Locus – Data to collect

Gene: *BIN1*

Evaluator: Kunkle

Date: 11/14/19

1. Significance
   a. discovery study^1^
   i. study size (#cases/controls)

Stage 1: 3,006 cases and 14,642 controls (CHARGE, TGRI, Mayo)

GWAS array

Lead SNP: rs744373

OR (95% CI): 1.13 (1.06 -1.21)

P-value: 4.93 x 10^-4^

MAF: 0.291

ii. Ancestry

non-Hispanic white

iii. evaluation of population heterogeneity (yes/no/comments)

Yes

iv. multiple test/model corrections (yes/no/comments)

Cox proportional hazards and logistic regression models adjusting for age, sex and principal components. CHS also adjusted for study site, and FHS accounted for familial relationships.

b. meta-analysis – discovery study

v. study size (#cases/controls)

Stage 2: 2,032 cases and 5,328 controls (EADI)

GWAS array

OR (95% CI): 1.14 (1.08 - 1.20)

P-value: 1.02 x 10^-5^

Meta-analysis Stage 1+2+3: 8,371 cases and 26,965 controls (CHARGE+TGRI+Mayo+EADI+GERAD)

Stage 3: 3,333 cases and 6,995 controls (GERAD)

GWAS array

OR (95% CI): 1.13 (1.06 - 1.21)

P-value: 2.9 x 10^-11^

Replication Stage: 1,140 cases and 1,209 controls (Fundacio ACE)

OR (95% CI): 1.17 (1.03 - 1.33)

P-value: 0.02

vi. evaluation of population heterogeneity (yes/no/comments)

Yes

vii. multiple test/model corrections (yes/no/comments)

Inverse-variance weighted fixed effects meta-analysis

c. replication studies, common variants
- independent investigators and data – RARE
  a. Moreno et al. 2017^2^
  b. Study size (#cases/controls): 280 cases and 357 controls
  c. Ancestry: Hispanic – Colombia
  d. Lead SNP: rs3764650
  e. MAF: 0.31 in cases, 0.27 in controls
  f. OR (95% CI): 1.42
  g. P-value: 0.015
  h. evaluation of population heterogeneity (yes/no/comments):
  i. multiple test/model corrections (yes/no/comments):
  j. Reitz et al. 2013 (GENE-BASED)^3^
  k. Study size (#cases/controls): 280 cases and 357 controls
  l. Ancestry: African-American
  m. Most-significant SNP: rs55636820
  n. MAF: 0.02
  o. Gene-based P-value: 7 x 10^-4^
  p. evaluation of population heterogeneity (yes/no/comments): yes
  q. multiple test/model corrections (yes/no/comments): age, sex, APOE and PC
- larger study with added samples^4,5,6,7,8,9,10^

Naj et al. 2011

a. Study size (#cases/controls): 11,840 cases and 10,931 controls (ADGC)
b. Ancestry: non-Hispanic white
c. Lead SNP: rs7561528
d. MAF: 0.35
e. OR (95% CI): 1.17 (1.13 – 1.22)
f. P-value: 4.0 x 10^-14^
g. evaluation of population heterogeneity (yes/no/comments): yes
h. multiple test/model corrections (yes/no/comments): yes

Hollingworth et al. 2011

i. Study size (#cases/controls): 19,870 cases and 39,846 controls (GERAD, ADNI, TGEN, Mayo, EADI, CHARGE)
j. Ancestry: non-Hispanic white
k. Lead SNP: rs744373
l. MAF: 0.29
m. OR (95% CI): 1.17 (1.12 – 1.21)
n. P-value: 2.6 x 10^-14^
o. evaluation of population heterogeneity (yes/no/comments): yes
p. multiple test/model corrections (yes/no/comments): yes

Lambert et al. 2013

q. Study size (#cases/controls): 25,580 cases and 48,466 controls (EADI, ADGC, CHARGE, GERAD)
r. Ancestry: non-Hispanic white
s. Lead SNP: rs6733839
t. MAF: 0.409
u. OR (95% CI): 1.22 (1.18 – 1.25)
v. P-value: 6.9 x 10^-44^
w. evaluation of population heterogeneity (yes/no/comments): yes
x. multiple test/model corrections (yes/no/comments): yes

Jun et al. 2017

y. Study size (#cases/controls): 26,320 non-Hispanic White, 4,983 African-American, 1,845 Japanese, 115 Israeli-Arab
z. Ancestry: non-Hispanic White, Japanese, African-American, Israeli-Arab
aa. Lead SNP: rs6733839
bb. MAF: NHW=0.42, AA=0.38, JPN=0.33, IA=0.38
cc. OR (95% CI): ALL=1.17 (1.13-1.22); NHW=1.19 (1.14-1.23), AA=1.09 (0.99-1.21), JPN=1.19 (0.99-1.41), IA=0.92 (0.54-1.57)
dd. P-value: ALL=3.3 x 10^-17^; NHW=1.2x10^-16^, AA=0.08, JPN=0.05, IA=0.77
ee. evaluation of population heterogeneity (yes/no/comments): yes
ff. multiple test/model corrections (yes/no/comments): yes

Kunkle et al. 2019

gg. Study size (#cases/controls): 30,344 cases and 52,427 controls (ADGC, EADI, CHARGE, GERAD)
hh. Ancestry: non-Hispanic white
ii. Lead SNP: rs6733839
jj. MAF: 0.407
kk. OR (95% CI): 1.20 (1.17 – 1.23)
ll. P-value: 2.1 x 10^-44^
mm. evaluation of population heterogeneity (yes/no/comments): yes
nn. multiple test/model corrections (yes/no/comments): yes

Jansen et al. 2019

oo. Study size (#cases/controls): 71,880 cases and 383,378 controls (ADSP, IGAP, PGC-ALZ, UKBB)
pp. Ancestry: non-Hispanic white
qq. Lead SNP: rs4663105
rr. MAF: 0.415
ss. OR (CI): not available
tt. P-value: 3.38 x 10^-44^
uu. evaluation of population heterogeneity (yes/no/comments): yes
vv. multiple test/model corrections (yes/no/comments): yes

Marioni et al. 2019

ww. Study size (#cases/controls): 67,614 cases and 320,710 controls (UK Biobank + IGAP)
xx. Ancestry: non-Hispanic white
yy. Lead SNP: rs6733839
zz. MAF:
aaa. OR (CI):
bbb. P-value: 2.4 x 10^-69^
ccc. evaluation of population heterogeneity (yes/no/comments): yes
ddd. multiple test/model corrections (yes/no/comments): yes
b. replication studies, rare studies
c. replication studies – evidence
- signal direction: consistent across studies
- other ethnic groups study size (#cases/controls): see above under replication studies
- evaluation of population heterogeneity (yes/no/comments)
- multiple test/model corrections (yes/no/comments)
  i. Pleiotropy
  d. AD-related phenotype (*e.g*. CSF biomarker association)

Dementia (UK Biobank – see Jansen et al. 2019 and Marioni et al. 2019 above), Neurofibrillary tangles (Beecham et al. 2014)^11^, neuritic plaques and neurofibrillary tangles (Chung et a. 2018)^12^

e. Non-AD phenotype (*e.g*. coronary artery disease, *APOE*)

Creatine kinase levels? (1 x 10^-9^), acute myeloid leukemia? (9 x 10^-16^), prostate carcinoma? (8 x 10^-6^), blood protein levels? (1 x 10^-7^)

i. Co-segregation in families
- Vardarajan 2015^13^ significant rare coding variant and also segregated in 2 of 6 families
- Vardarajan 2018^14^ significant gene-based test (P=0.0098)
  ii. Recommended tier (locus): tier 1
  iii. Recommended tier (gene): tier 1? *see functional evidence below
  iv. Supporting references

Functional evidence:

Chapius et al. Increased expression of BIN1 mediates Alzheimer genetic risk by modulating tau pathology. *Mol Psychiatry*. 2013 Nov;18(11):1225-34.

AD transcript levels increased in AD brains; identified novel 3 bp insertion allele ~28 kb upstream of *BIN1* which increased (1) transcriptional activity in vitro, (2) BIN1 expression levels in human brain, and (3) AD risk in three independent case-control cohorts. Also showed *Drosophila* BIN1 ortholog Amph suppressed tau-mediated neurotoxicity and that the 3bp insertion was associated with tau but not amyloid loads in AD brains.

Novikova et al. Integration of Alzheimer’s disease genetics and myeloid cell genomics identifies novel causal variants, regulatory elements, genes and pathways. *BioRxiv*. 2019.

Identifies candidate risk enhancer with *BIN1* as target

Kunkle et al. Kunkle et al. Genetic meta-analysis of diagnosed Alzheimer’s disease identifies new risk loci and implicates A-beta, tau, immunity and lipid processing. Nat Genet. 2019. 51:414-30.

*BIN1* nominated as top prioritized risk gene in locus based on combined evidence from (1) deleterious coding, LOF or splicing variants in the gene; (2) significant gene-based tests; (3) expression in a tissue relevant to Alzheimer’s disease (astrocytes, neurons, microglia/macrophages, oligodendrocytes); (4) a HuMi microglial-enriched gene; (5) having an eQTL effect on the gene in any tissue, in Alzheimer’s disease–relevant tissue, and/or a co-localized eQTL; (6) being involved in a biological pathway enriched in Alzheimer’s disease (from the current study); (7) expression correlated with the BRAAK stage; and (8) differential expression in a 1 + Alzheimer’s disease (A

*CASP7*

Locus – Data to collect

Gene: CASP7

Evaluator: Farrer

Date: 1/7/20

1. Significance
a. discovery study

i. study size (#cases/controls) -- 507 enriched cases, 4,917 controls (EA)

172 enriched cases, 179 controls (CH)^a^

ii. evaluation of population heterogeneity (yes/no/comments) – yes
iii. multiple test/model corrections (yes/no/comments) – yes (rs116437863, p = 2.44x10^-10^ which is study-wide significant, i.e., threshold p = 4.98x10^-7^ in EAs )
     b. meta-analysis – discovery study -- N/A
iv. study size (#cases/controls)
v. evaluation of population heterogeneity (yes/no/comments)
vi. multiple test/model corrections (yes/no/comments)
    c. replication studies – replication attempted in 2 EA GWAS datasets with multiplex families:

MIRAGE (704 cases, 449 controls) and NIA-LOAD (1,457 cases, 1,568 controls)^a^

- independent investigators and data -
- larger study with added samples
  d. replication studies – evidence -- p = 0.061 in total replication sample; significance increased in discovery + replication samples (p = 1.92x10^-10^)^a^
- signal direction – same in discovery and both replication datasets
- other ethnic groups study size (#cases/controls)
- evaluation of population heterogeneity (yes/no/comments)
- multiple test/model corrections (yes/no/comments)
  e. other genetic studies –

i. A genome-wide haplotype association study identified *CASP7* as associated with AD in Caribbean Hispanic individuals (global haplotype p = 9.5x10^-4^; haplotype specific p = 5.33x10^-5^).^b^ Data were obtained from a previous association study of CNVs and AD.^c^
ii. Ayers et al. analyzed genetic and phenotypic data from 6 datasets (including the ADSP discovery WES dataset) consisting of 12,248 AD cases and 20,067 controls identified a loss of function variant in *CASP7* (rs10553596) that protects against AD in *APOE* ε4 homozygotes (OR = 0.45, p = 0.004).^d^ This variant is different from the one identified in the ADSP enriched cases study. Analysis of RNA sequencing derived gene expression data indicated the variant correlates with reduced caspase 7 expression in multiple brain tissues.^d^
2. Pleiotropy
   a. AD-related phenotype (*e.g*. CSF biomarker association)
i. *CASP7* is a protease involved in apoptosis and inflammation.^e^ Activation of caspase apoptotic pathways involves apoptosome assembly^f^ and multiple recent studies link this process to aging and AD neuropathology, including caspase cleavage of amyloid precursor protein^g,h,i^ and tau.^h,j^ Alternate processing of APP may result in cleavage of the C31 fragment by the protease encoded by CASP7.^h^ C31 is one of several C-terminal fragments produced from APP and there is some evidence that it is toxic.^k^ Roles in AD for multiple caspases are well described and supported. Two rare variants in *CASP8* associated with AD by targeted sequencing of genes involved in amyloid metabolism.^l^
ii. In another study, *CASP7* was among a small group of genes in a cluster identified by pathway analysis that was seeded with GWAS results for multiple MRI and cognitive endophenotypes in the ADNI dataset.^m^
    b. Non-AD phenotype (*e.g*. coronary artery disease, *APOE*)
       - Recommended tier – gene tier 1, locus tier 1
       - Supporting references –
1. Zhang et al. A rare missense variant of *CASP7* is associated with familial late-onset Alzheimer's disease. Alzheimer Dement 2019; 15(3):441-452
2. Shang Z, Lv H, Zhang M, Duan L, Wang S, Li J, Liu G, Ruijie Z, Jiang Y. Genome-wide haplotype association study identify TNFRSF1A, CASP7, LRP1B, CDH1 and TG genes associated with Alzheimer's disease in Caribbean Hispanic individuals. Oncotarget. 2015; 6:42504-42514.
3. Ghani M, Pinto D, Lee JH, Grinberg Y, Sato C, Moreno D, Scherer SW, Mayeux R, St George-Hyslop P, Rogaeva E. Genome-wide survey of large rare copy number variants in Alzheimer's disease among Caribbean hispanics. G3 (Bethesda). 2012; 2:71-78
4. Ayers KL, Mirshahi UL, Wardeh AH, Murray MF, Hao K, Glicksberg BS, Li S, Carey DJ, Chen R. A loss of function variant in CASP7 protects against Alzheimer's disease in homozygous APOE ε4 allele carriers. BMC Genomics. 2016; 17 Suppl 2:445.
5. Lamkanfi M, Kanneganti TD. Caspase-7: a protease involved in apoptosis and inflammation. Int J Biochem Cell Biol. 2010; 42:21-24.
6. Cullen SP, Martin SJ. Caspase activation pathways: some recent progress. Cell Death Differ. 2009; 16:935-938.
7. Zhao M, Su J, Head E, Cotman CW. Accumulation of caspase cleaved amyloid precursor protein represents an early neurodegenerative event in aging and in Alzheimer's disease. Neurobiol Dis. 2003; 14:391-403.
8. Rohn TT, Kokoulina P, Eaton CR, Poon WW. Caspase activation in transgenic mice with Alzheimer-like pathology: results from a pilot study utilizing the caspase inhibitor, Q-VD-OPh. Int J Clin Exp Med. 2009; 2:300-308.
9. Fiorelli T, Kirouac L, Padmanabhan J. Altered processing of amyloid precursor protein in cells undergoing apoptosis. PLoS One. 2013; 8:e57979.
10. Cotman CW, Poon WW, Rissman RA, Blurton-Jones M. The role of caspase cleavage of tau in Alzheimer disease neuropathology. J Neuropathol Exp Neurol. 2005; 64:104-112.
11. Nhan HS, Chiang K, Koo EH. The multifaceted nature of amyloid precursor protein and its proteolytic fragments: friends and foes. Acta Neuropathol. 2015; 129:1-19
12. Rehker J, Rodhe J, Nesbitt RR, Boyle EA, Martin BK, Lord J, et al. Caspase-8, association with Alzheimer's Disease and functional analysis of rare variants. PLoS One. 2017; 12:e0185777
13. Sloan CD, Shen L, West JD, Wishart HA, Flashman LA, Rabin LA, Santulli RB, Guerin SJ, Rhodes CH, Tsongalis GJ, McAllister TW, Ahles TA, Lee SL, Moore JH, Saykin AJ. Genetic pathway-based hierarchical clustering analysis of older adults with cognitive complaints and amnestic mild cognitive impairment using clinical and neuroimaging phenotypes. Am J Med Genet B Neuropsychiatr Genet. 2010; 153B:1060-1069.

*CD33*

**Gene: CD33**

**Evaluator: Mayeux**

**Date 11/14/19**

**A. Primary evidence**

Locus

MAF	p	rs3826656	0.36	FBAT/GEE 0.4E-5 (first paper)

intronic variant

rs3865444	0.21	SNV

Bertram L, Lange C, Mullin K, Parkinson M, Hsiao M, Hogan MF, Schjeide BM, Hooli B, Divito J, Ionita I, Jiang H, Laird N, Moscarillo T, Ohlsen KL, Elliott K, Wang X, Hu-Lince D, Ryder M, Murphy A, Wagner SL, Blacker D, Becker KD, Tanzi RE. Genome-wide association analysis reveals putative Alzheimer's disease susceptibility loci in addition to APOE. Am J Hum Genet. 2008; 83:623-32. PubMed PMID: 18976728; PubMed Central PMCID: PMC2668052.

Jiang YT, Li HY, Cao XP, Tan L. Meta-analysis of the association between CD33 and Alzheimer's disease. Ann Transl Med. 2018 May;6(10):169. doi: 10.21037/atm.2018.04.21. PubMed PMID: 29951491; PubMed Central PMCID: PMC5994519. (p<0.01).

Naj AC, et al. Common variants at MS4A4/MS4A6E, CD2AP, CD33 and EPHA1 are associated with late-onset Alzheimer's disease. Nat Genet. 2011;43:436-41. PubMed PMID: 21460841; PubMed Central PMCID: PMC3090745. (GERAD met p = 1.6E-9).

Hollingworth P, et al. Common variants at ABCA7, MS4A6A/MS4A4E, EPHA1, CD33 and CD2AP are associated with Alzheimer's disease. Nat Genet. 2011;43:429-35. PubMed PMID: 21460840; PubMed Central PMCID: PMC3084173. (ADGC p = 2.0E-7).

Jansen IE, et al. Genome-wide meta-analysis identifies new loci and functional pathways influencing Alzheimer's disease risk. Nat Genet. 2019; 51:404-413. PubMed PMID: 30617256; PubMed Central PMCID: PMC6836675. (rs3865444, p= 6.3E-9)

1. Altered function SNV (stop gain, stop loss, splicing)

Intronic variants and SNV associated with late onset AD

Splice variant affects expression of CD33 which may affect amyloid- clearance.

Siddiqui SS, et al. The Alzheimer's disease-protective CD33 splice variant mediates adaptive loss of function via diversion to an intracellular pool. J Biol Chem. 2017 15; 292:15312-15320 PubMed PMID: 28747436; PubMed Central PMCID: PMC5602391.

2. Structural variants – disrupt expression

HLOF are known to affect expression but not associated with AD. rs3865444 is associated with greater expression of the full-length CD33 isoform, but rs3826656 needs further characterization.

Malik M, et al. CD33 Alzheimer's risk-altering polymorphism, CD33 expression, and exon 2 splicing. J Neurosci. 2013; 33:13320-5. PubMed PMID: 23946390; PubMed Central PMCID: PMC3742922.

The association of TREM2 expression with AD may be mediated by accumulation of amyloid pathology, as is the case for CD33 suggesting an interaction between TREM2 and CD33.

3. Significant association evidence with AD

See above. CD33 locus is only significant in meta-analyses listed above

4. Replication in another population

A meta-analyses of several studies demonstrated that different variants in *CD33* were nominally associated with AD (rs3865444: OR =0.94; 95% CI, 0.90-0.98, P<0.01; rs3826656: OR =0.94; 95% CI, 0.62-1.41, P<0.01). We made subgroup analysis which was stratified by race. There were protective associations in Caucasians but not in Asians among *CD33* rs3865444 polymorphism (Caucasians: OR =0.92; 95% CI, 0.90-0.94, P=0.05; Asians: OR =0.87; 95% CI, 0.65-1.17, P<0.01).

5. Gene expressed in an AD-relevant tissue/cell type

CD33 is expressed primarily in microglia.

**B. Supporting evidence**

Three common variants significantly associated with AD. Main papers are listed above.

**C. Pleiotropy**

Variant rs3865444 in CD33 has been associated with the development, maturation, or regulation of WBC types (meta-analysis p=6.8E-14). Tajuddin SM et al. Large-Scale Exome-wide Association Analysis Identifies Loci for White Blood Cell Traits and Pleiotropy with Immune-Mediated Diseases. Am J Hum Genet. 2016;99:22-39PubMed PMID:27346689; PubMed Central PMCID: PMC5005433.

D. Common variants in the same gene in another ethnic group: African Americans, Caribbean Hispanic, Korean, Japanese.

**Asian (Chinese and Korean) All weak associations**

Yuan Q, Chu C, Jia J. Association studies of 19 candidate SNPs with sporadic Alzheimer's disease in the North Chinese Han population. Neurol Sci. 2012;33(5):1021-8. PubMed PMID: 22167654.

Deng YL, et al. The prevalence of CD33 and MS4A6A variant in Chinese Han population with Alzheimer's disease. Hum Genet. 2012 131:1245-9. PubMed PMID: 22382309.

Chung SJ, et al. Association of GWAS top hits with late-onset Alzheimer disease in Korean population. Alzheimer Dis Assoc Disord. 2013;27:250-7. PubMed PMID: 22975751.

Li X, et al.. CD33 rs3865444 Polymorphism Contributes to Alzheimer's Disease Susceptibility in Chinese, European, and North American Populations. Mol Neurobiol. 2015; 52(1):414-21. PubMed PMID: 25186233.

**African Americans (no association with rs3865444)**

Hohman TJ, et al. Global and local ancestry in African-Americans: Implications for Alzheimer's disease risk. Alzheimers Dement. 2016;12:233-43. PubMed PMID: 26092349; PubMed Central PMCID: PMC4681680.

Logue MW, et al. A comprehensive genetic association study of Alzheimer disease in African Americans. Arch Neurol. 2011;68:1569-79. PubMed PMID: 22159054; PubMed Central PMCID: PMC3356921. (novel variant rs10419982; p = 5.4E-4).

**Colombian Hispanic (with rs3865444)**

Moreno DJ, et al. Association of GWAS Top Genes With Late-Onset Alzheimer's Disease in Colombian Population. Am J Alzheimers Dis Other Demen. 2017;32:27-35. PubMed PMID: 28084078. (p,<0.05).

**E. Co-segregation in families**

None

**F. Other**

Chan G, et al. CD33 modulates TREM2: convergence of Alzheimer loci. Nat Neurosci. 2015; 18:1556-8. PubMed PMID:26414614; PubMed Central PMCID: PMC4682915.

“Consistent with this latter study, our results reporting an association of the CD33 AD risk allele with increased TREM2, as well as higher cortical *TREM2* RNA expression with increasing amyloid pathology, support a pathogenic role for increased TREM2 expression by peripherally-derived myeloid cells in AD susceptibility.”

Chauhan G, et al. Association of Alzheimer's disease GWAS loci with MRI markers of brain aging. Neurobiol Aging. 2015;36:1765.e7-1765.e16. PubMed PMID: 25670335; PubMed Central PMCID: PMC4391343.

Rs3865444 was associated with intracranial volume (b -5.2, p = 5.8E-3)

Walker DG, et al. Association of CD33 polymorphism rs3865444 with Alzheimer's disease pathology and CD33 expression in human cerebral cortex. Neurobiol Aging. 2015; 36:571-82. PubMed PMID: 25448602; PubMed Central PMCID: PMC4315751.

“…reduced levels of CD33 protein in brain tissue of cases by genotype (rs3865444) and the demonstration of downregulation of CD33 in human microglia treated with Aβ or certain inflammatory agents. It had been hypothesized that as the CD33 rs3865444 SNP was located just 373 bp upstream of exon 1 of CD33, it might affect gene transcription or RNA stability.”

Wang WY, et al. Impacts of CD33 genetic variants on the atrophy rates of hippocampus and parahippocampal gyrus in normal aging and mild cognitive impairment. Mol Neurobiol 2017; 54: 1111-8; PMID 26803496

“SNPs were significantly associated with advanced atrophy of left hippocampal gyrus (rs73932888 and rs8112072) and CA1 region (rs1803254).” There was no association between expression and rs3865444

**Recommendation:** Tier 3 locus and Tier 3 Gene.

SCARA3 (CLU locus)

Locus – Data to collect

Gene: SCARA3 (CLU locus)

Evaluator: Wang

Date: October 3, 2019

Significance

A. discovery study: **1 Baker 2019**

1. study size (#cases/controls)

a. Stage 1: GERAD
b. 3,332 cases / 9832 controls
c. GWAS chip with HRC r1.1 2016 imputation
d. POLARIS Gene-based analysis (Table 1) with 240 SNPs
   i. P = 7.8x10^-7^
   ii. Beta = 0.526
e. POLARIS Gene-based analysis conditioned on APOE (Table 1)
   i. P= 8.3x10^-7^
   ii. Beta = 0.537

2. evaluation of population heterogeneity (yes/no/comments)

a. No

b. multiple test/model corrections (yes/no/comments)

c. Yes (significant after correcting for number of genes tested)

3. meta-analysis – discovery study NONE

- study size (#cases/controls)
- evaluation of population heterogeneity (yes/no/comments)
- multiple test/model corrections (yes/no/comments)

4. replication studies Not replicated in 2 Marioni 2018

a. UK Biobank 314,278 participants

b. 27,696 maternal cases, 14,338 paternal cases, proxy for AD genetic study

c. Meta-analysis with IGAP summary statistics (N=74,046)

d. Gene-based analysis with 155 SNPs (Table S5)

e. P=2.2x10^-2^

f. Non replication may be due to older imputation panel / fewer SNPs

5. replication studies – evidence NONE

a. signal direction

b. other ethnic groups study size (#cases/controls)

c. evaluation of population heterogeneity (yes/no/comments)

d. multiple test/model corrections (yes/no/comments)

6. Pleiotropy

a. AD-related phenotype (*e.g*. CSF biomarker association) NONE

b. Non-AD phenotype (*e.g*. coronary artery disease, *APOE*) Cancer, by gene expression not genetics.

- Clinical samples showed an inverse correlation between SCARA3 gene expression, myeloma progression, and favorable clinical prognosis (3 Brown 2013).
- *SCARA3* mRNA is overexpressed in ovarian carcinoma compared with breast carcinoma effusions (4 Bock 2012)

c. Recommended tier. Tier 2 (P<2.5x10^-6^ but no replication)

- Supporting references

1. *Baker, E. et al. Gene-based analysis in HRC imputed genome wide association data identifies three novel genes for Alzheimer's disease. PLoS One* ***14****, e0218111 (2019).*
2. *Marioni, R.E. et al. GWAS on family history of Alzheimer’s disease. Transl Psychiatry 2018 8(1):99.*
3. *Brown et al. Scavenger receptor class A member 3 (SCARA3) in disease progression and therapy resistance in multiple myeloma. Leukemia Research 37(8):963-969 (2013).*
4. *Bock et al.* *SCARA3 mRNA is overexpressed in ovarian carcinoma compared with breast carcinoma effusions. Human Pathology 43(5):669-674 (2012)*

Other articles:

1. Sun et al. Molecular differences in Alzheimer's disease between male and female patients determined by integrative network analysis. J Cellular and Molecular Medicine doi.org/10.1111/jcmm.13852 (2018)
   a. SCARA3 is differentially expressed control vs AD in dorsolateral prefrontal cortex
2. Humphries et al. Alzheimer disease (AD) specific transcription, DNA methylation and splicing in twenty AD associated loci. Molecular and Cellular Neuroscience 67:37-45 (2015)
   a. SCARA3 has higher expression in LOAD and Dementia with Lewy Bodies vs normal control.
3. Xu et al. A SAGE study of apolipoprotein E3/3, E3/4 and E4/4 allele-specific gene expression in hippocampus in Alzheimer disease. Molecular and Cellular Neuroscience 36(3):313-331 (2007).

*CR1*

Locus – Data to collect

Gene: CR1

Evaluator: Wang

Date: November 27, 2019

1. Significance

A. discovery study.

Earliest discovery were from the two Nature Genetics papers in 2009

- Lambert Nature Genetics 2013
- Top SNP in CR1 locus: rs6656401 (chr1:207692049, GRCh37)
- Stage 1: 17,008 cases, 37,154 controls of European ancestry
  - Imputed to 1000 genome panel (7,055,881 SNPs)
  - Top SNP: MAF=0.197, OR=1.17, P=7.7x10^-15^
- Stage 2: 8,572 cases, 11,312 controls of European ancestry
  - 11,632 SNPs IGAP chip
  - OR=1.21, P=7.9x10^-11^
- Meta-analysis Stage 1+2:
  - OR=1.18, P=5.7x10^-24^

B. meta-analysis – discovery study

C. replication studies.

- Kunkle Nature Genetics 2019
  - Top SNP in CR1 locus: rs4844610 (chr1:207802552, GRCh37)
  - Stage 1: 21,982 cases, 41,944 controls, 63,926 total of European ancestry
    - Imputed to newer 1000 genome panel (9,456,058 SNPs)
    - Top SNP: MAF=0.187, OR=1.16, P=8.2x10^-16^
  - Stage 2: 18,845 total of European ancestry
    - 11,632 SNPs IGAP chip
    - OR=1.20, P=3.8x10^-10^
  - Mehta-analysis Stage 1+2:
    - OR=1.17, P=3.6x10^-24^
- IGAP Exome Chip study Nature Genetics 2017
  - h exome chip study 2017 Nature Genetics
  - Rs6656401 MAF=6.7%, P=7e-11, OR-=1.17

D. replication studies – evidence

- African Americans:
  - Reitz JAMA 2013
    - 1,968 cases and 3,928 controls, African American GWAS
    - rs146366639 OR=0.82, *P* = .0005
- Asian population:
  - Jin Neuroscience Letter 2012
    - 1019 cases, 1080 controls Han Chinese
    - Rs66546401
    - OR=1.69, P=0.005
  - Ma Neurobiology of Aging 2014
    - Targeted sequencing of 100 to find 22 variants in CR1, followed by targeted SNP genotyping. Total 2292 individuals
    - Adjusting for sex, age at onset, and APOE ε4 status
    - rs116806486: dominant model: *p* = 0.006, OR = 3.381
    - rs6691117: additive model: *p* = 0.021, OR =1.204, similar significance in dominant and recessive models
  - Miyashita 2013 PLoS ONE
- Japanese (1008 cases and 1016 controls) and Korean (339 cases and 1129 controls)
- Rs6656401:
- Japanese MAF=0.04, P=9e-3, OR=1.38
- Korean MAF=0.04, P=0.0375, OR=1.24
- Rs3818361:
- Japanese MAF=0.39, P=0.254, OR=0.94
- Korean MAF=0.31, P=0.408, OR=0.92
- Family history
  - GWAS on family history of Alzheimer's disease, Translational Psychiatry 2018
    - up to 42,034 British ancestry individuals with parental history of Alzheimer's disease, at least 272,244 British ancestry individuals with no parental history of Alzheimer's disease, 25,580 Alzheimer's disease cases, 48,466 controls
  - Genome-wide meta-analysis identifies new loci and functional pathways influencing Alzheimer's disease risk. Jansen Nature genetics 2019
    - Proxy GWAS
- Rare/Coding/intronic variants
  - Rs3818361 CR1 intronic
  - Li Y Mol Neurobiol 2016
    - Chinese: 1244 cases 2803 controls, N=4047
    - European: use summary stats from Harold Nat Genet 2009 (N=3941 cases/7848 controls)
    - rs3818361
    - Chinese: OR=1.16 P = 6.00E-03
    - European: OR=1.15 P = 5.00E-03
  - Keenan HMG 2012
    - Rs4844609 p.Ser1610Thr
    - ROS/MAP N=1709
    - rs4844609 (*P* = 0.003)
  - Van Cauwenberghe Neurobiol aging 2013
    - Rs4844609 p.Ser1610Thr
    - Flanders-Belgian study cohort: 1276 case, 1128 control
    - Not associated with AD, memory impairment, total tau, Ab42, pTau
    - Does not explain rs3818361/rs6656401 or CR1CNV
    - Question the role of this variant
- Structural variants
  - Brouwers et al. Molecular Psychiatry 2012
    - 1883 subjects from a Flanders-Belgian cohort
    - Copy number variation: haplotypes carry different overlapping copy numbers of 18-kb long low-copy repeats (LCRs) and long homologous repeats (LHRs)
    - Replicated in French cohort n=2003
    - Combined analysis OR=1.32, P=0.0025

2. Pleiotropy

- AD-related phenotype (e.g., CSF biomarker association)
  - Brouwers 2012 Molecular Psychiatry
    - CR1 haplotype interacts with APOE (CSF biomarker association, P_adj<0.006, OR=1.5)
    - Four SNPs are associated with Increased Ab1-42 level in CSF
- Non-AD phenotype (e.g., coronary artery disease, *APOE*)
- Naitza PLoS Genet 2012
  - 4,694 Sardinian individuals
  - Inflammatory biomarkers: erythrocyte sedimentation rate (ESR)
  - Rs12034598 P=9e-14, RAF=0.408
- Li Pharmacogenet Genomics 2016
  - 1390 Europeans
  - paliperidone efficacy
  - Rs56161922: P=7e-6, 0.38 unit decrease
- Zhu 2019 Hum Genet
  - 54,162 European ancestry cases, 51,750 European ancestry individuals
  - Rs6656401
  - Fasting insulin: beta=9.6e-3, P=0.018
  - Protein QTL: Polymeric immunoglobulin receptor, Protein S100-A5, Complement decay-accelerating factor 3.71e-15

3. Recommended tier:

- Locus: Tier 1. highly confident: p< 2.5x-10-6 and replication
- Gene: Tier 2. Suggestive evidence of an association (CNV evidence)

*HLA-DRB5*

Locus – Data to collect

Gene: *HLA-DRB1/HLA-DRB5*

Evaluator: Kunkle

Date: 1/16/2020

A. Significance

1. discovery study (Lambert et al. 2013)^1^

study size (#cases/controls)

Stage 1: 17,008 cases, 37,154 controls

GWAS array

Lead SNP: rs9271192

OR (95% CI): 1.11 (1.07-1.16)

P-value: 1.6 x 10^-8^

MAF: 0.276

a. ancestry	non-Hispanic white

b. evaluation of population heterogeneity (yes/no/comments)

Yes

c. multiple test/model corrections (yes/no/comments)

logistic regression models adjusting for age, sex, principal components and APOE

d. meta-analysis – discovery study

study size (#cases/controls)

Stage 2: 8,572 cases and 11,312 controls

GWAS array

OR (95% CI): 1.12 (1.06 - 1.18)

P-value: 4.2 x 10^-5^

Meta-analysis Stage 1+2: 25,580 cases and 48,466 controls

GWAS array

OR (95% CI): 1.11 (1.08 - 1.15)

P-value: 2.9 x 10^-12^

e. evaluation of population heterogeneity (yes/no/comments)

Yes

f. multiple test/model corrections (yes/no/comments)

Inverse-variance weighted fixed effects meta-analysis

2. larger study with added samples^2,3,4^

a. Kunkle et al. 2019

Study size (#cases/controls): 30,344 cases and 52,427 controls (ADGC, EADI, CHARGE, GERAD)

Ancestry: non-Hispanic white

Lead SNP: rs9271058

MAF: 0.270

OR (95% CI): 1.10 (1.07 – 1.13)

P-value: 1.4 x 10^-11^

evaluation of population heterogeneity (yes/no/comments): yes

multiple test/model corrections (yes/no/comments): yes

b. Jansen et al. 2019

Study size (#cases/controls): 71,880 cases and 383,378 controls (ADSP, IGAP, PGC-ALZ, UKBB)

Ancestry: non-Hispanic white

Lead SNP: rs6931277

MAF: 0.148

OR (CI): not available

P-value: 8.41 x 10^-11^

evaluation of population heterogeneity (yes/no/comments): yes

multiple test/model corrections (yes/no/comments): yes

Recommended tier (locus): tier 1

Recommended tier (gene): tier 2

*IGHV1-67*

***IGHV1-67: chr 14* Tier 4: Insufficient evidence to determine whether an association exists**

A) p = 7.9×10^−8^

<https://www.ncbi.nlm.nih.gov/pubmed/24922517>

International Genomics of Alzheimer's Project Consortium, comprising over 7 m genotypes from 25,580 Alzheimer's cases and 48,466 controls.

Based on IGAP p-values; not individuals level data; Fisher test

SNP filters: autosomal SNPs, (MAF) ≥0.01 and imputation quality >=0.3 in each individual study

B) Not listed in <https://www.nature.com/articles/s41398-018-0150-6>
C) Not replicated by <https://www.sciencedirect.com/science/article/pii/S2352396418304122>
D) Not listed in GWAS catalog

Locus – Data to collect

Locus: IGHV1-67

Evaluator: DeStefano

Date:	12/4/2019

E) Significance

1. discovery study^1^

a. Gene-based approach: Fisher’s statistic applied to summary data (note: Kwok et al raise b. issues around type I error for this test)

b. study size (#cases/controls):

- Stage 1 IGAP GWAS
- Imputed with Impute2 or MACH using 1000G
- 17,008 cases/37,154 controls
- rs2011167 (OR = 1.082, p-value = 2.23E-3), rs1961901 (OR = 0.924, p-value = 2.23E-3). Gene-wide p-value: 2.3 x 10^-4^
- MAF cases not mentioned, MAF controls not mentioned

c. evaluation of population heterogeneity (yes/no/comments)

- not mentioned

d. multiple test/model corrections (yes/no/comments)

- Loci with p-value (combined over all SNPs at the locus) < 10^-4^ were selected for replication

e. Stage 2 (Replication)

- 8,572 cases/11,312 controls of European ancestry
- Genotyped
- MAF cases not mentioned /MAF controls not mentioned;
- rs2011167 (OR = 1.238, p-value = 0.0013), rs1961901 (OR = 0.8135, p-value = 0.0018). Gene-wide p-value: 3.2 x 10^-5^
- Effect direction/approximate effect size same as discovery
- evaluation of population heterogeneity (yes/no/comments); not mentioned
- multiple test/model corrections (yes/no/comments); yes multiple testing

f. Combined Discovery + Replication ^1^

- Combined Gene-wide P-value from Fisher’s method = 7.9 x 10^-8^

g. Replication attempts (on same data as Discovery Study above)

h. gene-based approach: an overall P-value for the association of each autosomal gene with late-onset AD was obtained by combining P-values for the association of all SNPs within each gene using GATES

i. study size (#cases/controls):

- Stage 1 IGAP GWAS
- Imputed with Impute2 or MACH using 1000G
- 17,008 cases/37,154 controls
- Did not meet genome-wide threshold P-value < .000002

j. AD-related phenotype (*e.g*. CSF biomarker association)

k. Nothing listed in GWAS catalog. Function of this gene is unknown

l. Non-AD phenotype (*e.g*. coronary artery disease, *APOE*)

MS4A

Gene(s): MS4A6A, MS4A4A

Evaluator: Alison Goate and Edoardo Marcora Date: January 16, 2020

### 1. Significance

- Hollingworth et al. (2011) Nat Genet. 43:429-35 [DOI:10.1038/ng.803](https://doi.org/10.1038/ng.803)
  - GWAS chips (Illumina and Affy) + HapMap/1000G CEU imputation
  - SNP-based analysis
    - rs610932-T (11:59939307:T:G:hg19, gnomAD-NFE AF=0.60, 3’ UTR of MS4A6A)
    - rs670139-T (11-59971795-G-T:hg19, gnomAD-NFE AF=0.43, intergenic between MS4A6A and MS4A4E)
  - Stage 1 (discovery, 496,763 SNPs, genome-wide sig thresh P=5.0x10^-8^)
    - GERAD1, ADNI, TGEN1 and EADI1
    - 6,688 cases and 13,685 controls
    - rs610932-T
      - P = 1.8x10^-8^ (the only SNP that passed sig thresh!)

## OR = 0.88 (0.85-0.92)

- rs670139-T
  - P = 1.0x10^-5^

## OR = 1.11 (1.06-1.16)

- Stage 2 (replication, 12 SNPs selected from 61 with P≤1x10^−5^, Bonferroni-adjusted sig thresh P=4.2x10^−3^)
  - GERAD2, deCODE, AD-IG
  - 4,896 cases and 4,903 controls
  - rs610932-T
    - P=1.6x10^-3^

## OR=0.90 (0.84-0.96)

- rs670139-T
  - P = 1.1x10^-3^

## OR = 1.11 (1.04-1.19)

- Stage 3 (replication, 3 SNPs selected from 5 with P≤0.05, Bonferroni-adjusted sig thresh P=1.7×10^−2^)
  - EADI2, CHARGE, Mayo2
  - 8,286 cases and 21,258 controls
  - rs610932-T
    - P=2.1x10^-5^

## OR=0.91 (0.87-0.95)

- rs670139-T
  - P = 3.2x10^-3^

## OR = 1.06 (1.02-1.11)

- Meta-analysis of stages 1, 2 and 3
  - 19,870 cases and 39,846 controls
  - rs610932-T
    - P=1.8x10^-14^

## OR=0.90 (0.87-0.92)

- rs670139-T
  - P = 1.4x10^-9^

## OR = 1.09 (1.06-1.12)

- Meta-analysis of this study with Naj et al. (2011) after removing overlapping samples (GERAD+ and ADGC)
  - In main table/text but not described in Methods
  - rs610932-T
    - P=1.2x10^-16^

## OR=0.91 (0.88-0.93)

- rs670139-T
  - P = 1.1x10^-10^

## OR = 1.08 (1.06-1.11)

- Region of association spans ~300Kb and includes 6/16 MS4A cluster genes:
  - MS4A2, MS4A3, MS4A4A, [MS4A4E], MS4A6A and MS4A6E
  -
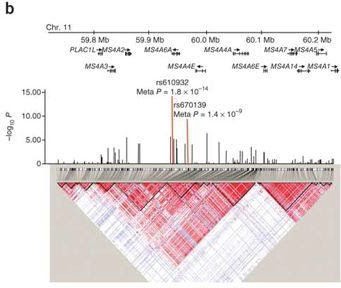

  - rs610932-T has a weak eQTL signal in some brain regions, but not in LCLs or other brain regions
  - The non-synonymous SNP most strongly associated with rs610932-T is rs2304933 (P=0.006 in Stage 1)
- Naj et al. (2011) Nat Genet. 43:436-41 [DOI:10.1038/ng.801](https://doi.org/10.1038/ng.801)
  - GWAS chips (Illumina and Affy) + HapMap CEU imputation
  - SNP-based analysis
    - rs4938933-C (11:60034429:C:T:hg19, gnomAD-NFE AF=0.62, intergenic between MS4A4E and MS4A4A)
  - Stage 1 (discovery, 2,324,889 SNPs, genome-wide sig thresh P=5.0x10^-8^)
    - ACT, ADC1, ADC2, ADNI, GenADA, UM/VU/MSSM, MIRAGE, NIA-LOAD, OHSU, TGEN2
    - 8,309 cases and 7,366 controls
    - rs4938933-C
      - Pjoint = 4.5x10^-8^
      - ORjoint = 0.87 (0.83-0.92)
  - Stage 2 (replication, ??? SNPs at 9 loci excluding APOE with P≤1x10^−6^ + ABCA7 SNPs, Bonferroni-adjusted sig thresh P=???)

#### ADC3, MAYO, ROSMAP, UP, WU

- 3,531 cases and 3,565 controls
- rs4938933-C
  - Pjoint = 4.4x10^-3^
  - ORjoint = 0.90 (0.84-0.97)
- Meta-analysis of stage 1 and 2
  - 11,840 cases and 10,931 controls
  - rs4938933-C
    - Pmeta = 1.7x10^-9^ (the only SNP that passed sig thresh!)
    - ORmeta = 0.88 (0.85-0.92)
  - rs4939338-T (11:60099225:T:C:hg19, gnomAD-NFE AF=0.65, intergenic

between MS4A4A and MS4A6E)

- Pmeta = 2.6x10^-11^
- ORmeta = 0.87 (0.84-0.91)
- Meta-analysis of this study (stage 1+2) with Hollingworth et al. (2011) after removing overlapping samples (ADGC and CHARGE/GERAD/EADI1)
  - 18,762 cases and 29,827 controls
  - rs4938933-C
    - Pmeta = 8.2x10^-12^
    - ORmeta = 0.89 (0.87-0.92)

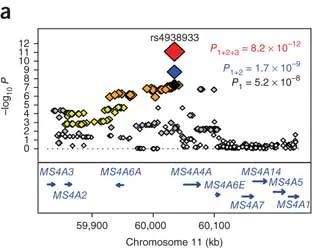

- Lambert et al. (2013) Nat Genet. 45:1452-8 [DOI:10.1038/ng.2802](https://doi.org/10.1038/ng.2802)
  - GWAS chips (Illumina and Affy) + 1000G EUR imputation
  - SNP-based analysis
    - rs983392-G (11:59923508:A:G:hg19, gnomAD-NFE AF=0.40, intergenic between MS4A4E and MS4A4A)
  - Stage 1 (discovery, 7,055,881 SNPs, genome-wide sig thresh P=5.0x10^-8^)

#### ADGC, CHARGE, EADI, GERAD

- 17,008 cases and 37,154 controls
- rs983392-G
  - P = 2.8x10^-11^

## OR = 0.90 (0.87-0.93)

- Stage 2 (replication, 11,632 SNPs at 14 loci excluding APOE with P≤1x10^−3^, Bonferroni-adjusted sig thresh P=4.3x10^-6^)
  - Other cohorts from Austria, Belgium, Finland, Germany, Greece, Hungary, Italy, Spain, Sweden, the UK and the United States
  - 8,572 cases and 11,312 controls
  - 116 SNPs below sig thresh in stage 2, 80 of these below genome-wide sig thresh in stage 1 (only CD33 and DSG2 below genome-wide sig thresh in stage 1 did not replicate in stage 2)
  - rs983392-G
    - P = 4.5x10^-6^

## OR = 0.90 (0.86-0.94)

- Meta-analysis of stage 1 and 2
  - 25,580 cases and 48,466 controls
  - rs983392-G
    - P = 6.1x10^-16^

## OR = 0.90 (0.87-0.92)

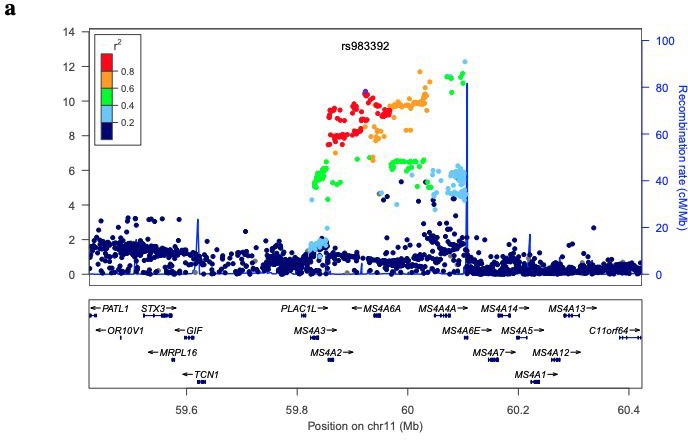

- Marioni et al. (2018) Transl Psychiatry. 8:99 [DOI:10.1038/s41398-018-0150-6](https://doi.org/10.1038/s41398-018-0150-6)
  - GWAS chips (Axiom) + HRC imputation
  - SNP-based analysis
    - rs1582763-A (11:60021948:G:A:hg19, gnomAD-NFE AF=0.36,

intergenic between MS4A4E and MS4A4A)

- Meta-analysis of maternal and paternal AD/dementia GWAX (7,795,605 SNPs, genome-wide sig thresh P=5.0x10^-8^)

#### UKBB

- 27,696 cases of maternal AD/dementia (260,980 controls)
- 14,338 cases of paternal AD/dementia (245,941 controls)
- rs1582763-A
  - P = 1.0x10^-5^
- Meta-analysis of this study (UKBB AD GWAX) and Lambert et al. (2013) stage 1 merged with stage 1+2
  - rs1582763-A
    - P = 1.0x10^-18^ [PIGAP = 1.8x10^-15^]
  - rs983392-G
    - P = 4.9x10^-18^ [PIGAP = 6.1x10^-16^]

#### OR = 0.92 [ORIGAP = 0.90]

- Gene-based analysis of UKBB AD GWAX + IGAP AD GWAS
  - A total of 87 genes were significant at a Bonferroni threshold of 2.7x10^-6^
  - MAGMA SNP-wise (mean) model with default settings and 1000G phase3 reference LD panel
  -
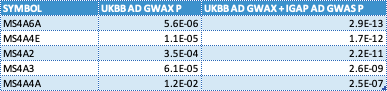

- Locus-to-gene mapping: Performed SMR analysis using Common Mind Consortium brain tissue eQTLs but not much signal overall (and no signal at MS4A locus)
- Jansen et al. (2019) Nat Genet. 51:404-13 [DOI:10.1038/s41588-018-0311-9](https://doi.org/10.1038/s41588-018-0311-9)
  - SNP-based analysis
    - rs2081545-A (11:59958380:C:A:hg19, gnomAD-NFE AF=0.38,

intergenic between MS4A6A and MS4A4E)

- AD and AD-by-proxy (71,880 cases and 383,378 controls)
-
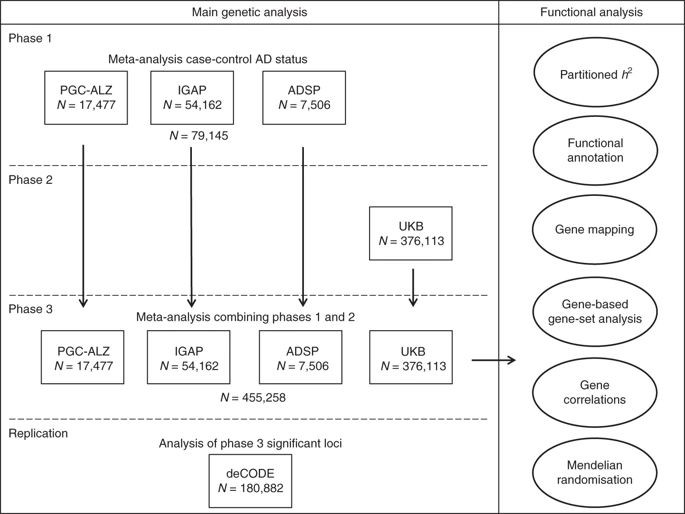

- 9,862,738 SNPs in Phase 1, 1000G phase3 imputation for GWAS and WES, HRC imputation for GWAX
-
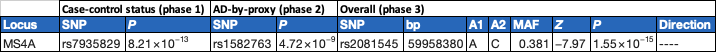

- Gene-based analysis (using MAGMA)
  - A total of 97 genes were significant at a Bonferroni threshold of 2.7x10^-6^
  - MAGMA SNP-wise (mean) model with default settings and 1000G phase3 reference LD panel
  -
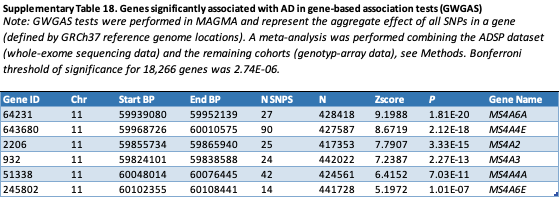

- Locus-to-gene mapping (using FUMA)
  -
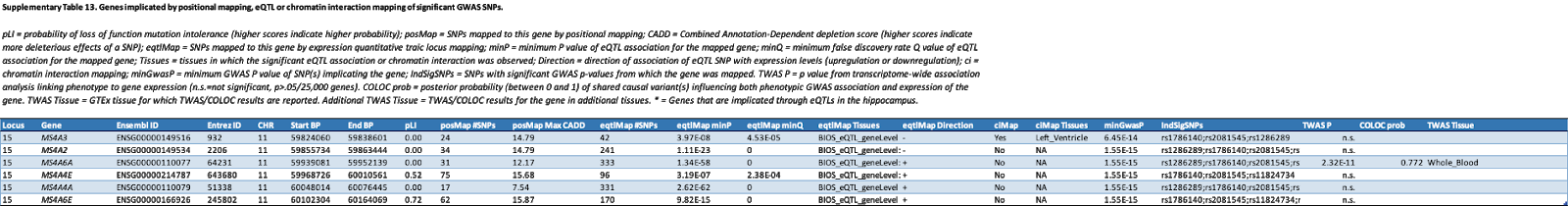

- Kunkle et al. (2019) Nat Genet. 51:414-30 [DOI:10.1038/s41588-019-0358-2](https://doi.org/10.1038/s41588-019-0358-2)
  - GWAS chips (Illumina and Affy) + 1000G EUR imputation
  - SNP-based analysis
    - rs7933202-C (11:59936926:A:C:hg19, gnomAD-NFE AF=0.38,

intergenic between MS4A2 and MS4A6A)

- Stage 1 (discovery, 9,456,058 common variants and 2,024,574 rare variants, genome-wide sig thresh P=5.0x10^-8^)

#### ADGC, CHARGE, EADI, GERAD

- 21,982 cases and 41,944 controls
- rs7933202-C
  - P = 2.2x10^-15^

## OR = 0.89 (0.86-0.92)

- Stage 2 (replication, 11,632 SNPs as in Lambert et al. (2013), Bonferroni-adjusted sig thresh P=4.3x10^-6^)
  - Other cohorts
  - 8,362 cases and 10,483 controls
  - 116 SNPs below sig thresh in stage 2, 80 of these below genome-wide sig thresh in stage 1 (only CD33 and DSG2 below genome-wide sig thresh in stage 1 did not replicate in stage 2)
  - rs7933202-C
    - P = 1.6x10^-5^

## OR = 0.90 (0.86-0.95)

- Meta-analysis of stage 1 and 2
  - 25,580 cases and 48,466 controls
  - rs7933202-C
    - P = 1.9x10^-19^

## OR = 0.89 (0.87-0.92)

- Locus-to-gene mapping (priority score system):
-
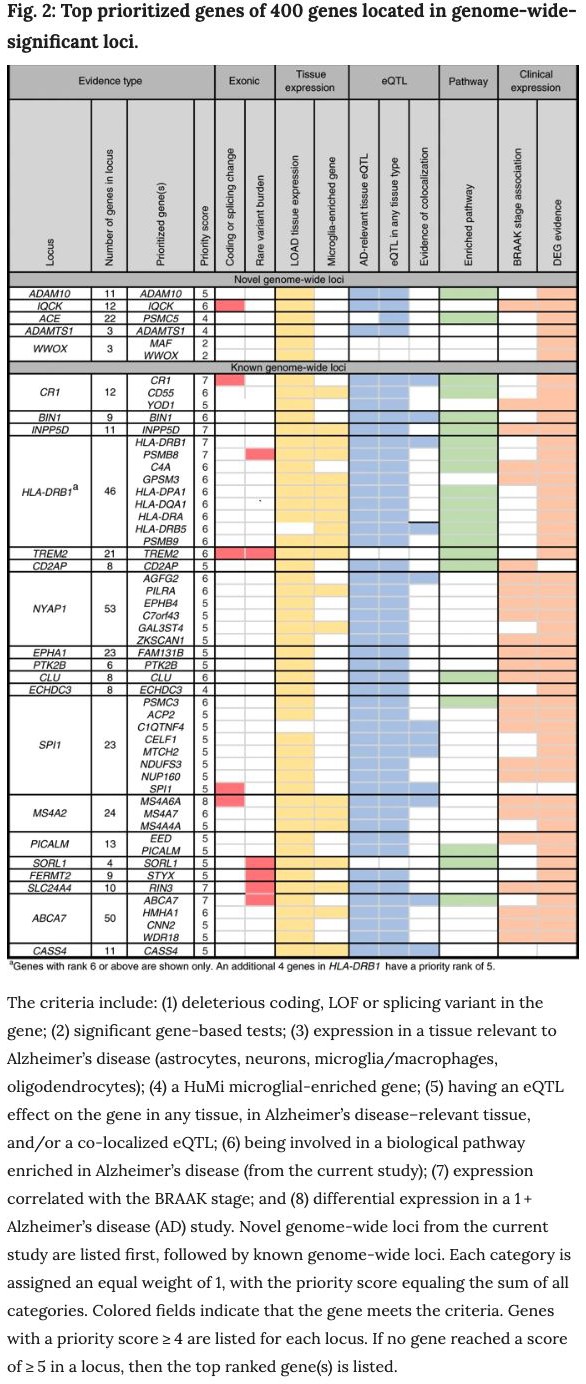

### 2. Pleiotropy

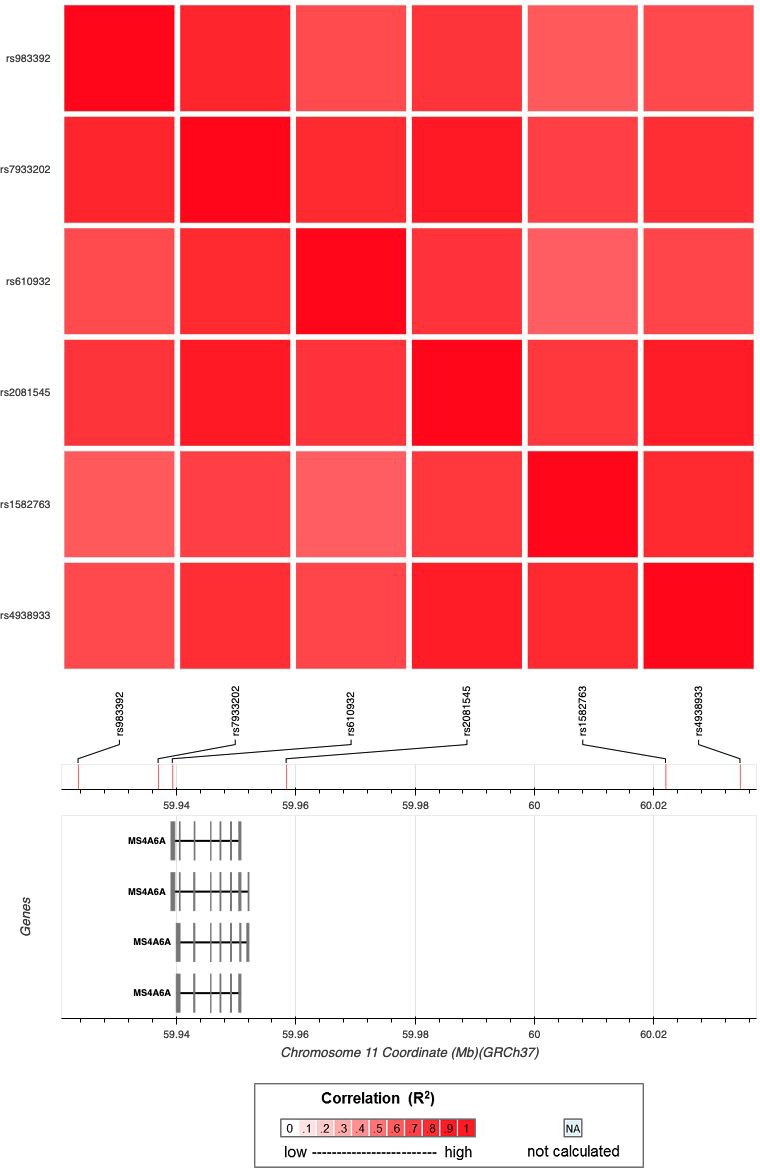

●

- Searched the above SNPs in various GWAS catalogs and noted phenotypes associated with those SNPs or SNPs in the region that are in high LD (R^2^ > 0.6) with them.
- GWAS Catalog:
- CSF sTREM2 levels (increased, rs1582763-A IGAP AD risk-decreasing allele)
  - Deming et al. (2019) Sci Transl Med. [DOI:10.1126/scitranslmed.aau2291](https://doi.org/10.1126/scitranslmed.aau2291)
- C-reactive protein levels (decreased, rs1582763-A IGAP AD risk-decreasing allele)
  - Ligthart et al. (2018) Am J Hum Genet. 103:691-706 [DOI:10.1016/j.ajhg.2018.09.009](https://doi.org/10.1016/j.ajhg.2018.09.009)
- Fibrinogen levels
- Factor VII activity or levels
- Heel bone mineral density
- Acute myeloid leukemia?
  - GWAS Atlas:
- Heel bone mineral density
- Myeloid white cell count
- Neutrophil count
- Lymphocyte percentage of white cells
  - Other:
- Age-at-onset defined survival (AAOS)
  - Huang, Marcora et al. (2017) Nat Neurosci. 20:1052-61 [DOI:10.1038/nn.4587](https://doi.org/10.1038/nn.4587)

### 3. Functional genomics

- Huang, Marcora et al. (2017) Nat Neurosci. 20:1052-61 [DOI:10.1038/nn.4587](https://doi.org/10.1038/nn.4587)
- AD risk alleles/SNP heritability is enriched in epigenomic annotations of myeloid lineage cells
- Coloc and SMR with monocyte and macrophage eQTLs implicates MS4A4A and MS4A6A as candidate causal genes
  - Novikova et al. (2019) bioRxiv <https://www.biorxiv.org/content/10.1101/694281v2>
- AD risk alleles/SNP heritability is enriched in **active** enhancer of monocytes, macrophages and microglia
- Integration of Alzheimer’s disease genetics and myeloid genomics (monocyte/macrophage/microglia epigenomic annotations, chromatin activity, chromatin interactions, eQTLs) implicates MS4A4A and MS4A6A as candidate causal genes
-
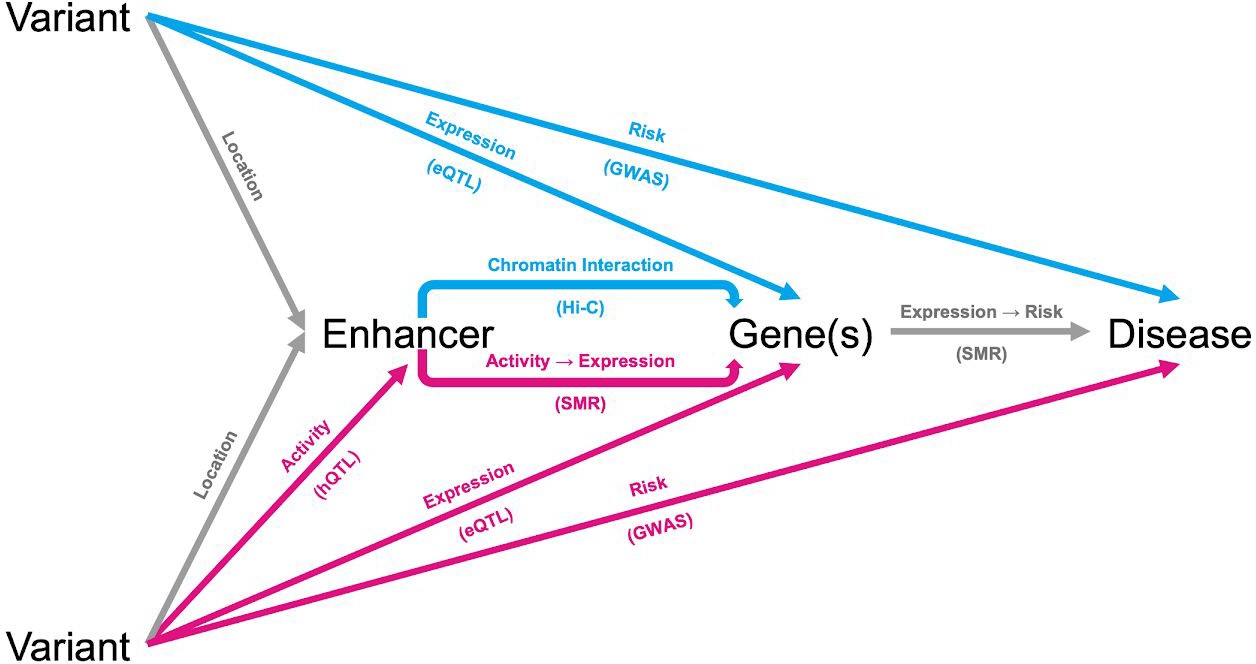

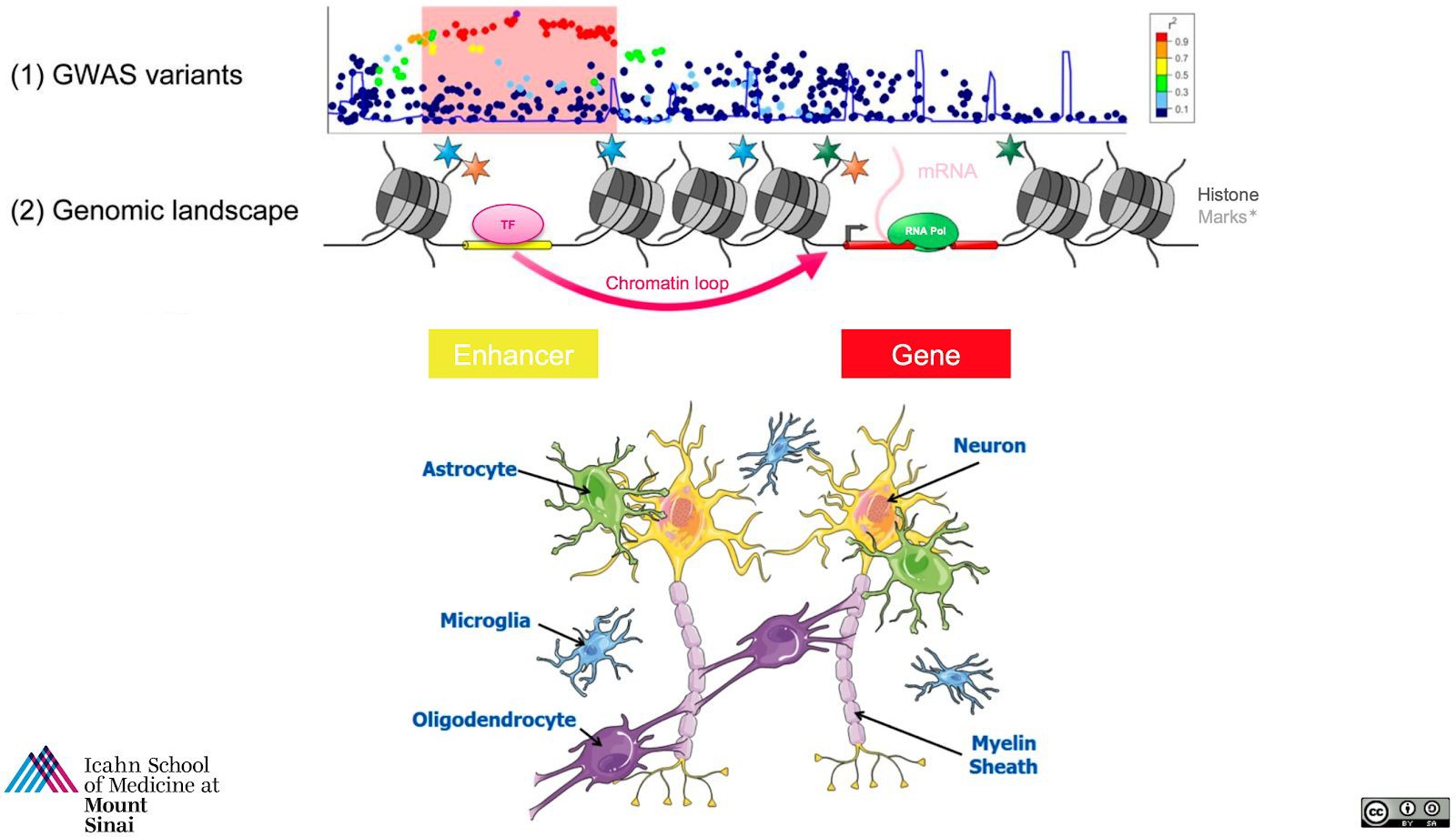

-
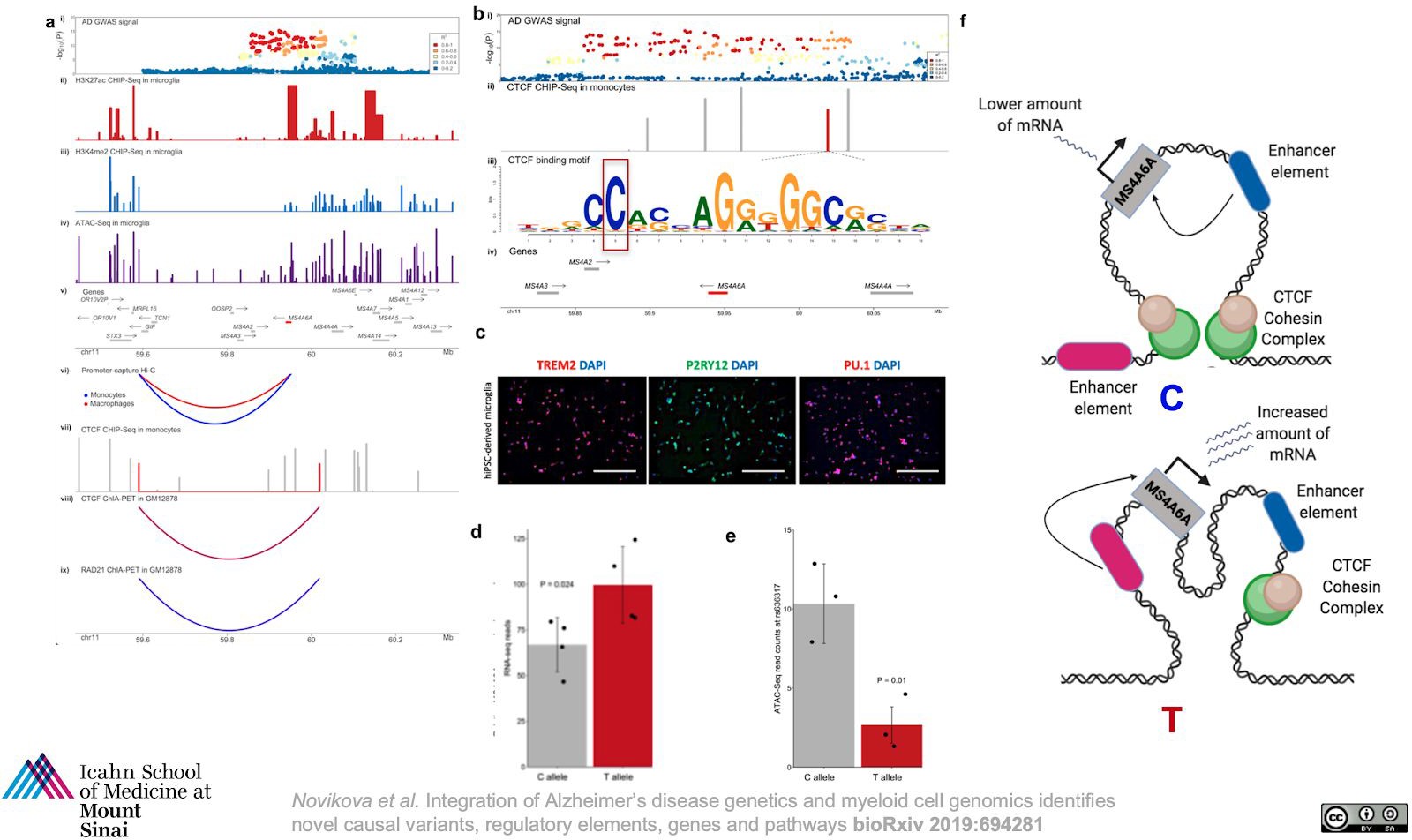

  - Nott et al. (2019) Science. 366:1134-9 [DOI:10.1126/science.aay0793](https://doi.org/10.1126/science.aay0793)
-
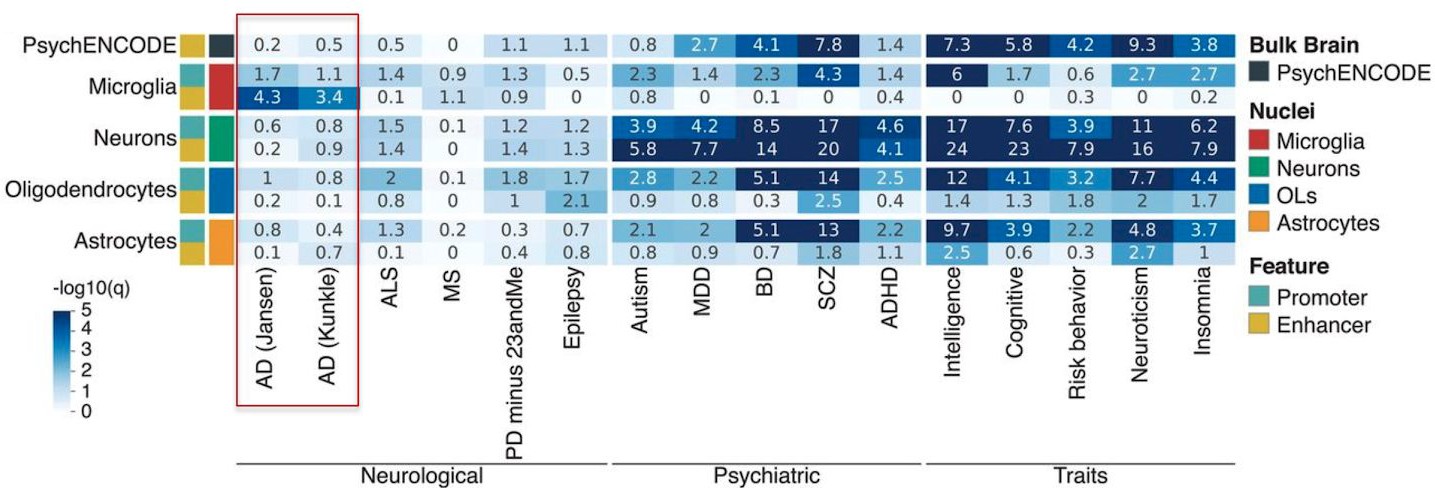
AD risk alleles/SNP heritability is enriched in microglial enhancers but not in enhancers or promoters of other brain cell types
- Mapping of microglial enhancers with promoter of target genes using PLAC-Seq implicates MS4A7 as a target gene
  - Ma et al. (2019) Aging Cell. 18:e12964 [DOI:10.1111/acel.12964](https://doi.org/10.1111/acel.12964)
- Sliding-window aggregation of AD risk effects of CpG-related single nucleotide polymorphisms (CGS, which may perturb DNA methylation → gene expression)
- Genome-wide significant associations with two windows at MS4A6A and two windows near MS4A4A. The total number of CGS-derived CpG dinucleotides in the window near MS4A4A was associated with AD risk (P = 2.67x10^-10^), brain DNA methylation (P = 2.15x10^-10^), and gene expression in brain (P = 0.03) and blood (P = 2.53x10^-4^)

### 4. Additional information

- Brain cell type-specific expression:

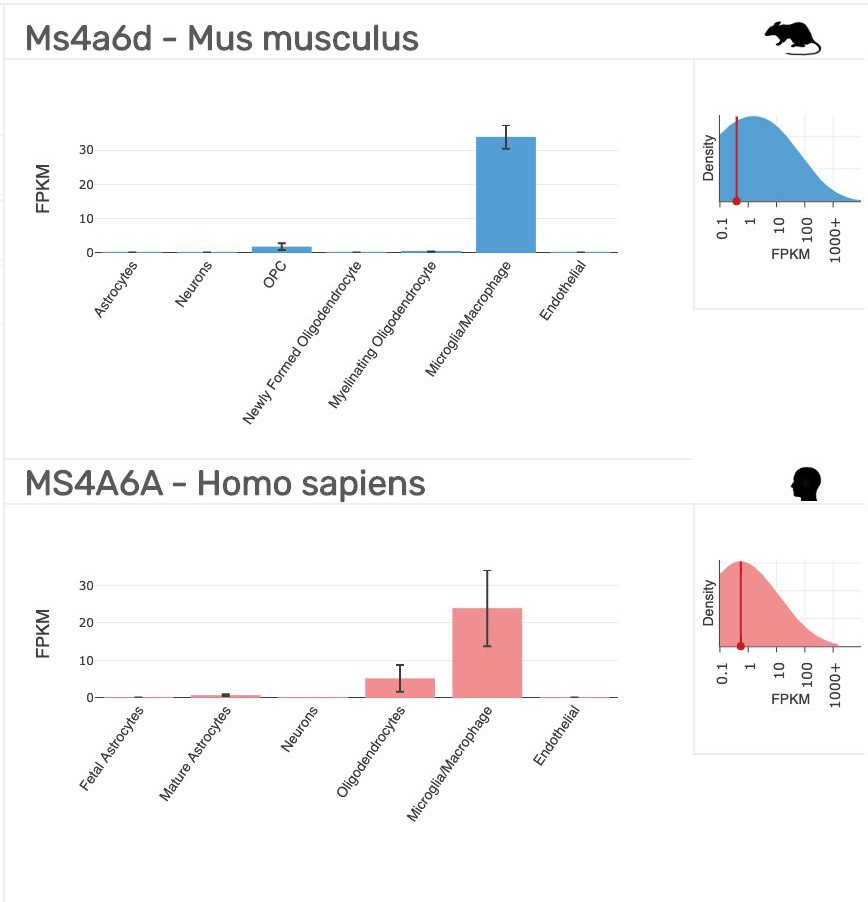

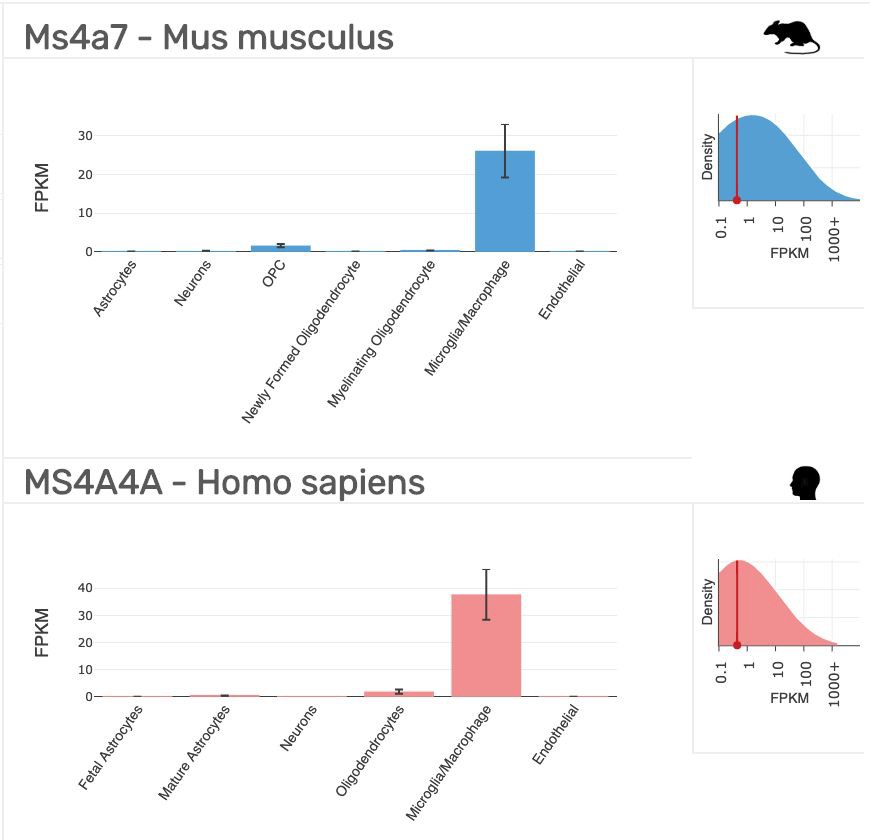

- Stronger association in APOE ɛ4- subjects (PMID:25778476)
- Interaction between variants in CLU and MS4A4E modulates AD risk (PMID:26449541)
- MS4A4A/6A interacts with CLEC7A/Dectin-1 in lipid rafts on the plasma membrane of macrophages and modulate ITAM signaling downstream of CLEC7A (PMID:31263276)
- MS4A4A interacts with TREM2 in lipid rafts on the plasma membrane of macrophages (PMID:31413141)
- Like TREM2, MS4A4A/6A are upregulated upon M2 polarization of macrophages, for example upon exposure to IL4 (PMID:31263276, PMID: 28303902)
- MS4A cluster genes are expressed in a subpopulation of olfactory receptor cells in the mouse and appear to mediate intracellular calcium responses upon exposure to lipid odorants (PMID:27238024), similar to TREM2 (another ITAM receptor that senses lipids)

### 5. Recommended tier

- Locus: Tier 1 (sufficient evidence)
- Gene(s): MS4A6A and MS4A4A Tier 1 or 2 (depending on tier definition, discussion needed! FTO locus as an example, see next page) and possibly other genes (e.g., MS4A7)

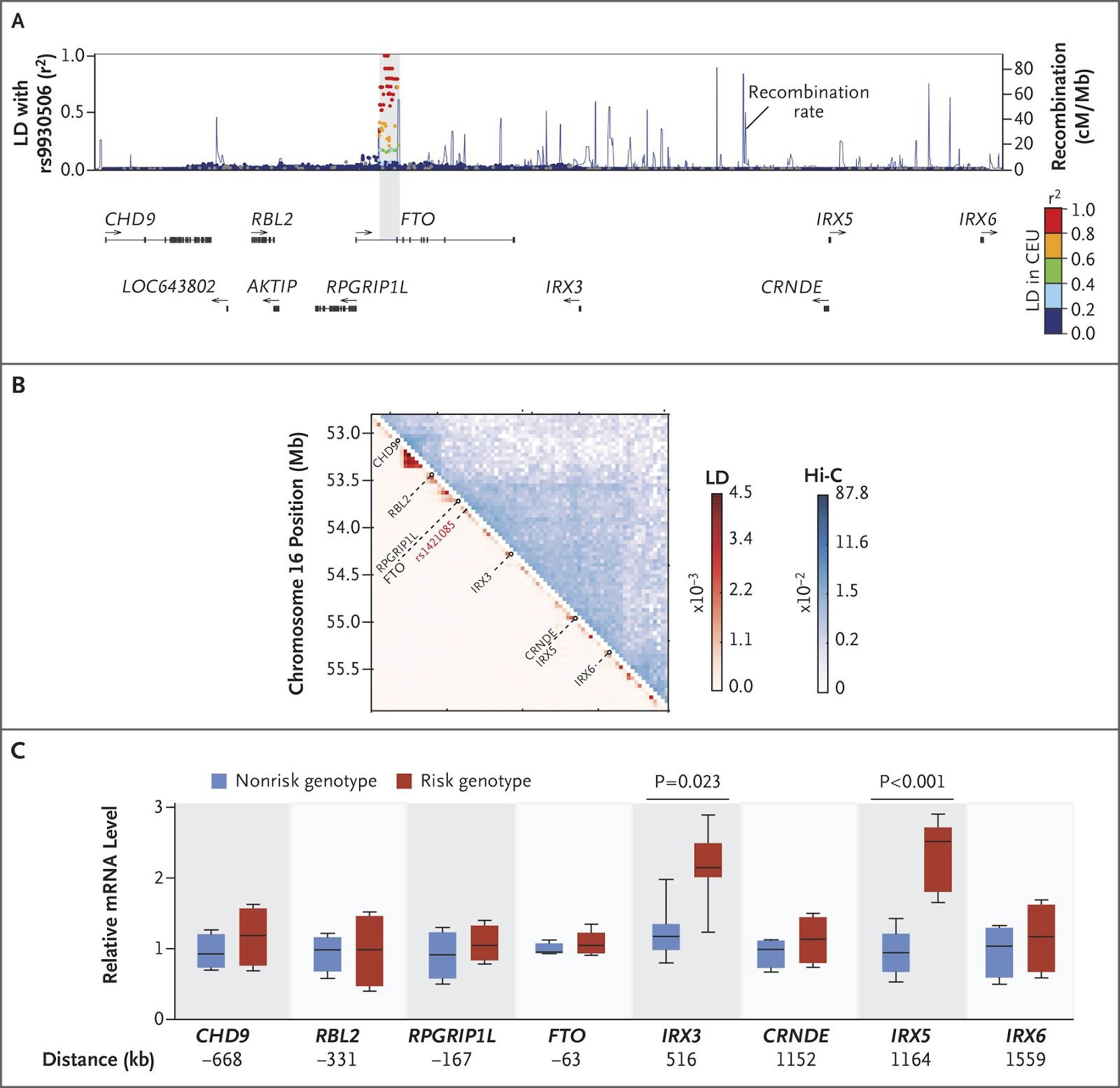

**Activation of IRX3 and IRX5 Expression in Human Adipocyte Progenitors by the FTO Obesity Risk Genotype.** Panel A shows gene annotations and LD with array tag variant rs9930506 in a 2.5Mb window; LD is expressed as r2 values in the CEU population. Arrows indicate the direction of transcription of annotated genes in the locus. Panel B shows chromosome conformation capture (Hi-C) interactions contact probabilities in human IMR90 myofibroblasts, revealing a 2-Mb topologically associating domain, and LD mean r2 statistics for all SNV pairs at 40-kb resolution. Panel C shows box plots for expression levels, after 2 days of differentiation, in human adipose progenitors isolated from 20 risk-allele carriers and 18 nonrisk-allele carriers, evaluated by means of a quantitative polymerase-chain-reaction analysis for all genes in the 2.5Mb locus. The horizontal line within each box represents the median, the top and bottom of each box indicate the 75th and 25th percentile, and I bars indicate the range.

Claussnitzer et al. (2015) N Engl J Med. 373:895-907 DOI:10.1056/NEJMoa1502214

*PLCG2*

Locus – Data to collect

Locus: **PLCG2**

Evaluator: Schellenberg

Date 9/23/2019

A. Significance

a. discovery study^1^

- study size (#cases/controls)
- Stage 1
- 16,097 cases/18,077 controls
- Exome chip genotyping
- P = 1.19 x 10^-5^; OR = 0.65 (rs72824905)
- MAF cases 0.006; MAF controls 0.011
  - evaluation of population heterogeneity (yes/no/comments)
- Yes
  - multiple test/model corrections (yes/no/comments)
- Single model
  - meta-analysis – discovery study^1^
  - study size (#cases/controls):
- Stage 2
- De novo genotyping and imputation, 43 variants
- 14,041 cases/ 21,921 controls
- P = 1.35 x 10^-4^; OR = 0.70 (rs72824905)
- MAF cases 0.006, MAF controls 0.007
- Stage 3
- Array genotypes used for imputation (some validated against exome chip data)
- 6,652 cases/8,345 controls
- P = 2.48 x 10^-2^; OR = 0.69 (rs72824905)
- MAF cases 0.006, MAF controls 0.007
- Meta-analyses(stages 1-3)
- 5.38 x 10^-10^; OR = 0.68
- MAF cases 0.006, MAF controls 0.009
  - evaluation of population heterogeneity (yes/no/comments)
- yes
  - multiple test/model corrections (yes/no/comments)
- single model

b. replication studies

- independent investigators and data^2^
- 4,985 cases/9,238 controls (completely independent of Discovery study^1^)
- Single SNV tested (rs72824905)
- Array data/Taq-Man data/imputed data
- MAF cases 0.0056/MAF controls 0.0102;
- P = 0.63 x 10^-4^; OR = 0.57 (0.41-0.78)
- Effect direction/approximate effect size same as discovery study
- evaluation of population heterogeneity (yes/no/comments); yes
- multiple test/model corrections (yes/no/comments); single model

c. other ethnic groups study size (#cases/controls)

- African Americans 180 cases/331 controls;
- Not significantly different (P = 0.35); study too small^3^
- Argentinian; 419 cases/486 controls^4^
- Not significant – too small

d. larger study with added samples

- Pleiotropy

e. AD-related phenotype (*e.g*. CSF biomarker association)

- Dementia (UK Biobank)^2^
- 16,968 father cases/358,468 father controls
- 32,262 mother cases/346,999 mother controls
- Single SNV (rs72824905) reported
- P = 1.8 x 10^-3^; OR = 0.88 (0.81 – 0.95)
- Same effect direction
  - Dementia with Lewy-bodies^2^
- 1,446 cases/5,286 controls
- Single SNV (rs72824905) reported
- P = 0.045, OR = 0.54 (0.30-0.99)

f. Frontotemporal dementia^2^

- 2,437 cases/10,647 controls
- Single SNV (rs72824905) reported
- P = 0.011, OR = 0.61 (0.41-0.89)

g. Non-AD phenotype (*e.g*. coronary artery disease, *APOE*)

- Longevitry^2^
- Subcortical brain volumes in young individuals^5^
- PLCG2 associated antibody deficiency and immune dysregulation (PLAID)^6-10^
  - Recommended tier (locus): tier 1

*PPARGC1A*

Locus – Data to collect

Locus: ***PPARGC1A***

Evaluator: **Naj**

Date **10/2/2019**

1. Significance

a. discovery study^1^

- study size (#cases/controls)
- Stage 1 (GERAD)
- 3,332 cases/9,832controls
- HRC-imputed GWAS from GERAD
- http://genomics.senescence.info/genes/entry.php?hgnc=PPARGC1A
- *P* = 2.2 × 10^−6^; β = 0.877, SE = 0.1851 [OR (95% CI): 2.40 (1.67, 3.45) ??]
- Conditioning on *APOE*: *P* = 1.0 × 10^−6^; β = 0.92, SE = 0.1885
  - evaluation of population heterogeneity (yes/no/comments)
- No- within GERAD, adjustment for principal components; for replication, used only summary statistics
  - multiple test/model corrections (yes/no/comments)
- Two model: 1) general, and 2) conditioned on *APOE*
- “Gene-wide significance” of *P* < 2.5 × 10^−6^
  - meta-analysis – discovery study^1^
  - study size (#cases/controls):
- None
- IGAP without GERAD was used to derive weights for gene-based testing and therefore not available as independent replication
  - evaluation of population heterogeneity (yes/no/comments)
- N/A
  - multiple test/model corrections (yes/no/comments)
- N/A

b. replication studies

independent investigators and data^2^

- None
- evaluation of population heterogeneity (yes/no/comments); **N/A**
- multiple test/model corrections (yes/no/comments); **N/A**

c. other ethnic groups study size (#cases/controls)

- Shibata et al. 2013 (PMID: 23741228)
- Japanese AD patients and controls
- 171 AD patients/136 age-matched controls/53 PDD patients;
- Four exonic SNPs of both genes (rs1801282 and rs3856806 of the PPAR-γ gene, rs3736265 and rs8192678 of the PGC-1α gene
- Haplotype analysis and logistic regression analysis with *APOE* status
- No significant association in single variant or haplotypic associations

d. larger study with added samples

**e.** Not observed in Escott-Price et al, 2014 (PMID: 24922517)

f. Pleiotropy

- AD-related phenotype (*e.g*. CSF biomarker association)
  - Cognitive impairment
- Li et al. 2015 (PMID: 26252872)
- ADNI cases/controls
- Top SNV association at rs41359445: P = 7 x 10^-8^

g. Non-AD phenotype (*e.g*. coronary artery disease, *APOE*)

| **Variant and risk allele** | **Location** | ***P*** | **Reported trait** | **PubMed ID** |
| --- | --- | --- | --- | --- |
| [rs768695-?](https://www.ebi.ac.uk/gwas/variants/rs768695) | 4:23797195 | 6E-6 | Reaction time | [29844566](https://www.ebi.ac.uk/gwas/publications/29844566) |
| [rs188794202-C](https://www.ebi.ac.uk/gwas/variants/rs188794202) | 4:23848945 | 1E-7 | Cognitive decline rate in late mild cognitive impairment | [26252872](https://www.ebi.ac.uk/gwas/publications/26252872) |
| [rs41359445-A](https://www.ebi.ac.uk/gwas/variants/rs41359445) | 4:23792721 | 7E-8 | Cognitive decline rate in late mild cognitive impairment | [26252872](https://www.ebi.ac.uk/gwas/publications/26252872) |
| [rs2970870-?](https://www.ebi.ac.uk/gwas/variants/rs2970870) | 4:23891394 | 2E-9 | Red blood cell count | [30595370](https://www.ebi.ac.uk/gwas/publications/30595370) |
| rs10517032-**C**, rs10517031-**A**, rs2324237-**A**, rs16874420-**C**, rs10020457-**G**, rs10517030-**A**, rs2324241-**A** | 4:23966759\|4:23966672\|4:23964419\|4:23968416\|4:23961338\|4:23961283\|4:23953895 | 3E-13 | Core binding factor acute myeloid leukemia | [27903959](https://www.ebi.ac.uk/gwas/publications/27903959) |
| rs744790-**T**, rs7680809-**C**, rs4269165-**T** | 4:24414550\|4:24412314\|4:24414228 | 9E-6 | Core binding factor acute myeloid leukemia | [27903959](https://www.ebi.ac.uk/gwas/publications/27903959) |
| [rs687706-?](https://www.ebi.ac.uk/gwas/variants/rs687706) | 4:24044033 | 2E-9 | Adolescent idiopathic scoliosis | [30019117](https://www.ebi.ac.uk/gwas/publications/30019117) |
| [rs17590046-T](https://www.ebi.ac.uk/gwas/variants/rs17590046) | 4:24360918 | 1E-9 | Essential tremor | [27797806](https://www.ebi.ac.uk/gwas/publications/27797806) |
| [rs12501032-G](https://www.ebi.ac.uk/gwas/variants/rs12501032) | 4:23949395 | 2E-15 | Resting heart rate | [27798624](https://www.ebi.ac.uk/gwas/publications/27798624) |
| [rs12501032-?](https://www.ebi.ac.uk/gwas/variants/rs12501032) | 4:23949395 | 1E-7 | Gout | [22179738](https://www.ebi.ac.uk/gwas/publications/22179738) |
| [rs59143394-A](https://www.ebi.ac.uk/gwas/variants/rs59143394) | 4:24494344 | 2E-6 | Neuroticism | [29255261](https://www.ebi.ac.uk/gwas/publications/29255261) |
| [rs34751092-A](https://www.ebi.ac.uk/gwas/variants/rs34751092) | 4:24127414 | 6E-7 | Corneal astigmatism | [29422769](https://www.ebi.ac.uk/gwas/publications/29422769) |
| [rs73243607-C](https://www.ebi.ac.uk/gwas/variants/rs73243607) | 4:23757039 | 3E-8 | Estimated glomerular filtration rate | [31015462](https://www.ebi.ac.uk/gwas/publications/31015462) |
| [rs16875255-?](https://www.ebi.ac.uk/gwas/variants/rs16875255) | 4:24183778 | 6E-6 | Parental longevity (combined parental age at death) | [27015805](https://www.ebi.ac.uk/gwas/publications/27015805) |
| [rs11730701-G](https://www.ebi.ac.uk/gwas/variants/rs11730701) | 4:24130930 | 1E-6 | Schizophrenia | [26198764](https://www.ebi.ac.uk/gwas/publications/26198764) |
| [rs16875528-T](https://www.ebi.ac.uk/gwas/variants/rs16875528) | 4:24306456 | 9E-6 | Sense of smell | [26632684](https://www.ebi.ac.uk/gwas/publications/26632684) |
| [rs7698250-T](https://www.ebi.ac.uk/gwas/variants/rs7698250) | 4:24514008 | 2E-10 | Pulmonary emphysema | [24383474](https://www.ebi.ac.uk/gwas/publications/24383474) |
| [rs7679088-G](https://www.ebi.ac.uk/gwas/variants/rs7679088) | 4:24244513 | 4E-7 | Facial morphology (factor 20) | [28441456](https://www.ebi.ac.uk/gwas/publications/28441456) |
| [rs17650401-T](https://www.ebi.ac.uk/gwas/variants/rs17650401) | 4:24374210 | 2E-6 | Molar-incisor hypomineralization | [23918034](https://www.ebi.ac.uk/gwas/publications/23918034) |
| [rs372822970-T](https://www.ebi.ac.uk/gwas/variants/rs372822970) | 4:23916937 | 1E-56 | Pulse pressure | [30578418](https://www.ebi.ac.uk/gwas/publications/30578418) |
| [rs648674-?](https://www.ebi.ac.uk/gwas/variants/rs648674) | 4:24024858 | 9E-16 | Lung function (FEV1/FVC) | [30595370](https://www.ebi.ac.uk/gwas/publications/30595370) |
| [rs7667050-T](https://www.ebi.ac.uk/gwas/variants/rs7667050) | 4:23811486 | 3E-14 | Estimated glomerular filtration rate | [31152163](https://www.ebi.ac.uk/gwas/publications/31152163) |
| [rs4697428-T](https://www.ebi.ac.uk/gwas/variants/rs4697428) | 4:23933430 | 3E-6 | Coffee consumption | [31046077](https://www.ebi.ac.uk/gwas/publications/31046077) |

- Recommended tier (locus): **tier 3**

*PSEN1*

Genes – rare coding variants

Locus – *PSEN1*

Evaluator:	Blue

Date:	12/05/2019

A. Primary evidence

1. Linkage analyses

Multiple studies of familial, early-onset Alzheimer’s disease mapped the locus containing *PSEN1*.

- Mullan *et al.* (1992): multipoint maxLOD > 7 at D14S43 at ϴ = 0 across 9 families (PMID: 1303291).
- Schellenberg *et al.* (1992): maxLOD > 9 at D14S43 at ϴ = 0.01 across 9 non-Volga German families (PMID: 1411576).
- St. George-Hyslop *et al.* (1992): multiethnic study of 21 families with mean AAO between 42-84yrs, two-point maxLOD > 20 at D14S43 at ϴ = 0.05 (PMID: 1303289).
- Van Broeckhoven *et al.* (1992): two-point maxLOD > 13 at D14S43 at ϴ = 0 across 2 Belgian families (PMID: 1303290).

2. Non-synonymous SNV (CADD > 20?)

100s of non-synonymous variants that segregate with disease in early-onset AD. Initial report by Sherrington *et al.* (1995) described five different missense variants co-segregating with early-onset AD (PMID:7596406). Most if not all of these variants are missense variants with high CADD scores.

3. Altered function SNV (stop gain, stop loss, splicing)

There are a couple splicing variants reported as pathogenic or likely pathogenic in ClinVar.

4. Structural variants – disrupt expression

Small indels and genomic deletions have been reported (PMID: 27016693).

5. Significant association evidence with AD

Tons of case-control association studies and family-based studies with conflicting evidence (most tested single variant; 20 positive, 42 negative; <http://www.alzgene.org/>, PMID:17998437).

B. Replication in another population

1. Yes. For example, the Columbian founder mutation G280A (PMID: 9052708), Calabria, Naples, Australia (PMID: 20164095), Mexico (PMID: 16628450), Mexico and USA (PubMed: 16897084), etc.

2. Gene expressed in an AD-relevant tissue/cell type

*PSEN1* is broadly expressed, some of the highest levels of expression are observed in the brain (<https://gtexportal.org/home/gene/PSEN1>).

C. Supporting evidence

1. Multiple rare variants significantly associated with AD

Yes, see above. Also, some rare-variant enrichment/burden testing results show association between *PSEN1* and late-onset AD (ex. PMID: 22312439).

2. Common variants associated with the same gene

Mixed evidence, see above.

3. Pleiotropy

Common variants in *PSEN1* have been associated with highest math class taken (PMID: 30038396), nominal evidence of association with multiple AD biomarkers (<https://beta.niagads.org/>; p < 0.001 with cerebral amyloid angiopathy and hippocampal sclerosis (NG00041), CSF tau (NG00055), and late-onset AD risk in IGAP ε4 non-carriers (NG00078)).

4. Rare variants in the same gene in another ethnic group

Yes

5. Co-segregation in families

Yes

6. Other?

Animal and cell models carrying *PSEN1* mutations show altered APP metabolism and related phenotypes (ex., PMID 28304309, PMID:29153989, PMID: 29191219; <https://monarchinitiative.org/gene/HGNC:9508#ortholog-phenotypes>).

*PSEN2*

Genes – rare coding variants

Locus – *PSEN2*

Evaluator:	Blue

Date:	12/05/2019

A. Primary evidence

1. Linkage analyses

Multipoint linkage to D1S479 with maxLOD > 4 at ϴ = 0.10 in 7 Volga German families with autosomal dominant early-onset AD (PMID: 7638621) subsequently mapped to PSEN2 (PMID: 7638622).

2. Non-synonymous SNV (CADD > 20?)

Over a dozen pathogenic variants have been reported for *PSEN2* and AD. Most of these are missense. (PMID: 22581678, PMID: 20375137, <https://www.alzforum.org/mutations/>).

3. Altered function SNV (stop gain, stop loss, splicing)

Many of those missense variants appear to result in partial loss of function, and there is a frameshift causing an early termination reported (PMID:15663477).

4. Structural variants – disrupt expression

There is a large deletion in ClinVar reported as pathogenic (<https://www.ncbi.nlm.nih.gov/clinvar/variation/533784/>).

5. Significant association evidence with AD

Multiple mutations with evidence of segregation with AD in autosomal dominant, early-onset AD (above).

6. Replication in another population

Discovery in Volga Germans (PMID: 7638622), later observed different variants in Italian families (PMID: 7651536), Spain (PMID: 12925374), etc..

7. Gene expressed in an AD-relevant tissue/cell type

*PSEN2* is broadly expressed in the body including the brain.

B. Supporting evidence

1. Multiple rare variants significantly associated with AD

>10 variants reported to be associated with AD (PMID: 20375137, <https://www.alzforum.org/mutations/>).

2. Common variants associated with the same gene

Several case-control studies with conflicting evidence (most tested one variant; 2 positive, 10 negative; <http://www.alzgene.org/geneoverview.asp?geneid=55>). Nominal association with late-onset AD in a transethnic GWAS (PMID: 28183528).

3. Pleiotropy – similar to AD only

There is some nominal association with late-onset AD and biomarkers of AD (variants with p < 0.001 with hippocampal sclerosis of the elderly; PMID: 25188341). Also see association between *PSEN2* missense variants and age-at-onset of AD (PMID: 22312439).

4. Rare variants in the same gene in another ethnic group

Yes

5. Co-segregation in families

Yes

6. Other?

Human cell models of *PSEN2* mutations show altered APP metabolism; reversal of the *PSEN2* mutation ameliorated the phenotype (PMID: 29078805; several abstracts for AAIC in *Alzheimer’s and Dementia* from last few years that haven’t made it to publication yet).

*RORA*

Locus – Data to collect

Gene:	**RORA**

Evaluator: **Farrer**

Date: **10/2/19**

A, Significance

1. discovery study

- study size (#cases/controls) -- **3,332 cases, 9,832 controls (GERAD)^a^**
- evaluation of population heterogeneity (yes/no/comments) – **presumably yes**
- multiple test/model corrections (yes/no/comments) – **yes (gene-based p = 7.4x10^-7^ which is gene-wide significant after correcting for 18,087 tests, i.e., threshold p = 2.76x10^-6^ )**

2. meta-analysis – discovery study -- **N/A**

- study size (#cases/controls)
- evaluation of population heterogeneity (yes/no/comments)
- multiple test/model corrections (yes/no/comments)

3. replication studies – NO REPLICATION STUDIES, however result available from another study including UK Biobank (proxy phenotype) and IGAP (GERAD portion overlaps with discovery study) with a total sample of 314,278^b^

- gene-based p =0.87 (not significant)
- independent investigators and data - RARE
- larger study with added samples

4. replication studies – evidence **--** N/A

- signal direction
- other ethnic groups study size (#cases/controls)
- evaluation of population heterogeneity (yes/no/comments)
- multiple test/model corrections (yes/no/comments)
  - Pleiotropy

5. AD-related phenotype (*e.g*. CSF biomarker association)

- *RORA* expression upregulated in hippocampus^c^

6. on-AD phenotype (*e.g*. coronary artery disease, *APOE*)

- *RORA* implicated in PTSD^d,e^, autism^f^ & age-related macular degeneration^g,h^
- *RORA* has roles in immunity, cerebellum development, lipid metabolism , and inflammation^i-k^
- Recommended tier – no higher than tier 3, perhaps lower

Supporting references –

*SORL1*

Evaluator: Mayeux

Date 11/14/19

A. Primary evidence

Locus	MAF	meta p

rs11218343	.039	9.7 x 10^-15^

Lambert JC. Meta-analysis of 74,046 individuals identifies 11 new susceptibility loci for Alzheimer's disease. Nat Genet. 2013 Dec;45(12):1452-8. doi: 10.1038/ng.2802. Epub 2013 Oct 27. PubMed PMID: 24162737; PubMed Central PMCID: PMC3896259.

Kunkle BW. Genetic meta-analysis of diagnosed Alzheimer's disease identifies new risk loci and implicates Aβ, tau, immunity and lipid processing. Nat Genet. 2019 Mar;51(3):414-430. doi:

10.1038/s41588-019-0358-2. Epub 2019 Feb 28. Erratum in: Nat Genet. 2019

Sep;51(9):1423-1424. PubMed PMID: 30820047; PubMed Central PMCID: PMC6463297.

1. Altered function SNV (stop gain, stop loss, splicing)

Stop gain variants associated with both early and late onset AD

2. Structural variants – disrupt expression

None

3. Significant association evidence with AD

See above

4. Replication in another population

Multiple times

5. Gene expressed in an AD-relevant tissue/cell type

SORL1 is expressed in neurons, astrocytes and microglia

**B. Supporting evidence**

**Multiple rare variants significantly associated with AD**

**C. Pleiotropy**

A variant in SORL1 was recently described in a patient with clinically diagnosed FTLD (Ciani M et al Int J Mol Sci. 2019 Aug 10;20(16). pii: E3903. doi: 10.3390/ijms20163903.

**D. Rare variants in the same gene in another ethnic group:** African Americans, Caribbean Hispanic, Korean, Japanese.

1. Cuenco K, et al. Association of distinct variants in SORL1 with cerebrovascular and neurodegenerative changes related to Alzheimer disease. Arch Neurol. 2008 Dec;65(12):1640-8. doi:10.1001/archneur.65.12.1640. PubMed PMID: 19064752; PubMed Central PMCID:
2. PMC2719762.
3. Lee JH, Cheng R, Schupf N, Manly J, Lantigua R, Stern Y, Rogaeva E, Wakutani Y, Farrer L, St George-Hyslop P, Mayeux R. The association between genetic variants in SORL1 and Alzheimer disease in an urban, multiethnic, community-based cohort. Arch Neurol. 2007 Apr;64(4):501-6. PubMed PMID: 17420311; PubMed Central PMCID: PMC2639214.
4. Miyashita A. SORL1 is genetically associated with late-onset Alzheimer's disease in Japanese, Koreans and Caucasians. PLoS One. 2013;8(4):e58618. doi: 10.1371/journal.pone.0058618. Epub 2013 Apr 2. Erratum in: PLoS One. 2013;8(7). doi:10.1371/annotation/fcb56ea7-d32a-4e45-818d-39cef330c731. Moriaha, Takashi [corrected to Morihara, Takashi]. PubMed PMID: 23565137; PubMed Central PMCID: PMC3614978.
5. Vardarajan BN. Coding mutations in SORL1 and Alzheimer disease. Ann Neurol. 2015 Feb;77(2):215-27. doi: 10.1002/ana.24305. PubMed PMID: 25382023; PubMed Central PMCID: PMC4367199.

**E. Co-segregation in families**

1. Thonberg H et al. Acta Neuropathol Commun. (2017)
2. Gómez-Tortosa E et al. J Alzheimers Dis. (2018)

**F. Other**

1. Dodson S, et al. Loss of LR11/SORLA Enhances Early Pathology in a Mouse Model of Amyloidosis: Evidence for a Proximal Role in Alzheimer's Disease. J Neurosci 2008
2. Andersen OM. Risk factor *SORL1*: from genetic association to functional validation in Alzheimer’s disease. Acta Neuropath 2016

*SPI1*

Locus – CELF1

Gene: SPI1 and potentially others

Evaluator: Alison Goate and Edoardo Marcora

Date: January 3, 2020

D. Significance

A, Discovery study: IGAP, Lambert JC 2013

1. study size (#cases/controls)

2. Overall: IGAP

3. 17,008 cases / 37,154 controls

4. GWAS chip with 1000 Genome Project (2010 interim release) imputation

5. SNP-based analysis, top SNP rs10838725-C

- **Stage 1 (p=6.7x10^-6^) overall (P = 1.1x10^-8^ )**
- OR = 1.08 (1.05-1.11)

B. Meta-analysis of AD - IGAP Kunkle 2019 (N=74,046) stage 1 and overall GWS

1. Stage 1 N=63926; overall N= 82,771 participants
- GWAS chip with HRC imputation
- Top SNP rs3740688-G **p=5.4x10^-13^** (overall), **p=9.7x10^-11^** (stage1)
- OR=0.92 (0.89-0.94)

C. Meta-analysis of dementia– IGAP summary statistics plus UK Biobank (Marioni 2018)

1. 314,278 participants

27,696 maternal cases, 14,338 paternal cases, proxy for AD genetic study Top SNP rs12292911 p=3.3x10^-9^

2. MAGMA gene-based analysis for UK Biobank and IGAP meta-analysis summary results p=8.8x10^-7^

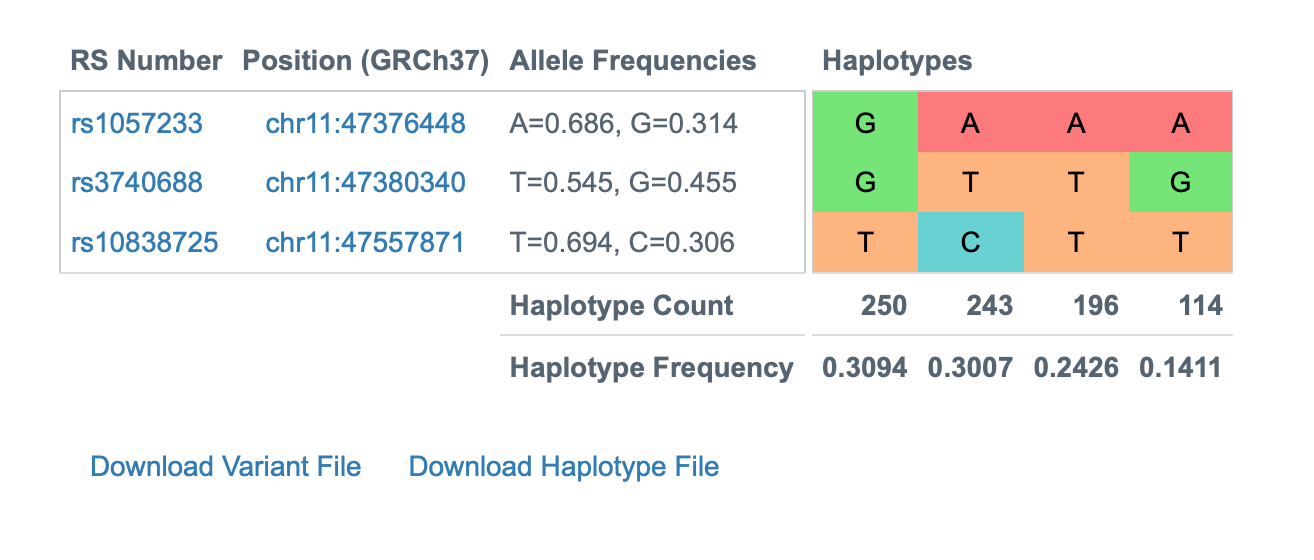

**Summary**: SNP level analyses

AD – IGAP SNP finding replicated in Kunkle 2019 p=5.4 x 10-13 (tier 1)

Dementia – IGAP SNP finding replicated in UK Biobank p=3.3 x 10-9 (tier 1)

Magma Gene-based analyses

AD - ?

Dementia –UK Biobank p=8.8x10-7

Note: Odds ratios flipped between IGAP and Kunkle but this reflects SNPs in high LD with flipped minor alleles (see figure)

D. Pleiotropy

1. AD-related phenotype (*e.g*. CSF biomarker association) Age at onset of AD
2. Huang et al., 2017 : <https://www.nature.com/articles/nn.4587.pdf>
3. 14,406 cases and 25,849 controls
4. Top SNP rs1057233-G from GWAS survival analysis
5. P=8.4x10^-6^
6. HR=0.94 (0.81-0.97)
7. Suggestive association with age at onset in subset of IGAP with AAO/ALA data

E. Non-AD phenotypes (*e.g*. coronary artery disease, *APOE*)

rs1057233 is associated with many traits particularly metabolic traits, psychiatric traits and height which all have GWAS signals <1x10^-10^ <https://atlas.ctglab.nl/PheWAS>

F, Functional Genomics

1. Enrichment of AD risk SNPs in myeloid active enhancers and cis-eQTL effects
2. Huang et al., 2017 : <https://www.nature.com/articles/nn.4587.pdf>
3. Raj et al., 2014: <https://www.ncbi.nlm.nih.gov/pubmed/24786080>
4. Novikova et al., [https://www.biorxiv.org/content/10.1101/694281v2](https://www.biorxiv.org/content/10.1101/694281v2" \o "https://www.biorxiv.org/content/10.1101/694281v2)
5. Tansey KE et al., 2018: <https://www.ncbi.nlm.nih.gov/pubmed/29482603>
6. Nott A, et al., 2019: [https://www.ncbi.nlm.nih.gov/pubmed/31727856](https://www.ncbi.nlm.nih.gov/pubmed/31727856" \o "https://www.ncbi.nlm.nih.gov/pubmed/31727856)

G. eQTLs in macrophages and monocytes

1. rs1057233-G
2. SPI1 monocytes p=1.50 × 10−105 ß= –1.11
3. SPI1 macrophages p=6.41 × 10−87 ß= –1.11
4. MYBPC3 monocytes p=4.99 × 10−51 –0.83
5. MYBPC3 macrophages p=5.58 × 10−23 –0.62
6. NUP160 macrophages 5.35 × 10−22 –0.62

H. Colocalization of eQTL signal and AD GWAS signal

1. SPI1 has ppH4> 0.8 in coloc for both MC and MP
2. MYBPC3 has ppH4>0.8 in coloc for MP only

I. SMR analysis

1. SPI1 and MYBPC3 significant in both MC and MP
2. NUP160 significant in MP only

J. SPI1 cistrome enriched for AD risk alleles in myeloid cells, experimental demonstration that manipulation of SPI1 expression results in change sin gene expression of other AD risk genes

1. Huang et al., 2017: <https://www.nature.com/articles/nn.4587.pdf>
2. Raj et al., 2014: <https://www.ncbi.nlm.nih.gov/pubmed/24786080>
3. Novikova et al., [https://www.biorxiv.org/content/10.1101/694281v2](https://www.biorxiv.org/content/10.1101/694281v2" \o "https://www.biorxiv.org/content/10.1101/694281v2)
4. Tansey KE et al., 2018: <https://www.ncbi.nlm.nih.gov/pubmed/29482603>
5. Nott A, et al., 2019: [https://www.ncbi.nlm.nih.gov/pubmed/31727856](https://www.ncbi.nlm.nih.gov/pubmed/31727856" \o "https://www.ncbi.nlm.nih.gov/pubmed/31727856)

Summary: Functional genomics provide evidence to support SPI1 from multiple datasets and cell types but also some evidence for MYBPC3 and NUP160

Recommended tier. Tier 1 for the locus, Tier 1 gene (SPI1) could be more than one signal and causal gene in the locus. No rare coding variants reported in any gene in this region to date.

Supporting references

- <https://www.nature.com/articles/nn.4587.pdf>
- <https://www.ncbi.nlm.nih.gov/pubmed/30820047>
- <https://www.nature.com/articles/ng.2802>
- <https://www.nature.com/articles/s41398-018-0150-6#MOESM2>

*TP53INP1*

***chr 8* Tier 4: Insufficient evidence to determine whether an association exists**

A) p = 1.4×10^−6^

<https://www.ncbi.nlm.nih.gov/pubmed/24922517>

International Genomics of Alzheimer's Project Consortium, comprising over 7 m genotypes from 25,580 Alzheimer's cases and 48,466 controls.

Based on IGAP p-values; not individuals level data; Fisher test

SNP filters: autosomal SNPs, (MAF) ≥0.01 and imputation quality >=0.3 in each individual study

B) P=7.8E-02 MAGMA gene-based associations for the UK Biobank and IGAP meta-analysis summary results <https://www.nature.com/articles/s41398-018-0150-6>
C) Not replicated by <https://www.sciencedirect.com/science/article/pii/S2352396418304122>
D) Listed in GWAS catalog for diabetes; not AD

Locus – Data to collect

Gene Based: TP53INP1

Evaluator: DeStefano

Date:	01/16/20

A. Significance

1. Mega meta-analysis – discovery study^1^

- study size (#cases/controls)
- Stage 1 IGAP GWAS
- Imputed with Impute2 or MACH using 1000G
- 17,008 cases/37,154 controls
- rs4735333 (OR = 1.056, p-value = 0.00044), rs1713669 (OR = 0.945, p-value = 0.0004104), rs896855 (OR = 1.054, p-value = 0.00084). Gene-wide p-value = 1.7 x 10^-2^
  - MAF cases not mentioned, MAF controls not mentioned
  - evaluation of population heterogeneity (yes/no/comments)
- not mentioned
  - multiple test/model corrections (yes/no/comments)
- Loci with p-value (combined over all SNPs at the locus) < 10^-4^ were selected for replication

2. Replication studies

- Replication within discovery manuscript^1^
- Stage 2
- 8,572 cases/11,312 controls of European ancestry
- Genotyped
- MAF cases not mentioned /MAF controls not mentioned;
- rs4735333 (OR = 1.099, p-value = 2.99E-5), rs1713669 (OR = 0.916, p-value = 0.00018), rs896855 (OR = 1.102, p-value = 3.21E-5). Gene-wide p-value = 4.5 x 10^-3^
- Effect direction/approximate effect size same as discovery
- evaluation of population heterogeneity (yes/no/comments); not mentioned
- multiple test/model corrections (yes/no/comments); yes multiple testing
- MAGMA gene-based associations for the UK Biobank and IGAP meta-analysis summary result (p-value = 5.3E-4) by Marioni et al.^2^
- Not replicated in a separate study using IGAP data by Kwok et al.^3^
- other ethnic groups study size (#cases/controls)
- Only done on European ancestry^1,2^
  - Combined gene-wide

3. Discovery + Replication ^1^

- Combined Gene-wide P-value from Fisher’s method = 1.4 x 10^-6^

4. Pleiotropy

5. AD-related phenotype (*e.g*. CSF biomarker association)

6. Non-AD phenotype (*e.g*. coronary artery disease, *APOE*)

- Type 2 diabetes^4^
- Eosinophil count^5^

7. Recommended tier:

- locus: 1 or 2
- Gene: 3.5

Note: Supplemental Table 4 from Lambert et al^6^ lists NDUFAF6 with p= 1.6x10-5 stage 1, p= 1.4x10-3 Stage 2, and p= 8.0x10-8 overall. This is in the region of TP53INP1 as shown in the plot from Novikova et al in BioRxiv <https://www.biorxiv.org/content/10.1101/694281v2.full>

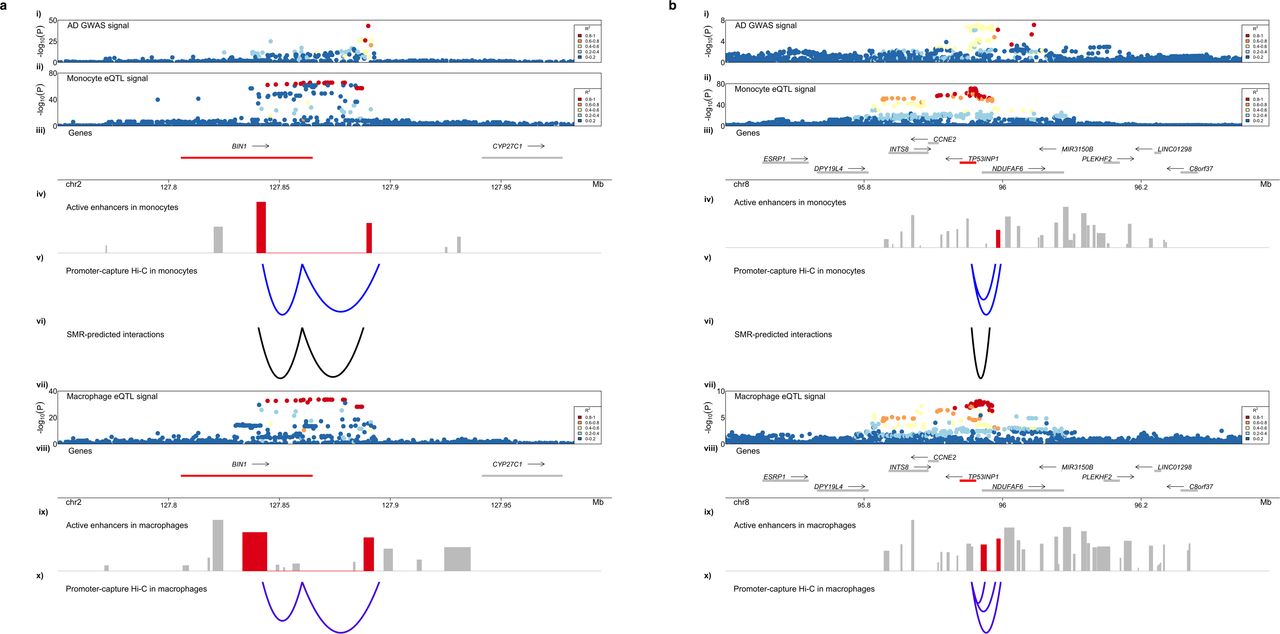

Additional Note: Except in the gene based test of Escott-Price et al (1) this locus met suggestive but not genome-wide significance in GWAS analysis, until Moreno-Grau et al 2019 (7) analyzed the IGAP I&II data in concert with the GRACE data achieving p= 2.54x10-8. However p=.40 in their replication data set. They note this SNP, rs10098778, is in the region of TP53INP1 but that the closest gene is NDUFAF6.

Stage I = GRACE (N=4120 cases; 3289 controls) plus IGAP I&II: OR=0.94 p = 2.54x10-8

Stage II = Spanish cohort (N=1943 cases; 3016 controls) OR=0.96 p=0.40

Stage I and II: OR=0.94 p=2.32x10-8

*TREM2*

Genes – rare coding variants

Locus – TREM2

Evaluator: Goate

Date: 9/25/2019

A. Primary evidence

1. Non-synonymous SNV (CADD > 20?)

R47H variant shows genome wide significant association with AD in European samples

a. Guerreiro R, Wojtas A, Bras J, Carrasquillo M, Rogaeva E, Majounie E, Cruchaga C, Sassi C, Kauwe JS, Younkin S, Hazrati L, Collinge J, Pocock J, Lashley T, Williams J, Lambert JC, Amouyel P, Goate A, Rademakers R, Morgan K, Powell J, St George-Hyslop P, Singleton A, Hardy J, Alzheimer Genetic Analysis Group. [TREM2 variants in Alzheimer's disease](https://www.alzforum.org/papers/trem2-variants-alzheimers-disease). *N Engl J Med*. 2013 Jan 10;368(2):117-27. Epub 2012 Nov 14 [PubMed](http://www.ncbi.nlm.nih.gov/pubmed/23150934).

b. Jonsson T, Stefansson H, Steinberg S, Jonsdottir I, Jonsson PV, Snaedal J, Bjornsson S, Huttenlocher J, Levey AI, Lah JJ, Rujescu D, Hampel H, Giegling I, Andreassen OA, Engedal K, Ulstein I, Djurovic S, Ibrahim-Verbaas C, Hofman A, Ikram MA, van Duijn CM, Thorsteinsdottir U, Kong A, Stefansson K. [Variant of TREM2 associated with the risk of Alzheimer's disease](https://www.alzforum.org/papers/variant-trem2-associated-risk-alzheimers-disease). *N Engl J Med*. 2013 Jan 10;368(2):107-16. Epub 2012 Nov 14 [PubMed](http://www.ncbi.nlm.nih.gov/pubmed/23150908).

2. Altered function SNV (stop gain, stop loss, splicing)

Stop gain variants associated with Nasu Hakola disease

3. Structural variants – disrupt expression

None

4. Significant association evidence with AD

Sims R et al., Nature Genetics

5. Replication in another population

R47H is much rarer in non-European populations so it has been difficult to establish association with disease.

6. Gene expressed in an AD-relevant tissue/cell type

TREM2 is expressed in myeloid cells, including microglia

7. Supporting evidence

a. Multiple rare variants significantly associated with AD

Gene-based tests demonstrate that multiple rare variants are associated with AD. R62H achieves genome-wide significance

b. Common variants associated with the same gene

An AD risk locus maps close to or within the TREM2 locus

8. Pleiotropy

Homozygosity for Loss of function variants in TREM2 results in a leukodystrophy called Nasu Hakola Syndrome associated with frontotemporal dementia and bone cysts. Some association with other neurodegenerative diseases but these have not been consistently replicated

9. Rare variants in the same gene in another ethnic group

Yes, association in African Americans. (Jin et al., 2015)

10. Co-segregation in families

Yes

11, .Other?

Animal and cell models of TREM2-/- and knock-in of TREM2 R47H have been created and crossed to 5XFAD Aß models. Loss of TREM2 and R47H results in fewer microglia around Aß plaques and change in structure of the plaque. SNPs in TREM2 and MS4A locus are associated with soluble TREM2 levels in CSF.

*ZNF423*

Locus – Data to collect

Gene: ZNF423

Evaluator: Lindsay Farrer

Date: 10/2/19

A. Significance

1. discovery study

- study size (#cases/controls) -- 3,332 cases, 9,832 controls (GERAD)^a^
- evaluation of population heterogeneity (yes/no/comments) – presumably yes
- multiple test/model corrections (yes/no/comments) – yes (gene-based p = 2.1x10^-6^ which is gene-wide significant after correcting for 18,087 tests, i.e., threshold p = 2.76x10^-6^ )

2. meta-analysis – discovery study -- **N/A**

- study size (#cases/controls)
- evaluation of population heterogeneity (yes/no/comments)
- multiple test/model corrections (yes/no/comments)

3. replication studies – NO REPLICATION STUDIES, however result available from another study including UK Biobank (proxy phenotype) and IGAP (GERAD portion overlaps with discovery study) with a total sample of 314,278^b^

gene-based p =0.43 (not significant)

- independent investigators and data - **RARE**
- larger study with added samples

4. replication studies – evidence -- **N/A**

- signal direction
- other ethnic groups study size (#cases/controls)
- evaluation of population heterogeneity (yes/no/comments)
- multiple test/model corrections (yes/no/comments)

5. Pleiotropy

5. AD-related phenotype (*e.g*. CSF biomarker association)

- ***ZNF423* resides in an AD-specific protein network^c^**

6. Non-AD phenotype (*e.g*. coronary artery disease, *APOE*)

- Recommended tier – **no higher than tier 3, perhaps lower**

Supporting references –

1. Baker et al. Gene-based analysis in HRC imputed genome wide association data identifies three novel genes for Alzheimer's disease. PLoS One 2019; 14(7):e0218111
2. Marioni RE, Harris SE, Zhang Q, McRae AF, Hagenaars SP, Hill WD, Davies G, Ritchie CW, Gale CR, Starr JM, Goate AM, Porteous DJ, Yang J, Evans KL, Deary IJ, Wray NR, Visscher PM. GWAS on family history of Alzheimer's disease. Transl Psychiatry 2018; 8(1):99.
3. Hu YS, Xin J, Hu Y, Zhang L, et al. Analyzing the genes related to Alzheimer’s disease via a network and pathway-based approach. Alzheimers Res Ther. 2017; 9(1):29
